# Modeling the impact of respiratory disease outbreaks on the United States agricultural workforce

**DOI:** 10.64898/2026.03.31.26349871

**Authors:** Katherine Bardsley, Luis X. de Pablo, Emma Keppler Canada, Naia Ormaza-Zulueta, Zia Mehrabi, Stephen M. Kissler

**Affiliations:** Department of Ecology and Evolutionary Biology, University of Colorado Boulder, Boulder CO USA; Department of Psychology and Neuroscience, University of Colorado Boulder, Boulder CO USA; Department of Environmental Studies, University of Colorado Boulder, Boulder CO USA; Department of Computer Science, University of Colorado Boulder, Boulder CO USA; Department of Epidemiology, Colorado School of Public Health, Aurora CO USA

## Abstract

**Background:** Emerging respiratory disease outbreaks pose a major threat to food production systems. Agricultural workers face health disparities that amplify their susceptibility to respiratory pathogens, yet the consequences for worker health and food production remain poorly understood.

**Methods:** We developed a household-structured susceptible-infectious-recovered (SIR) transmission model to compare disease dynamics between agricultural workers and the general U.S. population across six regions. We simulated outbreaks across a range of epidemiological scenarios and assessed productivity losses in California for three labor-intensive crops (oranges, iceberg lettuce, strawberries) with different harvest seasonalities.

**Results:** We show that, for a baseline reproduction number of *ℛ*_0_ = 2.0, the peak disease prevalence among agricultural workers is 1.41–1.69 times higher than that of the general population across regions, and final outbreak sizes are 1.29–1.42 times higher. Household structure accounts for 54%– 68% of the disease disparity. Peak productivity losses range from 0.50%–0.63% across crops, translating to millions in lost potential revenue. At higher transmissibility and severity (*ℛ*_0_ = 3 and assuming all infections are symptomatic), losses are 2.4 times higher.

**Conclusions:** Household crowding may lead to disproportionate respiratory disease burden among agricultural workers that is exacerbated by vaccination and obesity disparities, highlighting the need for targeted outbreak preparedness and mitigation strategies in the agricultural sector to maintain food system resilience and support public health in these communities.

## Introduction

Respiratory disease outbreaks can cause societal and economic disruptions that cascade beyond their direct public health burden. The food system, which encompasses food production, processing, and distribution, is particularly susceptible to labor-driven shocks.^1,2^ During the COVID-19 pandemic, agricultural labor shortfalls contributed to an estimated $309M reduction in U.S. farm output;^3^ meanwhile, outbreaks in meat packing plants forced widespread facility closures,^1,4^ and the closure of schools and restaurants caused distribution bottlenecks and shortages of staple goods.^5^ Recent infections of H5N1 influenza in dairy workers underscore the ongoing threat posed by respiratory viruses.^6^

The U.S. agricultural workforce, consisting of approximately 3.2 million people,^7^ sits at the foundation of the food system, and their health determines its capacity to function. Beyond their role in sustaining the food supply, maintaining the health of agricultural workers is clearly important in its own right, especially since agricultural workers are often more socially and medically vulnerable than the general population. Many of these health vulnerabilities put them at increased risk during respiratory disease outbreaks. Agricultural workers often have restricted access to healthcare,^8^ high exposure to hazardous work conditions such as pesticides and heat stress,^9^ high rates of chronic conditions including diabetes^10^ and obesity,^11^ along with high rates of baseline respiratory illnesses.^12,13^ Vaccination rates also tend to be low among agricultural workers.^14^ Furthermore, housing conditions, including household size and crowding, may contribute to poorer health and elevated risk of infection.^15–18^ Household crowding was a primary risk factor for SARS-CoV-2 infection among farmworkers^16^ and is associated with higher risk of COVID-19, influenza, and RSV infection and hospitalization.^19–24^ Together, this may help explain why COVID-19 infection rates were found to be consistently higher among agricultural workers *vs.* the broader community during the first year of the COVID-19 pandemic.^25,26^ Importantly, these documented disparities may under-estimate the true health challenges faced by agricultural workers, since informal and undocumented agricultural workers are often not covered by standard data collection mechanisms, despite constituting an estimated 40% of hired crop farmworkers.^27^

Conducting effective public health surveillance — *i.e.,* monitoring disease incidence for proactive maintenance of health and well-being — for agricultural workers has proven challenging.^28,29^ Various efforts are underway to bridge this gap, including the CDC-supported network of farmworker-serving organizations formed during the COVID-19 pandemic.^29–33^ Nevertheless, agricultural workers remain a challenging population to reach *via* traditional public health mechanisms, especially for those who are undocumented or on temporary visas. Furthermore, socioeconomic barriers often prevent workers from seeking care; and seasonal migration complicates longitudinal follow-up.^28,33^ In addition to improved data collection, modeling tools can help estimate the burden of disease and its drivers among agricultural workers from more readily available community-level data, helping to estimate risk in data-sparse environments and to inform more tactical data collection itself. Some efforts have begun to bridge this gap: compartmental models (*e.g.,* SIR) have demonstrated how contact patterns among essential workers translate into elevated infection risk,^34^ agent-based simulation tools have been developed to evaluate outbreak control strategies within individual food production facilities,^35^ and retrospective analyses have linked agricultural labor losses during the COVID-19 pandemic to reductions in farm output.^3^ However, we still lack a generalizable, population-level framework that mechanistically translates individual- and household-level risk factors into projections of how future respiratory disease outbreaks may affect agricultural workers and what the downstream consequences for food production may be. This has prevented us from identifying which structural differences between agricultural workers and the general population contribute most to infectious disease disparities, and from evaluating which interventions — ranging from immediately actionable efforts like targeted data collection and increasing vaccination rates, to longer-term efforts like reducing obesity and improving housing conditions — could most effectively mitigate the risks to worker health and food production posed by respiratory viruses.

This study aims to address these gaps by developing a household-structured disease transmission model that quantifies the differential impacts of respiratory disease outbreaks on agricultural workers relative to the general U.S. population. We focused on household structure, vaccination, and obesity as well-documented and quantifiable modulators of respiratory virus transmission and disease. We used household size and crowding distributions from national surveys to simulate epidemic dynamics under a range of epidemiological scenarios, assessing the impact of differences in transmissibility, within-household secondary attack rates, and population mixing patterns. We then translated the resulting labor impacts into crop-specific production losses. The impacts of labor shortfalls on agricultural output are likely to be greatest for labor-intensive, hand-harvested crops, where mechanization cannot easily compensate for worker absences. We therefore focus on California, which produces nearly two-thirds of US fruits, vegetables, and nuts,^36^ and examine three labor-intensive crops — strawberries, iceberg lettuce, and oranges — for which the state dominates national production.^37–39^ These crops also follow distinct harvest seasonalities, allowing us to assess how outbreak timing interacts with production vulnerability.

We find that, for a baseline reproduction number of ℛ_0_= 2.0, the peak disease prevalence among agricultural workers is 1.41–1.69 times higher than that of the general population across regions, and final outbreak sizes are 1.29–1.42 times higher. Household structure accounts for 54%–68% of the disease disparity. Peak productivity losses range from 0.50%–0.63% across crops, translating to millions in lost revenue. At higher transmissibility and severity (ℛ_0_ = 3 and assuming all infections are symptomatic), losses are 2.4 times higher. Our analysis provides a quantitative assessment of how outbreaks with varying characteristics might impact agricultural production, along with a general framework for anticipating the food system consequences of future respiratory disease outbreaks and designing effective interventions.

## Methods

### Data

#### Population characteristics

We obtained county-level data on overall population size, household size distribution (proportion of households of size 1, 2, 3, 4, 5, 6, or 7+), proportion of crowded households (i.e., with more than one individual per room), and proportion of agricultural workers from the U.S. Census Bureau’s 2022 American Community Survey (ACS) 5-year estimates.^40^ For agricultural workers specifically, we obtained regional data on household size distribution and proportion of crowded households from the 2018-2022 National Agricultural Workers Survey (NAWS).^41^ The NAWS data are stratified geographically into six regions: East, Southeast, Midwest, Southwest, Northwest, and California. To enable region-level analysis, we aggregated the county-level ACS data into the corresponding NAWS regions using population-weighted averages. Full details on the data extraction are given in the **Supplementary Methods.**

#### Crop harvest calendars and labor requirements

To approximate daily harvest volumes, we obtained data on specialty crop movements (point-to-point shipments) for strawberries, iceberg lettuce, and oranges from the United States Department of Agriculture’s (USDA’s) Agricultural Marketing Service.^42^ Data for six additional crops (apples, avocados, cherries, grapefruit, grapes, and lemons) were extracted from the same source and integrated into the interactive modeling tool. We extracted the total weekly weight of shipments originating in California for each of these crops between 1 Jan 2018 and 1 Jan 2025. We averaged the weekly shipment volumes for each crop across the seven available years to mitigate the impact of inter-annual variation. Then, we interpolated daily shipment volumes by assuming equal shipment volumes across each day of the week. We normalized these shipment volumes by the total annual shipment volume, so that the daily values reflected the proportion of the average annual harvest normally collected on that day. We cross-referenced the resulting production curves with independent reports on each crop’s production timing, confirming that the crop movement data are a reasonable proxy for seasonal harvest patterns^43–45^ (Supplementary Methods, Supplementary Figures S18-S19). The annual economic value of strawberries, oranges, and head lettuce (of which iceberg lettuce is a variety) produced in California were obtained from USDA National Agricultural Statistics Service reports capturing crop values in 2024, and used to normalize shipment volumes to actual harvests.^37–39^

### Disease transmission model

#### Model structure

We simulated respiratory disease transmission using a deterministic household-structured susceptible-infectious-recovered (SIR) model based on a previously developed framework.^46^ We split the population at the household level into “agricultural workers” and “general community”, assuming the proportion of households belonging to agricultural workers was equal to the proportion of the working population involved in agricultural work according to the ACS data.

The model tracks the number of infections over time in the population, explicitly accounting for transmission within and between households of various sizes (**Supplementary Figure S1**). The transmission model has three main epidemiological parameters: the between-household transmission rate (*β*), the within-household transmission rate (*τ*), and the recovery rate (*γ*). We allowed the within-household transmission rate *τ* to differ for uncrowded vs. crowded households, and we scaled τ by 1 – VC_x_ VE, where VCx is the vaccination coverage in the corresponding population group (agricultural workers [A] or general community [C]), and VE is the vaccine effectiveness.

We assumed that mixing among agricultural workers (A) and the general community (C) was assortative, governed by an assortativity parameter *η*, where *η* = 1 implies completely assortative mixing and *η* = 0 implies mixing proportional to each sub-population’s size. Following standard practice for disease transmission modeling in structured populations,^47^ we modeled these interactions using the mixing matrix

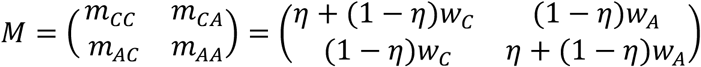

Here, *w_C_* is the fraction of the region’s population made up by the general community and *w_A_* is the fraction of the population made up by agricultural workers. This matrix modulates the between-household force of infection *λ* experienced by each population such that

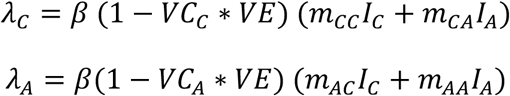

where *λ_i_* is the between-household force of infection for members of sub-population *i*; *β* is the between-household transmission constant; VC_C_ and VC_A_ are the vaccination coverage rates for the general community and agricultural workers, respectively; VE is the vaccine effectiveness; and *I_i_* is the proportion of infectious individuals in sub-population *i*. For full details on the model structure, see the **Supplementary Methods.**

#### Model parameterization

Following previous methods,^46^ we began by fixing the recovery rate *γ* = 1/5, which corresponds to a mean infectious period of 5 days, consistent with the infectious period of influenza.^48^ In sensitivity analyses, we considered infections periods between 3 and 10 days 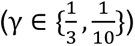. Then, given *γ* and the household secondary attack rate (SAR) (i.e., the probability that a household contact of an infected index case becomes infected themselves), we derived *τ* (**Supplementary Methods**). We set the SAR for uncrowded households at 20%, following estimates for influenza.^49–51^ For crowded households, we set the baseline SAR at 40%, reflecting approximately twice the uncrowded rate. Available evidence suggests that household crowding may increase the risk of respiratory infection by 1.2-to 3.3-fold,^52,53^ though estimates vary by pathogen and outcome measure. We therefore treat this parameter as uncertain and vary the crowded-household SAR from 20% to 60% in sensitivity analyses. Last, given values for *γ* and *τ*, we numerically identified the value of *β* that would achieve a desired basic reproduction number (ℛ_0_) when simulating outbreaks at the national level. Specifically, for a candidate *β* value, we ran the model with a single sub-population to equilibrium; then, we compared the outbreak’s final size with the theoretical prediction from the implicit relationship *R*(∞) = 1 − exp(−ℛ_0_ ⋅ *R*(∞)), where *R*(∞) is the final size of the outbreak.^54^ We adjusted *β* using a bisection search algorithm until the simulated final size was within 0.0005 of the theoretical value. We set the basic reproduction number to ℛ_0_ = 2, within the range of estimates for the 1918 influenza pandemic (median 1.80, IQR 1.47–2.27)^55^. Because a fraction of each population is vaccinated at baseline, the corresponding community effective reproduction number is ℛ*_eff_* ≈ 1.4 (1.40–1.50 across regions), consistent with estimates for the 2009 H1N1 pandemic (median 1.46, IQR 1.30–1.70)^55^. We computed ℛ*_eff_* from the simulated community final outbreak size using the SIR final-size relation ℛ*_eff_* = -ln(1 - ℛ_∞_)/ℛ_∞_, the same relation used to define ℛ_0_. In sensitivity analyses, we considered ℛ_0_values of 1.5 and 3.0. Baseline and sensitivity parameter values are listed in **Supplementary Table S2**.

We set the baseline assortativity parameter at η = 2/3, reflecting moderate preferential mixing within population groups. Empirical estimates of occupational assortative mixing are not available for agricultural workers, so we chose a baseline value reflecting the assumption that the majority of contacts occur within-group while allowing for substantial between-group mixing. Because this parameter is poorly constrained, we explored values of η ranging from 0 (proportionate mixing) to 3/4 (strong assortativity) in sensitivity analyses. Because agricultural workers constitute a small fraction of the total population (*w_A_* ranges from 0.7% to 2.2% across regions), the proportion of within-group contacts for agricultural workers is approximately equal to *η* (*m_AA_* = *η* + (1 − *η*)*w_A_* ≈ *η*, since *w_A_* is small). At the baseline *η* = 2/3, agricultural workers have between 66.9% and 67.4% of their contacts within their own group, depending on the region. The general community, being much larger, has nearly all contacts within their own group regardless of *η* (*m_CC_* ranges from 97.8% to 99.8% across all regions and values of *η*). At *η* = 0 (proportional mixing, and the first extreme of the sensitivity analysis), agricultural workers have between 0.7% - 2.2% of their contacts within their own group, depending on the region; at *η* = 3/4 (the second extreme of the sensitivity analysis), this rises to between 75.2% and 75.5%.

We incorporated vaccination into the model as a leaky vaccine that reduces the per-contact probability of infection. Specifically, we multiplied the force of infection experienced by each sub-population by a vaccination multiplier, 1 – VC_x_ VE, where VC_x_ is the vaccination coverage in subpopulation *x* and VE is the vaccine efficacy (the fractional reduction in per-contact transmission risk among vaccinated individuals). The between-household transmission rate *β* was calibrated in the absence of vaccination, so that ℛ_0_ represents the intrinsic transmissibility of the pathogen prior to any vaccination effect. For baseline values we used VE = 0.6, consistent with influenza vaccine effectiveness estimates in a good year;^56^ VC_C_ = 0.5 for the general community, reflecting U.S. influenza vaccine uptake;^57^ and VC_A_ = 0.4 for agricultural workers, reflecting the lower uptake in this population.^14^ In sensitivity analyses, we varied VC_A_ across {0.2, 0.4, 0.6, 0.8}, VC_C_ across {0.3, 0.4, 0.5, 0.6}, and VE across {0.2, 0.4, 0.6, 0.8}.

For the purposes of computing workforce absenteeism, we assumed symptoms began one day after infection and lasted for three days, consistent with (and on the conservative side of) findings that symptomatic influenza infections can cause 3–5 days of absenteeism from work. .^58,59^

We modeled the effect of obesity as an adjustment to the proportion of infected agricultural workers who develop work-limiting symptoms. We anchored the community-wide symptomatic probability at *p*_symp,C_ = 0.5, consistent with estimates of the asymptomatic rate of influenza^60,61^. We assumed that the probability of developing symptoms given infection differs between obese and non-obese individuals according to an odds ratio (OR), so that the obese and non-obese symptomatic probabilities (*p*_symp | ob_ and *p*_symp | nob_, respectively) satisfy

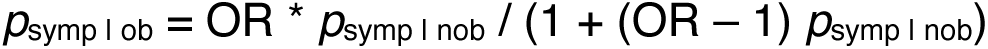

and the overall symptomatic fraction for a sub-population x with obesity prevalence *p*_ob,x_ is *p*_symp,x_ = p_ob,x_ *p*_symp | ob_ + (1-*p*_ob,x_) *p*_symp | nob_. Setting *p*_symp,C_ = 0.5 and *p*_ob,C_ = 0.4 for the general community gives a quadratic in *p*_symp | nob_, which we solved analytically to obtain *p*_symp | nob_ and *p*_symp | ob_. We then applied these to compute the agricultural worker symptomatic fraction using their obesity prevalence.

At baseline, we set OR = 1.5 and *p*_ob,C_ = 0.4 for the general community.^62^ The odds ratio of developing work-limiting symptomatic illness given obesity has not been directly characterized for influenza; we adopt OR = 1.5 as a baseline informed by evidence that obese individuals face approximately 1.5x higher risk of hospitalization from seasonal influenza than non-obese persons^63^ — hospitalization being a more severe endpoint than symptomatic illness, making this estimate conservative. We set *p*_ob,A_ = 0.55 as the baseline obesity prevalence among agricultural workers,^64^ yielding *p*_symp,A_ = 0.515 for agricultural workers. The community obesity prevalence was held fixed at 0.4 throughout all the analyses. In sensitivity analyses we varied OR across {1, 1.5, 3} and *p*_ob,A_ for agricultural workers across {0.4, 0.5, 0.55, 0.6, 0.7}.

Since production losses scale linearly with *p*_symp,A_, results for any agricultural worker symptomatic fraction can be obtained by rescaling from the *p*_symp,A_ = 1 upper bound (**Supplementary Table S6**).

The transmission model requires knowing the fraction of households of each size are crowded, but the ACS and NAWS data report household size distribution and crowding proportion separately. To assign crowding levels by household size, we assumed that the crowding probability increased linearly from households of size 2 to households of size 7+ (since households of size 1 by definition cannot be crowded). We obeyed the constraints that (1) the overall proportion of crowded households must match the ACS- or NAWS-reported proportion, and (2) households of size 7+ are *d* times as likely to be crowded as households of size 2 (**Supplementary Methods**). We used a baseline of *d* = 2 (i.e. households of size 7+ are twice as likely to be crowded as households of size 2). In sensitivity analyses, we considered *d* = 1 and *d* = 3.

#### Decomposition analysis

To quantify the relative contributions of household structure, vaccination coverage, and obesity to the disease disparity between agricultural workers and the general community, we conducted a factor decomposition analysis. We ran four epidemic scenarios at baseline parameter values using a one-at-a-time design: a *null* scenario, in which agricultural workers were assigned the general community’s household size distribution and vaccination coverage (*i.e.*, all factors equalized); a *household structure* scenario in which agricultural workers retained their NAWS-derived household size and crowding distributions but were assigned the community vaccination rate; a *vaccination* scenario in which agricultural workers retained their baseline vaccination coverage but were assigned the community household distribution; and a *full* scenario replicating the main analysis with all factors at their baseline, population-specific values. Because obesity affects the proportion of infected workers who develop work-limiting symptoms but not the transmission dynamics, its contribution was computed analytically *post hoc* by applying either a common symptomatic fraction (*p*_symp,C_ = 0.5, equalizing the factor) or the agricultural-worker-specific fraction (*p*_symp,A_ = 0.515, retaining the disparity) to the results of each epidemic scenario. This yielded eight total scenario combinations.

#### Model implementation

We implemented the transmission model in **`R`** (version 4.5.0) using **`odin`** (version 1.2.7). We initialized outbreaks by setting 0.1% of individuals in each sub-population as infectious (results were virtually unchanged when seeding the epidemic in just one of the two subpopulations). We distributed the initial infectious individuals proportionally across household types, approximating uniform random seeding of infections across each sub-population. We simulated outbreaks over 365 days.

In addition to the main regional simulations, we also generated county-level simulations to explore differences in transmission between agricultural workers and the general population at a finer geographic scale. Since the NAWS dataset does not report at the county level, we imputed county-level household size distributions and crowding proportions for agricultural workers using three different methods: an “additive” method, where county-level NAWS values were imputed by adding the difference between the county-level and regional ACS (general community) values to the regional NAWS value; a “multiplicative” method, where regional NAWS values were scaled by the ratio of county-level to regional ACS values; and a “null” method, where regional NAWS values were used without adjustment (**Supplementary Methods**). The “null” method yielded no variation in county-level agricultural household characteristics within a region; the “additive” method yielded an intermediate amount of variation; and the “multiplicative” method yielded a high amount of variation (**Supplementary Figures S3–S4**). We treated the “additive” method as a baseline and considered the “null” and “multiplicative” methods in sensitivity analyses. Due to the high uncertainty in these imputation methods, we emphasize that these county-level results have a low level of confidence and are intended to provide a rough estimate of within-region variation around the regional mean.

### Outcomes and measurements

We compared the difference in household size distribution (mean household size, proportion of households with 4 or more people) and the difference in household crowding rates between agricultural workers and the general community at the region level. For the outbreak simulations, we measured differences in peak prevalence, time to peak, final size, and maximum incidence deviation between agricultural workers and the general community.

To assess each factor’s impact on the disease disparity between agricultural workers and the general population, we used the results of the decomposition analysis: we measured each factor’s contribution to the A–C disparity in cumulative symptomatic cases as the disparity observed when only that factor differed between the two sub-populations, expressed as a percentage of the full model disparity. The remaining percentage, attributable to interactions among factors, was computed as the difference between 100% and the sum of the three single-factor contributions.

To translate disease dynamics into agricultural productivity impacts, we estimated the number of workers unable to perform harvest labor each day due to symptomatic illness. We assumed that symptomatic individuals could not perform agricultural labor. This allowed us to calculate a daily “workforce strength”, consisting of the fraction of agricultural workers still healthy. We multiplied this workforce strength by the daily harvest fraction for each crop to obtain an outbreak-adjusted harvest volume and summed this adjusted volume over the full year to measure the agricultural impact of the outbreak. We did this for outbreaks peaking on each day of the year to assess the impact of outbreak timing on agricultural productivity for each crop.

### Statistics and Reproducibility

All epidemic simulations are deterministic; the results contain no stochastic variation and are fully reproducible given the parameter values and initial conditions in **Supplementary Table S2.** The model was implemented in **`R`** (version 4.5.0) using **`odin`** (version 1.2.7).; all code is available at https://github.com/skissler/AgriResp.

To characterize uncertainty and assess robustness of findings, we conducted one-at-a-time sensitivity analysis, varying each epidemiological parameter in turn while holding all others at baseline values. Five sensitivity axes were explored: *R*_0_ ∈ {1.5,2.0,3.0} ; assortativity 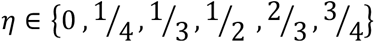; crowded-household secondary attack rate (SAR) ∈ {20%, 30%, 40%, 50%, 60%}; fold-difference in crowding probability for large *vs.* small households *d* ∈ {1,2,3}; and mean infectious period 1/*γ* ∈ {3,5,10} days. For vaccination and obesity parameters, we additionally varied vaccine efficacy *VE* ∈ {0.2,0.4,0.6,0.8} ; community vaccination coverage *VC_C_* ∈ {0.3,0.4,0.5,0.6}; agricultural worker vaccination coverage *VC_A_* ∈ {0.2,0.4,0.6,0.8}; agricultural worker obesity prevalence *p_ob_*_,*A*_ ∈ {0.40,0.50,0.55,0.60,0.70}; and odds ratio of symptomatic disease given infection while obese *OR* ∈ {1,1.5,3}. All baseline values and the full ranges explored are listed in **Supplementary Table S2**.

County-level ACS data were aggregated to NAWS regions using population-weighted averages. For county-level simulations, we compared three imputation approaches for agricultural worker household characteristics (null, additive, and multiplicative; see **Supplementary Methods**) to assess sensitivity to this methodological choice. County-level results are reported as medians and 20^th^–80^th^ percentile ranges across counties within each region.

### Statement on the use of generative AI

Claude Code (Opus version 2.1.37) was used to improve code efficiency, generate sensitivity analyses, review code for logical consistency, format figures, and review the manuscript for agreement with code structure and output. Code for the central disease transmission model was written independently by the authors, and all code and manuscript text were reviewed and approved by the authors.

## Results

Household structure, vaccination rates, and obesity lead to higher modeled disease prevalence among agricultural workers.

Agricultural worker households are substantially larger and more crowded on average than in the general U.S. population (**Figure 1, Supplementary Table S1**). The mean household size for agricultural workers ranged from 3.3–4.1 people across the six U.S. regions for which data were available, compared to 2.4–2.8 people for the general population. The proportion of households of size 4 or greater ranged from 41%–62% among agricultural workers *vs.* 20%–30% for the general population. The proportion of crowded households ranged from 11.2%–32.8% for agricultural workers compared to 1.9%–8.3% in the general population. This translated into crowding rates that were 3.3–8.8 times higher among agricultural workers than the general population across the six regions. The California region had the highest household crowding rates both for agricultural workers (32.8%) and the general population (8.3%, population-weighted mean across counties).

**Figure 1.**
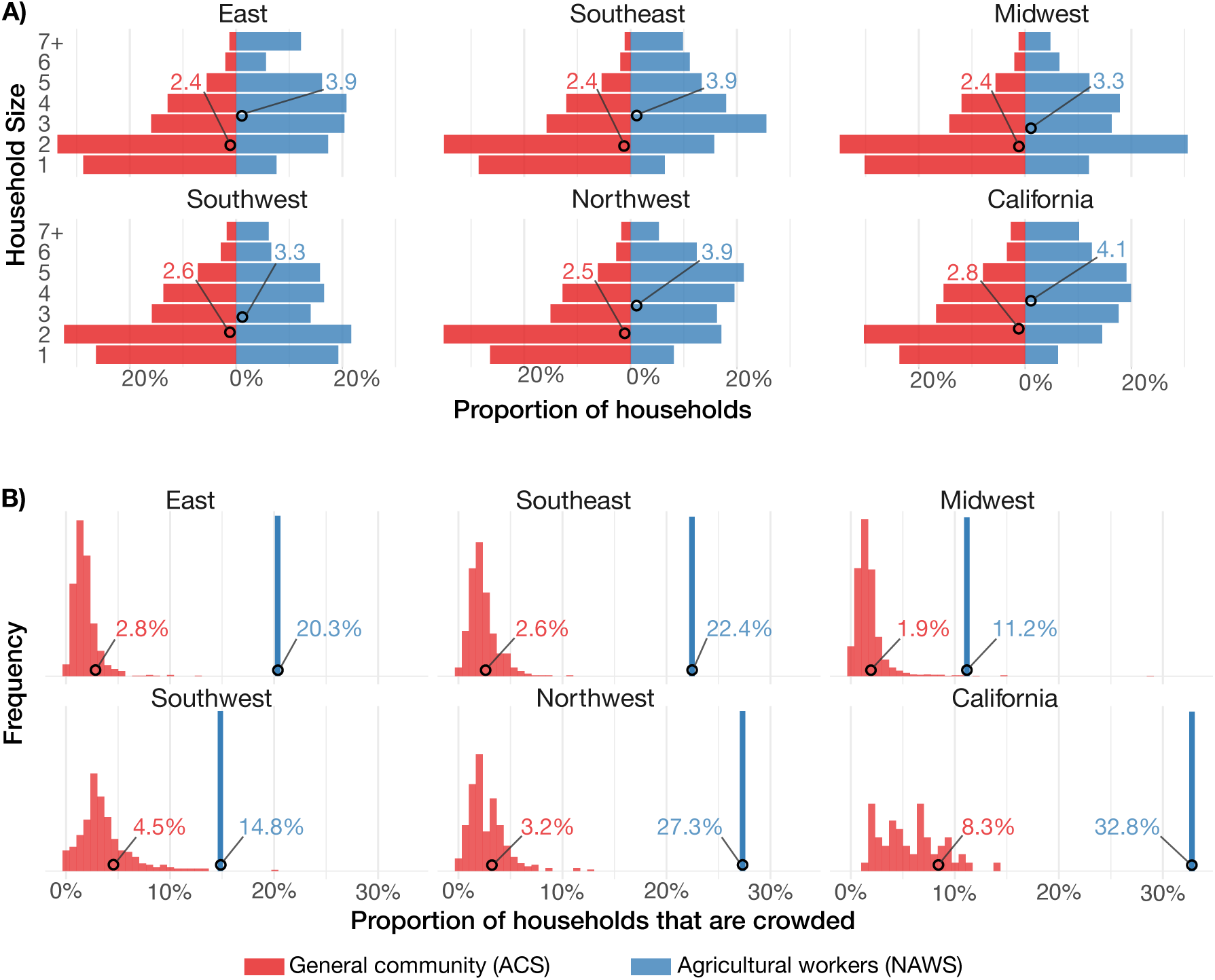
Household characteristics by region for agricultural workers and the general community. (A) Proportion of households of size 1 – 7+ for agricultural workers (blue) and the general community (red). Mean household sizes for each region and sub-population are depicted as circles. (B) Proportion of households that are crowded for agricultural workers (blue) and the general community (red). Histograms for the general community represent county-level differences in household crowding within each region. For agricultural workers, household crowding is available only at the region level, so these are depicted as single bars. Mean household crowding proportions for each region and sub-population are depicted as circles. Data for agricultural workers are extracted from the National Agricultural Workers Survey (NAWS) and data for the general community are extracted from the American Community Survey (ACS).

Outbreak simulations at the regional level revealed consistently higher disease burden among agricultural workers than in the general population (**Figure 2, Supplementary Tables S3-S4**). Under baseline assumptions (**Supplementary Tables S2**), peak prevalence among agricultural workers was 1.41–1.69 times that of the general population across regions. Final outbreak sizes were 1.29–1.42 times higher among agricultural workers, with final symptomatic attack rates of 34–39% among agricultural workers compared to 26–29% in the general population. At the point of maximum divergence between the two epidemic curves, prevalence among agricultural workers was 2.32–3.69 times that of the general community. Across regions, household size and crowding alone accounted for 54%–68% of the difference in final outbreak size; vaccination rates accounted for 26%–35% of the difference; and obesity accounted for 7–11% of the difference (**Supplementary Table S5, Supplementary Figure S2**).

**Figure 2.**
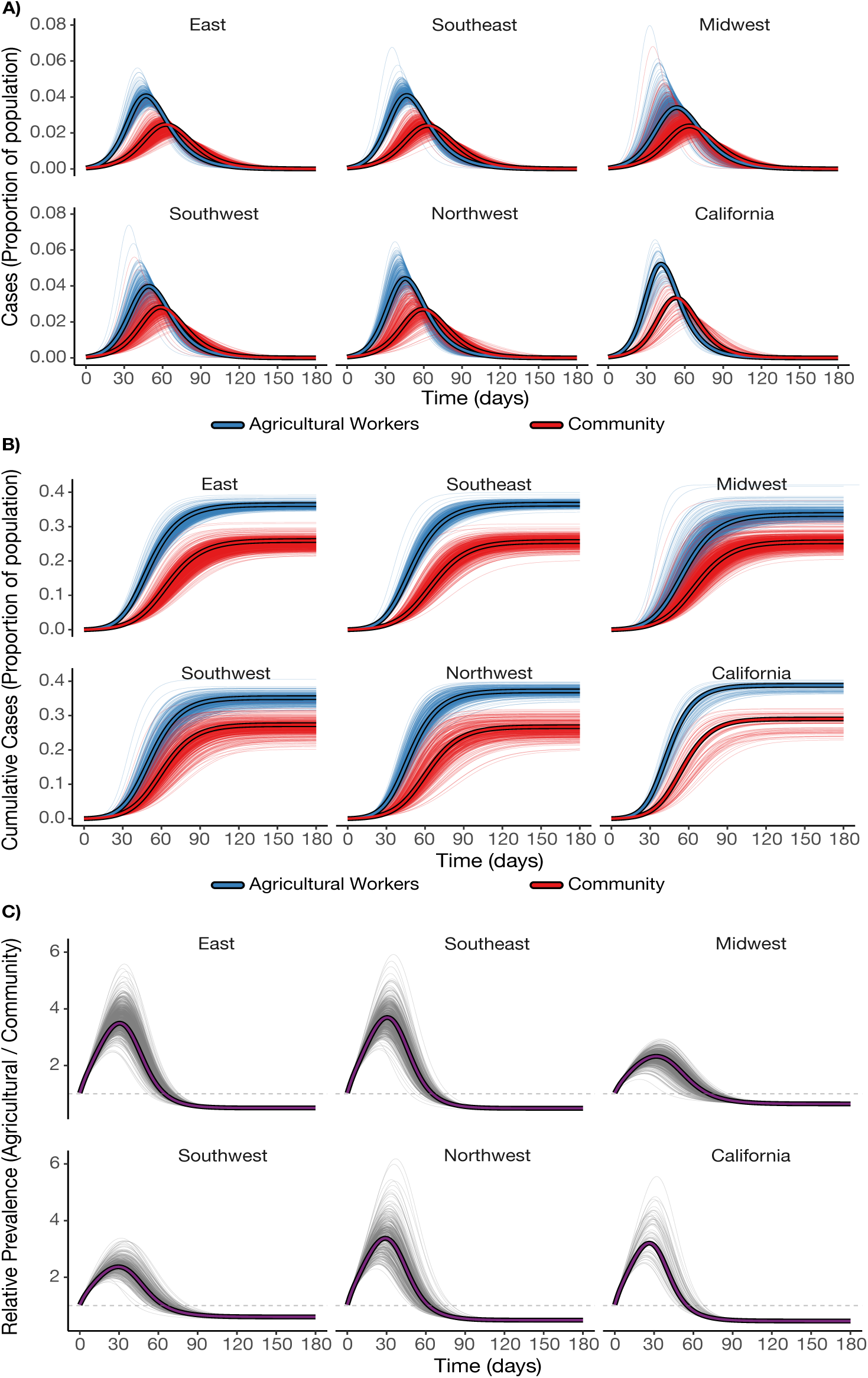
Simulated epidemic trajectories by region for agricultural workers and the general community. Infection prevalence (A) and cumulative infections (B) for agricultural workers (blue) and the general community (red), along with the ratio of agricultural worker to general community infection prevalence (C) in the six NAWS regions. Region-level simulations are depicted as thick lines with black outlines. County-level simulations are depicted as thin, partially transparent lines to illustrate within-region variation.

Differences in disease burden between agricultural workers and the general community were sensitive to the basic reproduction number (**Supplementary Figures S5-S6, Supplementary Tables S3-S4**). At a low reproduction number (ℛ_0_ = 1.5), the final outbreak sizes were most discordant between agricultural workers and the general community (1.99–3.21 times higher in agricultural workers across regions), and the peak prevalence ratio between agricultural workers and the general community ranged from 2.44–5.52 across regions. At ℛ_0_ = 3.0, final size ratios were 1.10–1.13 times higher in agricultural workers, while peak prevalence ratios were 1.19–1.26 times higher.

Increasing the secondary attack rate in crowded households generally led to greater differences in final size, peak prevalence, and peak timing between agricultural workers and the general community. Increasing assortativity (more within-group mixing) had a similar effect. Increasing vaccination rates in agricultural workers narrowed the gap in final size, peak prevalence, and peak timing, though parity was only achieved for vaccination rates of 60%. A long infectious period (10 days) extended the difference in peak timing between agricultural workers and the general community, and a short infectious period (3 days) compressed it, but final epidemic sizes were insensitive to the infectious period. The simulated epidemics were also largely insensitive to the fold-difference in crowding between the largest and smallest households (*d*) and to obesity-related factors (**Supplementary Figures S5–S15, Supplementary Tables S3-S4**).

County-level simulations demonstrated geographic heterogeneity in these infection disparities. Under the baseline parameter values, the median [20th, 80th percentile] county-level peak prevalence ratio ranged from 2.60 [2.14, 3.20] in the Southwest to 5.77 [4.25, 7.60] in the East. Similarly, the median [20th, 80th percentile] county-level symptomatic final size ratio ranged from 2.13 [1.84, 2.46] in the Southwest to 3.46 [2.81, 4.30] in the East. These results were sensitive to how county-level household characteristics were imputed for agricultural workers, with approaches that scaled household size and crowding rates according to the general population’s county-level variation producing wider ranges than approaches that assigned the regional value uniformly across counties (**Methods; Supplementary Figures S16–S17**).

Respiratory disease outbreaks among agricultural workers can lead to substantial productivity losses.

The simulated outbreaks yielded substantial productivity losses for all three crops we considered, with the impact varying by outbreak timing relative to peak harvest periods (**Figure 3, Supplementary Figure S20, Supplementary Table S6**). For the baseline analysis, we assumed half of infections caused work-limiting symptoms in the general community (*p_symp_* = 0.5), and we scaled this value to account for rates of obesity among agricultural workers based on its impact on increased symptom development (**Methods**). For strawberries, peak productivity losses (*i.e.*, assuming the worst possible outbreak timing and summing losses over that entire outbreak) were 0.63% with the worst outbreak timing being an epidemic peak in late May (day 150). For iceberg lettuce, maximum losses were 0.51% for outbreaks that peaked in early June (day 152). For oranges, peak losses were 0.50% for outbreaks peaking in early February (day 35). These translate into peak losses of roughly $21.6M, $6.3M, and $4.3M USD for strawberries, iceberg lettuce, and oranges, respectively. These findings were sensitive to ℛ_0_ and the symptomatic proportion (*p_symp_*), with a worst-case scenario (ℛ_0_ = 3, *p_symp_* = 1) yielding losses roughly 2.4 times higher than the base case (**Supplementary Table S6**).

**Figure 3.**
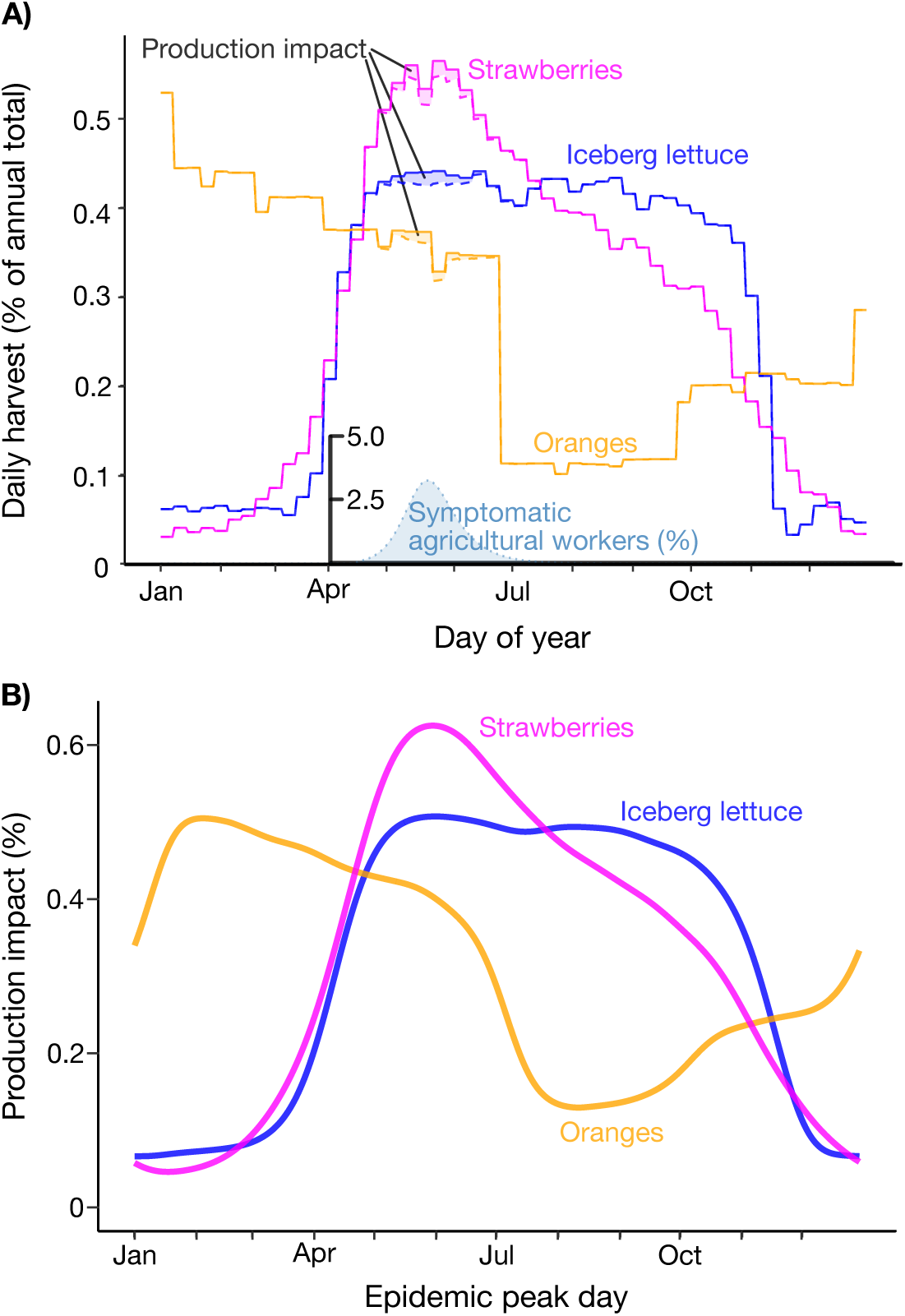
Simulated impact of a respiratory virus outbreak on harvesting of strawberries, iceberg lettuce, and oranges in California. (A) Illustration of the approach for calculating harvest impact. Here, an epidemic peaks in the general community on June 1st, leading to a peak in symptomatic disease among agricultural workers a few days earlier. The mean daily production of strawberries (magenta), iceberg lettuce (blue), and oranges (orange), averaged across 2018-2024, are depicted as solid lines. Dashed lines with shading depict the simulated production impact caused by the loss of labor due to symptomatic disease. The total impact (*i.e.*, the area of the shaded regions) is summed across the year, yielding a single point in plot (B) representing the overall impact of an epidemic peaking on June 1. (B) Simulated production impact on strawberries (magenta), iceberg lettuce (blue), and oranges (orange) for epidemics peaking in the general community on each day of the year.

## Discussion

Differences in household characteristics, vaccination rates, and comorbidities are sufficient to produce substantial disparities in the severity of respiratory disease outbreaks between agricultural workers and the general population. In our baseline scenario, representing a pandemic influenza-like virus (ℛ_0_ = 2.0), peak disease prevalence among agricultural workers was 1.4–1.7x higher than in the general community, with cumulative infections roughly 1.3–1.4x higher, depending on the region. At the point of maximum divergence, prevalence among agricultural workers was 2.3–3.7x higher than in the general community. Our findings indicate that community-wide disease indicators may substantially underestimate the burden of disease among agricultural workers, particularly during the early stages of an epidemic.

Disease among agricultural workers translates into lost labor and, consequently, reduced food production. For three labor-intensive California crops – strawberries, iceberg lettuce, and oranges — we estimated that a respiratory disease outbreak could reduce harvest volumes by 0.50%– 0.63% if its peak coincided with peak harvest periods, assuming roughly half of infections caused symptoms severe enough to prevent work, leaving food unharvested and lost. While these are modest reductions, they translate into estimated revenue losses of approximately $4.3–$21.6 million USD per crop, depending on the commodity. For a more transmissible or clinically severe pathogen, losses could be much higher: for example, at ℛ_0_ = 3 with a more clinically severe pathogen (*p_symp_* = 1), production losses would be substantially higher, impacting 1.21%–1.51% of the annual harvest for the labor-intensive crops we considered. Furthermore, these state-level averages mask potentially severe farm-level impacts; individual farms harvesting during concentrated windows could experience losses far exceeding the state mean if an epidemic coincides with their peak harvest.

Household size and crowding explained most of the disease burden disparity between agricultural workers and the general community. While vaccination and obesity rates also differ between the sub-populations, these were responsible for a smaller share of the disparity. Simulations indicate that increasing vaccination rates among agricultural workers to levels well above current influenza vaccination rates — but in line with COVID-19 vaccination rates in 2023^14^ — could largely eliminate the disparity in disease burden. Meanwhile, increasing the availability of adequate housing for agricultural workers and reducing the risk of indoor respiratory virus transmission (*e.g.*, by improving ventilation) offers a longer-term but potentially high-impact way of reducing the burden of not just respiratory disease, but also various other health conditions in agricultural workers.^15^

These findings are consistent with a growing body of evidence revealing the disproportionate impacts of respiratory disease on agricultural and food system workers. During the COVID-19 pandemic, prospective surveillance in California’s Salinas Valley found a SARS-CoV-2 test positivity rate of 22% among farmworkers, compared to 17% among other adults, with household crowding identified as a key risk factor.^16,25^ At a broader scale, COVID-19 led to an estimated 0.069% reduction in farm labor input across the United States, resulting in approximately $309 million in lost agricultural output.^3^ Our peak estimates of 0.50%–0.63% production loss for individual crops are several times higher, but the 0.069% figure encompasses all farm production (including less labor-intensive commodities) and reflects a pandemic whose surges were not necessarily aligned with peak harvest periods. We have focused on the impact of illness on agricultural workers, but elevated disease in agricultural workers may translate into higher disease rates for the general population: in U.S. counties with high concentrations of workers in fruit, vegetable, and horticultural production, COVID-19 rates were substantially elevated.^65^ Together, our results underscore the need for targeted interventions to protect the health of agricultural workers, both as a matter of equity and because of the downstream consequences for food production and overall population health.

Our baseline scenario represents an unmitigated outbreak of a novel respiratory pathogen with pandemic-scale transmissibility. The basic reproduction number (ℛ_0_= 2.0) falls within estimates for the 1918 influenza pandemic, and under baseline vaccination the corresponding community effective reproduction number is ℛ_eff_ ≈ 1.4, closely matching estimates for the 2009 H1N1 pandemic^55^. Under these assumptions the model projected a cumulative infection rate of 51%–58% in the general community, of which half (26%–29%) developed work-limiting symptomatic illness. These attack rates should be read as upper bounds: because the model assumes no pre-existing infection-acquired immunity (appropriate for a novel pathogen), homogeneous mixing within each sub-population, and no behavioral or public health mitigation, it yields final epidemic sizes near the theoretical maximum for a given reproduction number. Realized incidence in past pandemics has been considerably lower — an estimated 12%–25% of the U.S. population were infected during the 2009 H1N1 pandemic^67^, possibly owing to mitigation, population heterogeneity, and pre-existing immunity. Direct empirical validation of the agricultural-worker-specific predictions is not currently possible, as reliable data on respiratory disease incidence in this group are not available — a gap that is one of the central motivations for developing a model-based approach.

Various modeling frameworks have been developed to simulate the spread of COVID-19 among farmworkers^68,69^ and meat processing plant workers,^70,71^ but these have largely been retrospective, leaving a gap in prospective tools for anticipating how future respiratory disease outbreaks may unfold in agricultural communities. This study provides such a framework, explicitly linking household-level predictors of disease transmission to population-level epidemic dynamics and agricultural impacts. By design, our model isolates the contribution of household structure, vaccination rates, and obesity – risk factors for which population-level data are readily available — to differential disease burden. This makes it a natural baseline: any future model incorporating additional risk factors (e.g., occupational exposures, healthcare access, additional comorbidities) can be compared against these predictions to quantify the marginal contribution of each added factor. To accompany this study, we have developed an interactive simulation tool (https://kisslerlab.shinyapps.io/ag-epi-model/) to allow planners and researchers to simulate how outbreaks with different epidemiological characteristics and timing might affect various crops, supporting scenario-based preparedness efforts.

Our approach has several limitations. First, we did not incorporate seasonal variation in pathogen transmissibility (*e.g.*, higher transmission during winter months). Incorporating seasonality would likely increase the expected impact on oranges, which are primarily harvested from November through June, and decrease the expected impact on strawberries and iceberg lettuce, which are harvested mainly in the summer. Second, our crop impact analysis considered only harvest-phase labor. Outbreaks could also disrupt planting, tending, and post-harvest processing, as well as lagged production impacts through farm revenue losses, leading to additional production losses. More broadly, the impact of respiratory disease on the food system extends well beyond farm-level labor: the COVID-19 pandemic caused severe disruptions in meat and poultry processing facilities^1,4^ and demand-side effects such as panic buying and shifts from food service to at-home food consumption.^5^ Third, beyond household structure, vaccination coverage, and obesity, we did not account for occupational exposures that may independently elevate transmission risk among agricultural workers (such as shared transport, close-quarters fieldwork, pesticide exposure, and communal facilities), rates of chronic conditions beyond obesity (including diabetes), or limited access to healthcare, all of which could further compound the disparities we observe. Fourth, we assumed that labor losses during harvest translate directly into proportional production losses. In practice, this relationship is likely nonlinear in both directions. On one hand, many harvest operations require minimum crew sizes to function — for example, to staff a packing line or coordinate a picking crew — so production could drop more steeply than linearly if absences fall below a critical threshold. On the other hand, if additional workers are available, farms may compensate for absences by hiring replacement workers, extending shifts, or redistributing labor across tasks, partially buffering the impact. The linear impact assumption may be more defensible for crops with narrow harvest windows, like strawberries and iceberg lettuce which must be picked within days of maturity, than for oranges, which can remain on the tree for weeks, allowing workers to recover and make up for lost harvest time, although this comes with quality losses and likely revenue losses. Relatedly, presenteeism, which may be especially common among agricultural workers, could reduce the actual economic impact relative to what our model predicts.^72,73^ Fifth, data limitations constrained several aspects of the analysis. County-level household characteristics for agricultural workers are not available in the NAWS dataset, limiting the resolution of our geographic analysis. We assumed a linear relationship between crowding probability and household size, since these values are reported separately in available data; reassuringly, our results were largely insensitive to this choice. The assortativity of contacts between agricultural workers and the general community is poorly understood. Our use of point-to-point crop shipments as a proxy for harvest volumes may introduce a timing bias, particularly for crops like oranges that can be stored before shipment. We assessed the sensitivity of our findings to our assumptions about poorly constrained parameters, but our one-at-a-time sensitivity analysis does not capture potential interactions between parameters. Finally, our analysis assessed state-and region-level impacts, but farm-level impacts may differ considerably. Individual farms often harvest during concentrated windows, and an epidemic coinciding with such a window could devastate that farm’s production, while an epidemic at another time might have less impact. The smaller the geographic or operational scale, the more likely it is to see such “all or nothing” dynamics, and the synchrony in outbreaks across farms remains to be explored. Likewise, at smaller geographic scales, stochastic effects would play a larger role in shaping epidemic patterns, especially among the agricultural worker population that makes up a relatively small fraction of the overall population.

Effective disease mitigation among agricultural workers is essential both for protecting the health of this population and for safeguarding the food supply, yet it presents major unresolved challenges. Ultimately, reducing the vulnerability of agricultural workers to respiratory disease outbreaks will require structural interventions, including improvements in housing conditions, improved access to healthcare, provision of personal protective equipment, and paid sick leave policies, alongside the epidemiological tools for anticipating and responding to outbreaks that this study aims to provide.

## Author contributions

KB: study design, analysis, data interpretation, writing, editing. LXP: study design, analysis, data interpretation, writing, editing. EKC: study design, analysis, data interpretation, writing, editing. NOZ: study design, analysis, data interpretation, writing, editing. ZM: study design, analysis, data interpretation, writing, editing, project oversight. SMK: study design, analysis, data interpretation, writing, editing, project oversight.

## Data availability

All data associated with this manuscript can be accessed at: https://github.com/skissler/AgriResp. Data are public and available without restriction.

## Code availability

Code is available at https://github.com/skissler/AgriResp. Code was implemented in R (version 4.5.0) using odin (version 1.2.7). Key parameter values are listed in **Supplementary Table S2.** Code may be used and reproduced without restriction under the MIT license.

## Acknowledgements

The authors thank L. Hadley for comments on the manuscript.

## Supplementary Information

### Supplementary Methods

#### Data extraction

County-level household size distributions were obtained from American Community Survey (ACS) table B11016 (Household Type by Household Size), which reports counts of family and non-family households by size. We combined family and non-family counts for each household size (1 through 7+).

County-level household crowding proportions were obtained from ACS table B25014 (Tenure by Occupants per Room), where crowded households were defined as those with more than 1.00 occupants per room. We summed across owner- and renter-occupied units and all occupancy levels over size 1 (1.01–1.50, 1.51–2.00, and >2.00 persons per room). We normalized by the total population size (ACS table B01003).

To calculate the proportion of agricultural workers in a county using the ACS data, we extracted the number of individuals employed in “farming, fishing, and forestry occupations” (ACS occupation codes C24030_004 [males] and C24030_031 [females]) as a proportion of total employed individuals (C24030_001).

#### Data processing: calculating regional values for the general community

To enable region-level analysis, we aggregated the county-level ACS data into the corresponding National Agricultural Workers Survey (NAWS) regions using population-weighted averages. For each variable (household size proportions *p*, crowding proportions *q*, and proportion of agricultural workers *a*), we multiplied each county’s value by its population size, summed within each region, then divided by the total regional population size. Specifically, for each county *i* in region *r*, we computed:

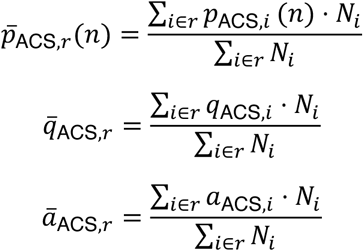

where *p*_ACS,*i*_(*n*) is the proportion of households with size *n* in county *i* according to the ACS data, *q*_ACS,*i*_ is the proportion of crowded households in county *i*, *a*_ACS,*i*_ is the proportion of agricultural workers in county *i*, and *N_i_* is the population size of county *i*. We re-normalized

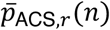

to ensure

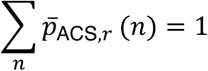

Household sizes for agricultural workers were derived from the NAWS D52 variable (total number of people sleeping in the housing unit). Households of size 7 or greater were grouped into a single “7+” category for consistency with the ACS data. Crowding status was derived from the CROWDED1 variable. Both household size and crowding data were weighted using the NAWS survey weights (PWTYCRD) and summarized by NAWS region.

#### Data processing: calculating crowding by household size

In both the ACS and NAWS datasets, household size and household crowding are reported separately, but for the disease transmission model, we required the proportion of households of a given size that are crowded. To assign crowding probabilities by household size, we used a linear relationship where larger households are progressively more likely to be crowded. For a household of size *n*, we defined a crowding multiplier:

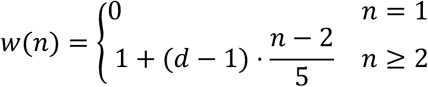

where *d* is a crowding fold-difference parameter (the ratio of crowding probability for size-7 households to size-2 households). For example, when *d* = 2 , *w*(*n*) = 0,1,1.2,1.4,1.6,1.8,2 for *n* ∈ 1,2, … ,7. Note that it is impossible for size-1 households to be crowded. We treated households of size 7+ as *n* = 7. For a given region and sub-population, the probability that a household of size *n* is crowded is then:

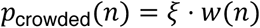

where the constant *ξ* is chosen so that the total proportion of crowded households in the region, 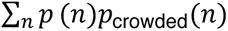, matches the observed fraction of crowded households, *P*_crowded_ (here, *p*(*n*) is the proportion of households that are size *n*). Specifically,

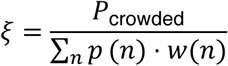

For example, for household size proportions *p*(*n*) = 0.1,0.2,0.3,0.2,0.1,0.05,0.05 for *n* ∈ 1,2, … ,7, and for an overall crowding fraction of

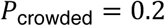

we have

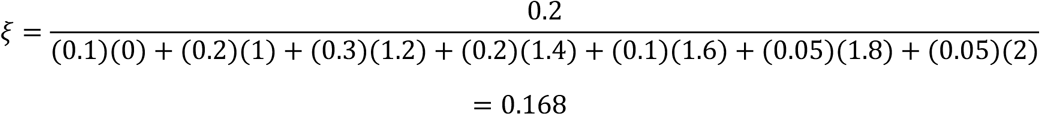

which ensures that

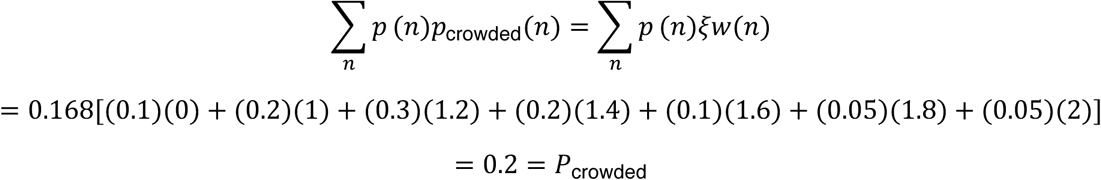

#### Data processing: imputing county-level household characteristics for agricultural workers

The NAWS dataset reports household characteristics for agricultural workers at the regional level only, while the ACS provides county-level data for the general population. While our main analysis was at the regional level, we also performed a county-level analysis to assess within-region variation in outbreak disparities between agricultural workers and the general community. To generate county-level population estimates for agricultural workers, we used county-level ACS variation to adjust the regional NAWS values. The underlying assumption is that county-level variation among agricultural workers follows a similar pattern to county-level variation in the general population; i.e., if a county’s general population has larger households than the regional average, agricultural workers in that county likely also have larger households than the regional average for agricultural workers.

We imputed county-level NAWS values using three methods (**Figures S2 and S3**):

*Additive method.* We shifted regional NAWS values by the difference between county-level and regional mean ACS values:

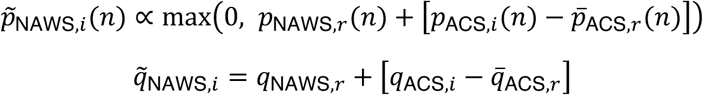

[Household size proportions were clamped to be non-negative before renormalization, and the crowding proportion was clamped to [0,1] (i.e., values below 0 were set to 0 and values above 1 were set to 1).

*Multiplicative method.* We scaled regional NAWS values by the ratio of county-level to regional mean ACS values:

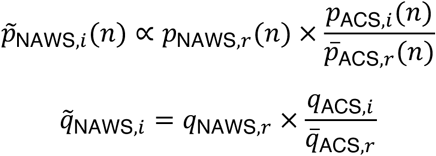

The household size distribution was renormalized to sum to 1, and the crowding proportion was clamped to the interval [0,1].

*Null method.* We used regional NAWS values directly without adjustment:

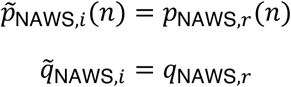

This method assumes no county-level variation in agricultural worker household characteristics within a region.

#### Data processing: assessing the validity of crop movements as a proxy for harvest

We used crop movements (point-to-point shipments) reported by the USDA in AMS origin-based movements as a proxy for harvest volumes of oranges, iceberg lettuce, and strawberries. Few available data sources capture crop-specific harvest volumes at sub-annual temporal resolution, whereas we needed up-to-date information on the seasonality of harvesting to assess the potential impact of epidemic timing on crop production. To validate the relationship between crop movements and harvests, we cross-referenced normalized average weekly crop movement volumes against independent harvest information for the same crops from University of California Agriculture and Natural Resources Cooperative Extension reports (**Figure S13**).^1–3^ For strawberries, the report gives the fraction of total annual harvest occurring in each month for the Central Coast Region of California (Santa Cruz, Monterey, and San Benito Counties): 5% in April, 12% in May, 25% in June, 26% in July, 18% in August, 12% in September, and 2% in October. For navel oranges, the report states that fruits in the San Joaquin Valley are “normally harvested from November to June”. For iceberg lettuce, the report states that planting in the Central Coast Region occurs “continuously from late December to mid-August” and that plants take up to 100 days to mature for cool-season plantings, with shorter maturation times in the warmer season.

We overlaid this information on the normalized crop movement data (**Figure S13**). For strawberries, we converted the monthly harvest proportions to approximate weekly rates by dividing the monthly harvest proportions by the number of weeks in the month (e.g., by 4.28 for a 30-day month). For oranges, we showed the November-to-June harvest window as a horizontal bar. For iceberg lettuce, we displayed both the planting window (late December to mid-August) as a lighter bar and an estimated harveouehost window (approximately late March to early October, obtained by shifting the planting window forward by 100 days at the cool-season end and 50 days at the warm-season end) as a darker bar. We do note differences in alignment between the crop movement data and the University of California reports^1–3^; for example, the crop movement data place peak strawberry shipments in late April and early June, whereas the University of California report indicates peak harvests in June and July; and iceberg lettuce shipments are shifted somewhat later than the estimated harvest window. These discrepancies may reflect limitations of using crop movements as a proxy for harvests, but may also reflect the fact that the movement data capture shipments from across the entire state of California, while the University of California reports^1–3^ pertain to specific sub-regions. With this caveat, we conclude that the crop movement data with origin in California provide a reasonable proxy for the seasonal pattern of crop harvests, that can be normalized to total harvested volumes.

#### Mathematical model structure

The household-structured SIR model (**Figure S1**) tracks the distribution of households across disease states. Let *H_k_*(*x*, *y*, *z*, *c*) denote the number of households in population *k* (where *k* ∈ {*C*, *A*} for community and agricultural populations) with *x* susceptible, *y* infected, and *z* recovered members, and crowding status *c* ∈ {0,1}. The total household size is *n* = *x* + *y* + *z*.

The dynamics are governed by three types of transitions:

- **Recovery transitions:** Infected individuals recover at rate *γ*, moving a household from state (*x*, *y*, *z*, *c*) to state (*x*, *y* − 1, *z* + 1, *c*): Recovery rate = *γ* ⋅ *y* ⋅ *H_k_*(*x*, *y*, *z*, *c*)
- **Within-household transmission:** Susceptible individuals are infected by household members at rate *τ_c_* = *τ*_base_ + *τ*_boost_ ⋅ *c*, moving a household from state (*x*, *y*, *z*, *c*) to state (*x* − 1, *y* + 1, *z*, *c*): Within-household infection rate = (1 − VC*_k_* ⋅ VE) *τ_c_* ⋅ *x* ⋅ *y* ⋅ *H_k_*(*x*, *y*, *z*, *c*)
- **Between-household transmission:** Susceptible individuals are infected through community contacts at rate *λ_k_*, determined by the mixing matrix and overall prevalence in each population: Between-household infection rate = *λ_k_* ⋅ *x* ⋅ *H_k_*(*x*, *y*, *z*, *c*)

The between-household force of infection for population *k* is: 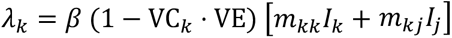 where *I_k_* is the prevalence in population 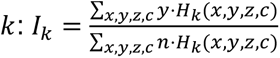. Throughout these equations, VC_k_ represents the vaccination coverage in population group *k* (agricultural workers or general community), and VE represents vaccine effectiveness. Vaccine protection is assumed to be “leaky”.

The mixing matrix elements are:

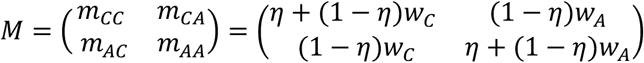

where is 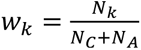 the population fraction in group *k*.

The complete system of ordinary differential equations is: 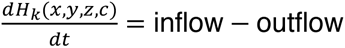

where inflow includes households transitioning into this state via infection (from state (*x* + 1, *y* − 1, *z*, *c*)) or recovery (from state (*x*, *y* + 1, *z* − 1, *c*)), and outflow includes households leaving this state through infection or recovery.

#### Mathematical model parameterization

We derived the within-household transmission rate, *τ*, using the recovery rate *γ* and the secondary attack rate (SAR). The SAR is determined by the competing rates of within-household infection (*τ*) and recovery (*γ*). The probability that the susceptible individual is infected before the infectious index case recovers is:

Solving for *τ*:

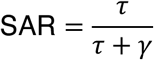

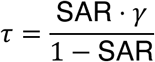

For the baseline uncrowded SAR of 20% and *γ* = 1/5:

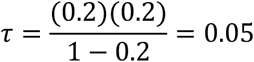

For crowded households, we computed *τ*_crowded_ using the same formula with the crowded SAR, then defined *τ*_boost_ = *τ*_crowded_ − *τ*. For the baseline crowded SAR of 40%: *τ*_crowded_ = 0.40 × 0.2/0.60 ≈ 0.133 and thus *τ*_boost_ ≈ 0.083.

This derivation is based on the pairwise secondary attack rate, i.e., the probability that a single susceptible household member is infected by a single infectious index case. In larger households, multiple generations of within-household transmission can produce realized attack rates that exceed this pairwise value. The simulation captures these dynamics, as the transmission model tracks all within-household infections over the course of an outbreak; the pairwise SAR is used here only to calibrate the within-household transmission rate τ.

Next, we calibrated the between-household transmission rate *β* to achieve target *R*_7_ values by running the model at the national level with aggregated ACS household data and systematically varying *β* until the final size matched theoretical predictions for the desired *R*_7_. For an SIR model, the relationship between *R*_0_ and final size, *R*_∞_, is given implicitly by: *R*_∞_ = 1 − *e*^−*R*)^*^R^*\*. For example, *R*_0_ = 1.5 corresponds to *R*_∞_ ≈ 0.58, and *R*_0_ = 3.0 corresponds to *R*_∞_ ≈ 0.94. We used a bisection search algorithm to find *β*, converging when the simulated final size was within 0.0005 of the theoretical value. Calibration was performed using a single-population simulation at the national level, with national household distributions computed as population-weighted averages of all county-level ACS data. The calibrated *β* values were then used in the regional and county-level simulations. We assumed no difference in *β* between agricultural workers and the general community, so that all of the transmission differences in the model would come from differences in household size and crowding. We note that this calibration defines *R*_0_ operationally via the simple SIR final size relationship, which is an approximation for the household-structured model. To calculate *R_eff_* in the presence of vaccination, we used the same relation as the one used to define *R*_0_: *R_eff_* = −*ln*(1 − *R*_∞_)/*R*_∞_, computed from the simulated community infection final size *R*_∞_. Under baseline vaccination, this gives *R_eff_* ≈ 1.4.

We initialized outbreaks by setting 0.1% of individuals in each sub-population as infectious. To distribute these initial infections across household types, we moved a fraction 0.001 × *n* of households of size *n* from the fully susceptible state (*x* = *n*, *y* = 0, *z* = 0) to the single-infection state (*x* = *n* − 1, *y* = 1, *z* = 0). Because this fraction scales with household size *n*, exactly 0.1% of individuals are initially infected regardless of household size, approximating uniform random seeding of infections across the population.

#### Symptomatic infection and workforce impact

To translate epidemic dynamics into agricultural workforce availability, we computed the proportion of the agricultural workforce experiencing symptoms on each day. We first calculated the number of new daily infections for each day as *i_t_* = *S*(*t* − 1) − *S*(*t*), where *S*(*t*) is the proportion susceptible at time *t*. We assumed symptoms began one day after infection onset and lasted for three days, so that individuals infected on day *t* were symptomatic on days *t* + 1, *t* + 2, and *t* + 3. The proportion of symptomatic individuals on day *t* was then:

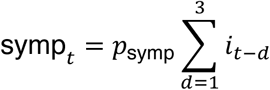

where *p*_symp_ is the probability that an infection is symptomatic. Daily workforce strength was defined as wf(*t*) = 1 − symp*_t_*.

To assess the impact of epidemic timing on crop production, we shifted the epidemic curve so that the community symptomatic peak aligned with each day of the calendar year. For each epidemic curve, we calculated the outbreak-adjusted harvest volume for each crop on each day as *V*_adj_(*t*) = *V*(*t*) × wf(*t*), where *V*(*t*) is the mean harvest volume on day *t*. The total production loss for that epidemic curve, expressed as a percentage, was 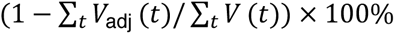. We repeated this calculation for epidemics peaking on each day of the year.

## Supplementary Figures

**Figure S1.**
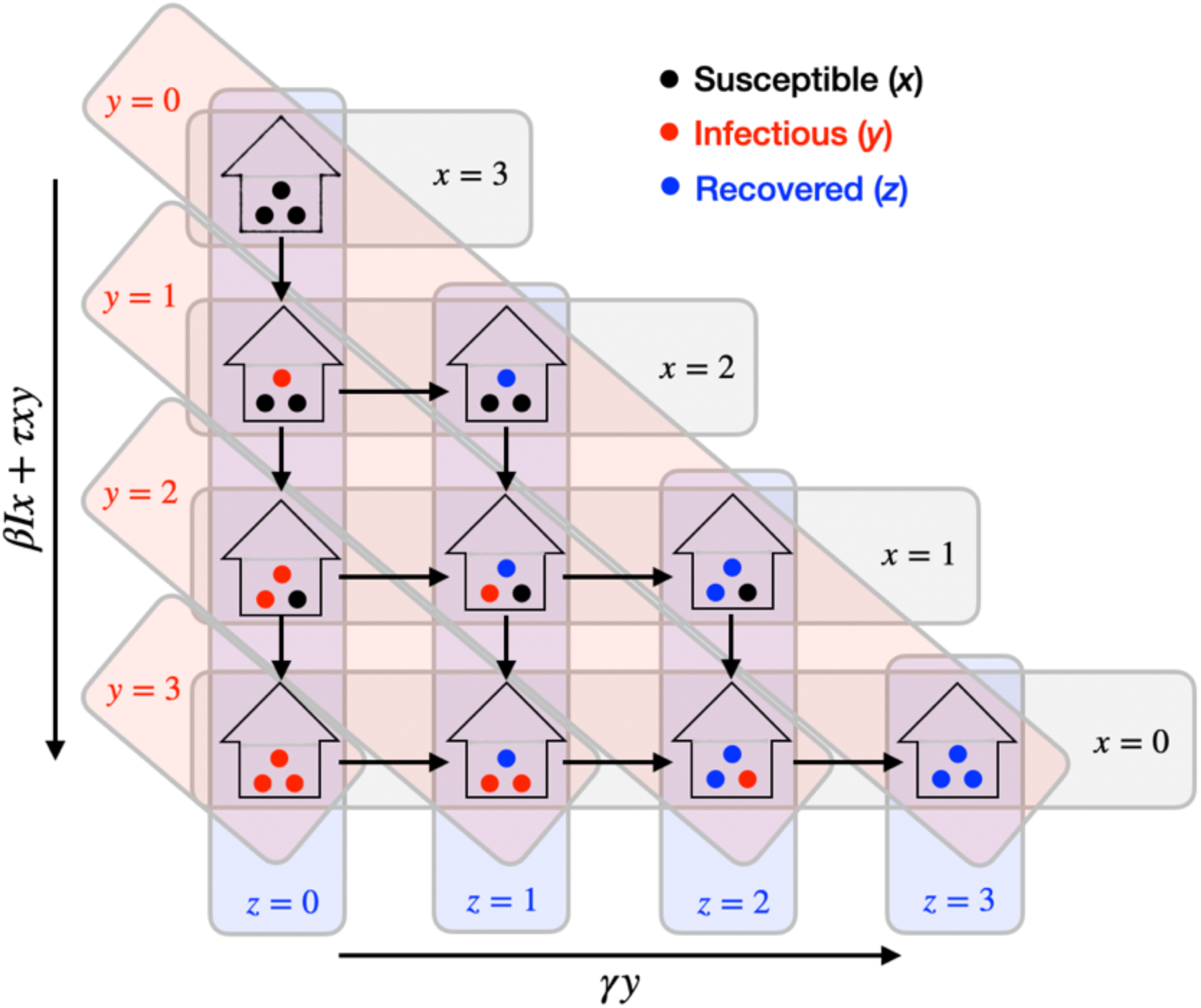
Schematic of the disease transmission model for a household of size 3. An uncrowded agricultural household of size 3 that begins with all members (discs) susceptible (black) is represented as *H_A_*(3,0,0,0) (top-most household). New infections (downward movements; red discs) can occur at rate *βIx* + *τxy*; but since *y* = 0, the force of infection is given fully by the between-household force of infection, *βIx*. Once an infection occurs within the household, either a new household member can become infected at rate *βIx* + *τxy* (downward movement), or the initially infected individual can recover (left-to-right movement; blue discs) at rate *γy*.

**Figure S2.**
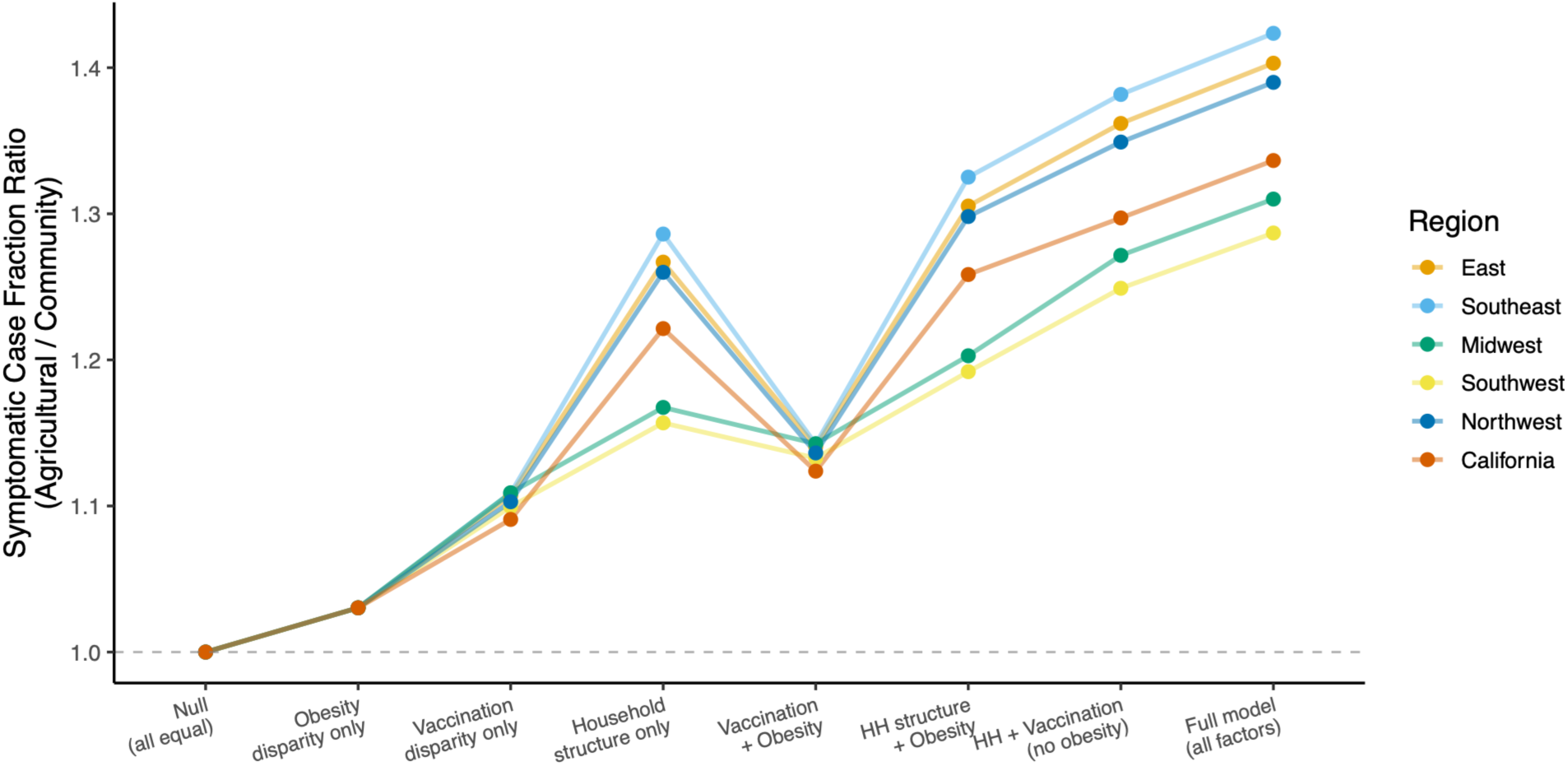
Ratio of total symptomatic cases in the agricultural community *vs.* the general population across model scenarios. Model scenarios (horizontal axis) represent the outcomes of simulations where the labeled factor is allowed to vary between agricultural workers and the general community; all other factors are held constant at the general-community level. The symptomatic case fraction ratio (vertical axis) represents the proportion of total symptomatic cases in agricultural workers *vs*. the general community. Household structure accounts for most of the difference in total cases between the two sub-populations.

**Figure S3.**
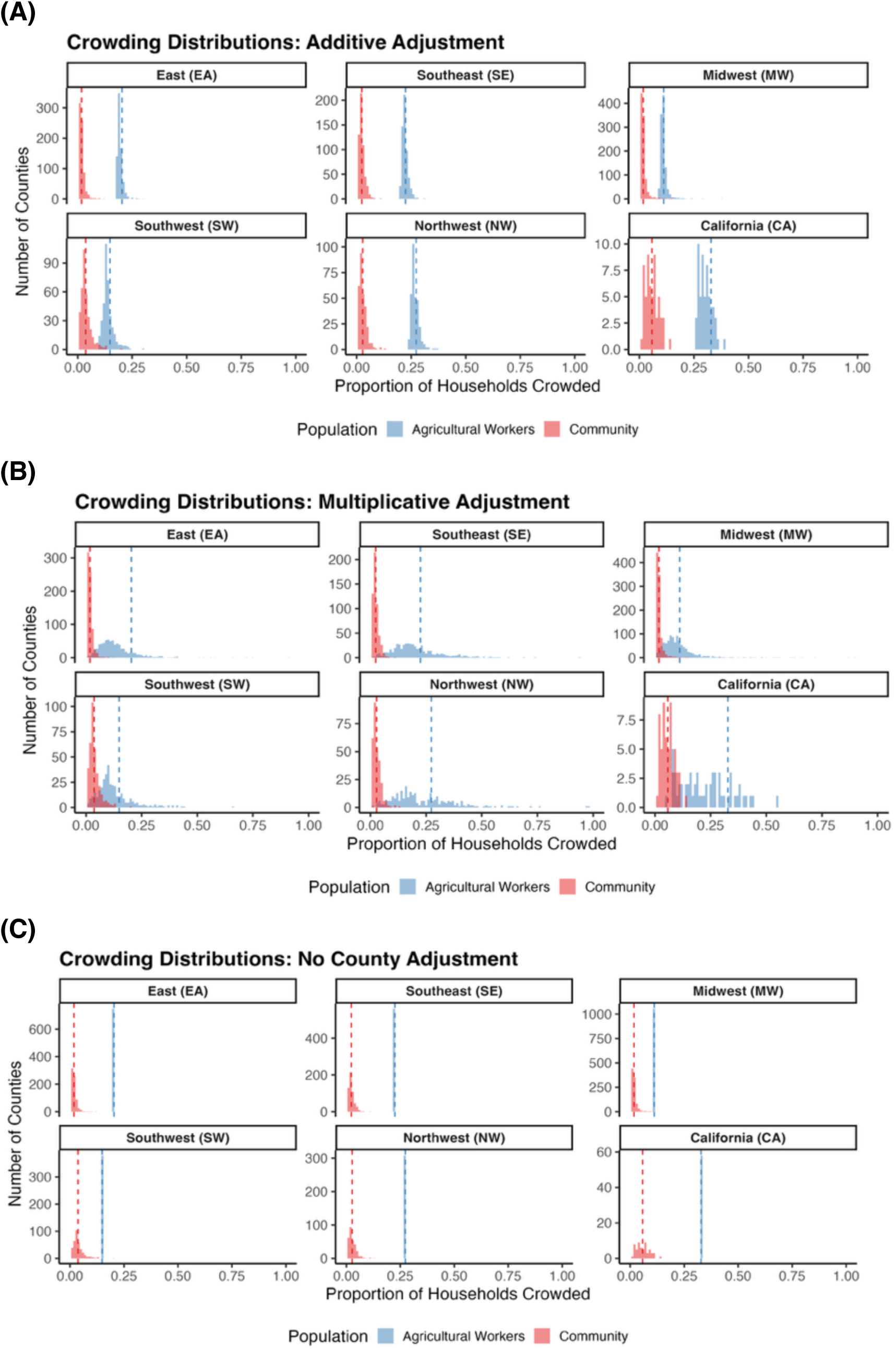
County-level household crowding distributions for agricultural workers and the general community under three imputation methods. Histograms depict the proportion of households crowded in counties within each of the six NAWS regions, for agricultural workers (blue) and the general community (red). County-level crowding rates for the general community are taken from the ACS. County-level crowding rates for agricultural workers are imputed using three different methods: (A) an additive adjustment, where county-level crowding rates for agricultural workers are shifted by the difference between county-level and regional mean ACS values; (B) a multiplicative adjustment, where county-level crowding rates for agricultural workers are shifted by the ratio between county-level and regional mean ACS values; and (C) no adjustment, where crowding rates across counties for agricultural workers are equal to the regional mean. Red dashed vertical lines indicate the regional ACS mean, and blue dashed vertical lines indicate the regional NAWS estimate for agricultural workers.

**Figure S4.**
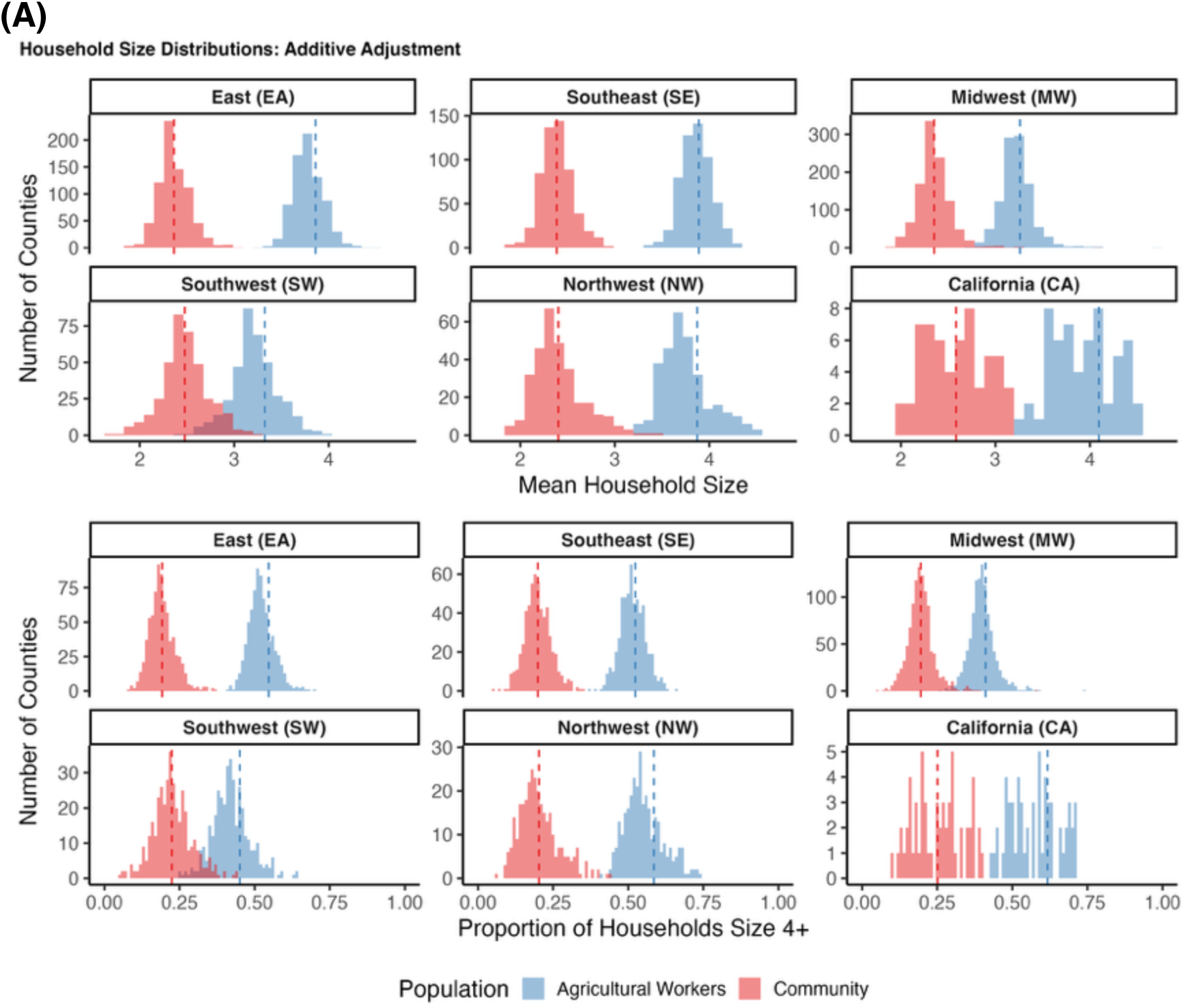
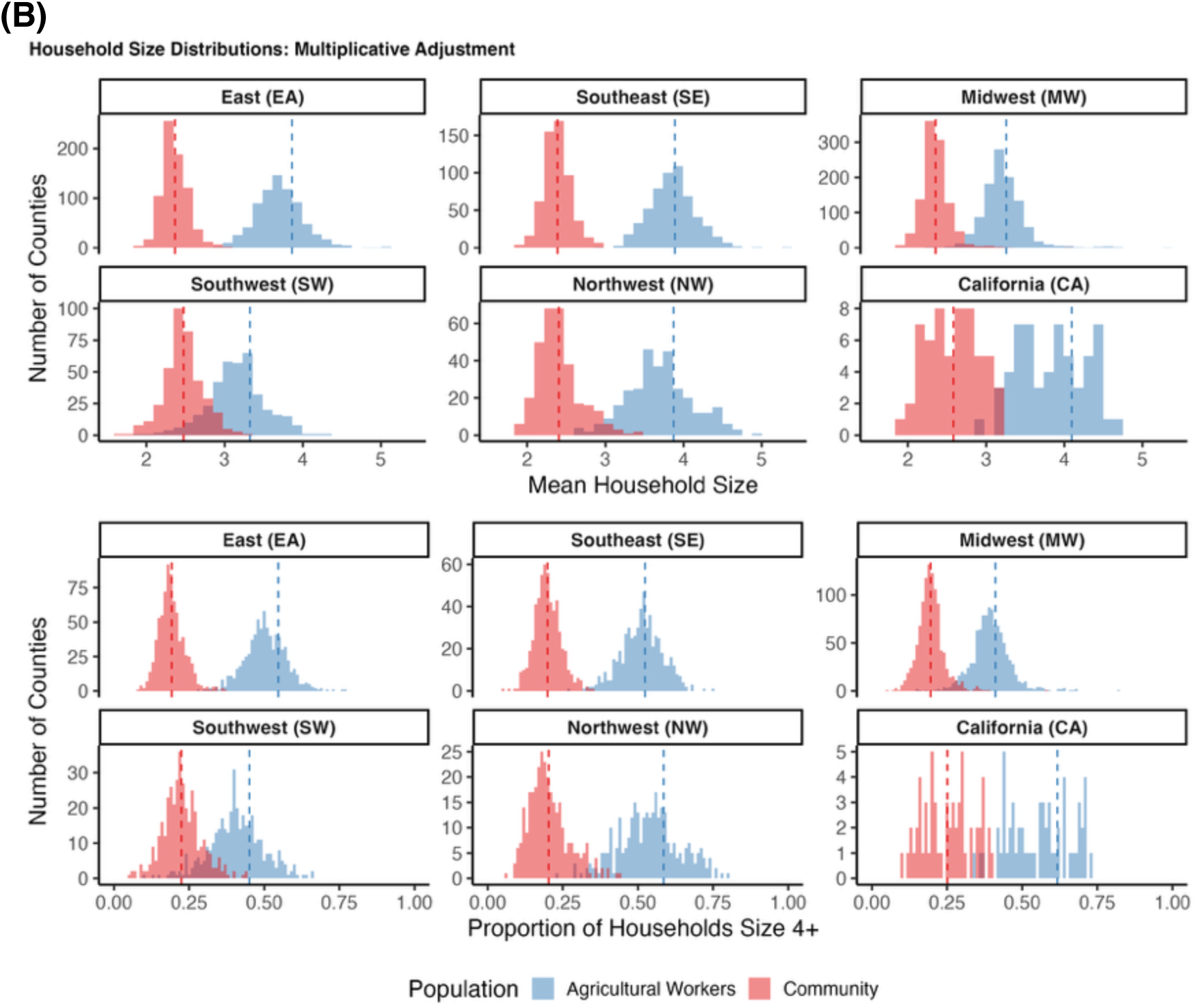
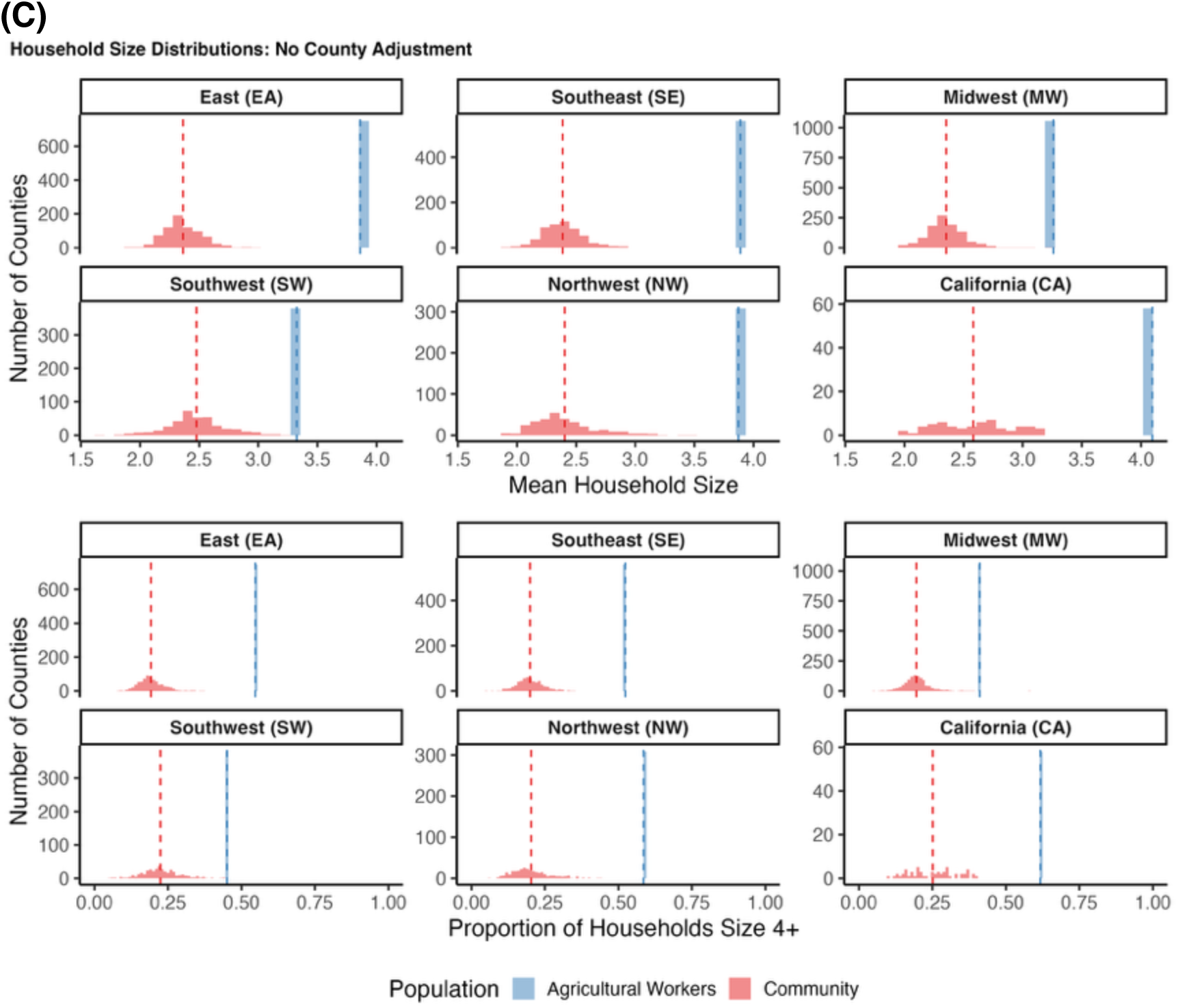
County-level household size distributions for agricultural workers and the general community under three imputation methods. Histograms depict the mean household size (A, C, E) and the proportion of households with 4 or more occupants (B, D, F) in counties within each of the six NAWS regions, for agricultural workers (blue) and the general community (red). County-level household size distributions are taken from the ACS. County-level household size distributions for agricultural workers are imputed using three different methods: (A) an additive adjustment, where county-level household size distributions for agricultural workers are shifted by the difference between county-level and regional mean ACS values and re-normalized; (B) a multiplicative adjustment, where county-level household size distributions for agricultural workers are shifted by the ratio between county-level and regional mean ACS values and renormalized; and (C) no adjustment, where household size distributions across counties for agricultural workers are equal to the regional mean. Red dashed vertical lines indicate the regional ACS mean, and blue dashed vertical lines indicate the regional NAWS estimate for agricultural workers.

**Figure S5.**
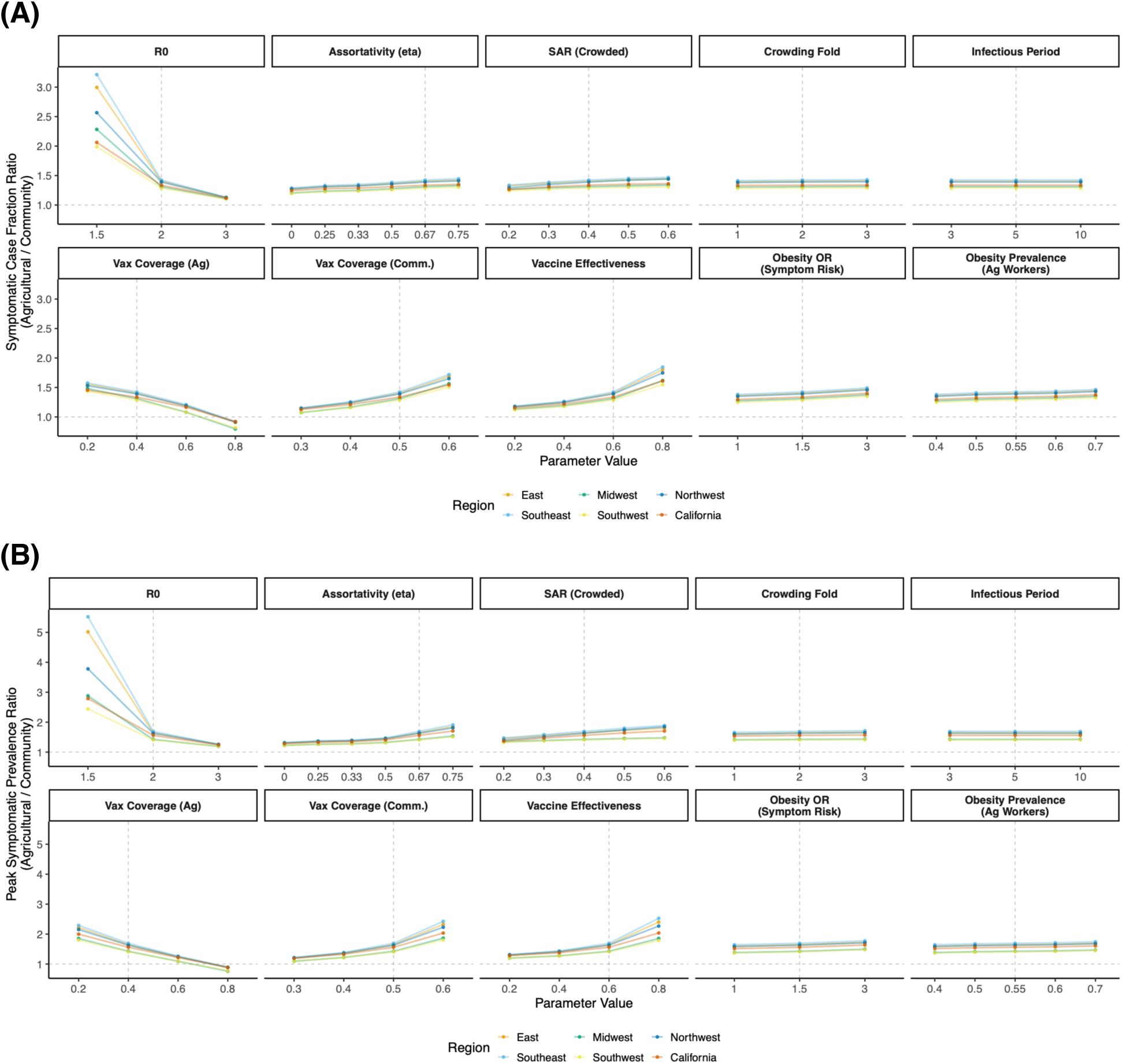
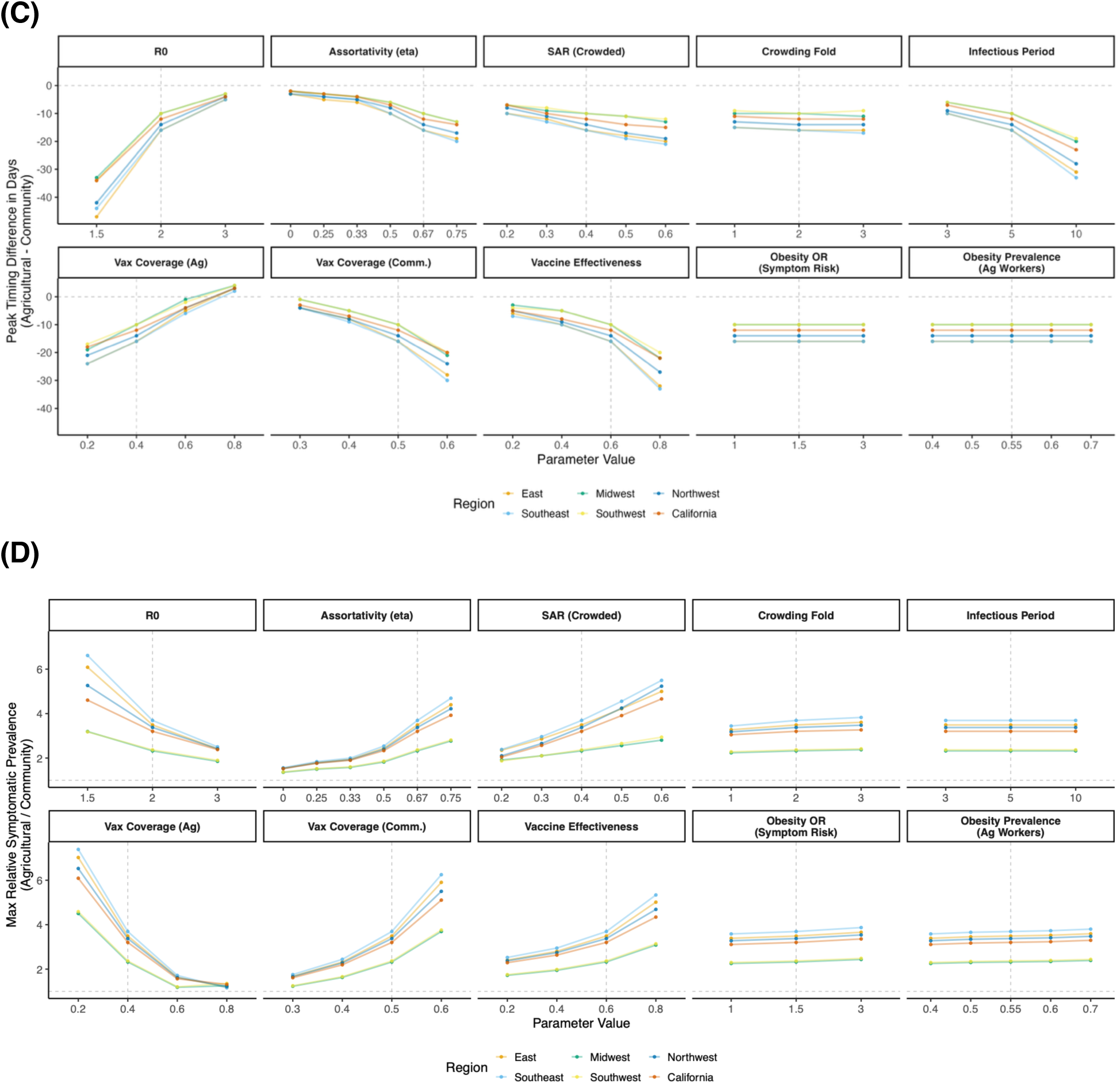
Sensitivity of epidemic summary statistics to key parameters. Panels depict the impact of the basic reproduction number (*R*_0_), assortativity (*η*), secondary attack rate (SAR) in crowded households, fold-difference in crowding rates between households of size 2 and households of size 7+ (*d*), infectious period (1/γ), vaccination coverage in agricultural (Ag) workers, vaccination coverage in the general community (Comm.), vaccine effectiveness, odds ratio of developing symptoms when infected for individuals with *vs.* without obesity, and the prevalence of obesity in agricultural (Ag) workers, on (A) the ratio of final sizes, (B) the ratio of peak sizes, (C) the time difference between peaks, and (D) the maximum prevalence ratio between agricultural workers and the general community. All parameter values are held at their baseline values (**Supplementary Table S2**) except for the one being varied in the panel. Colors represent the various NAWS regions. Dashed horizontal lines mark the value indicating “no difference”. Dashed vertical lines mark the baseline values used in the main analysis.

**Figure S6.**
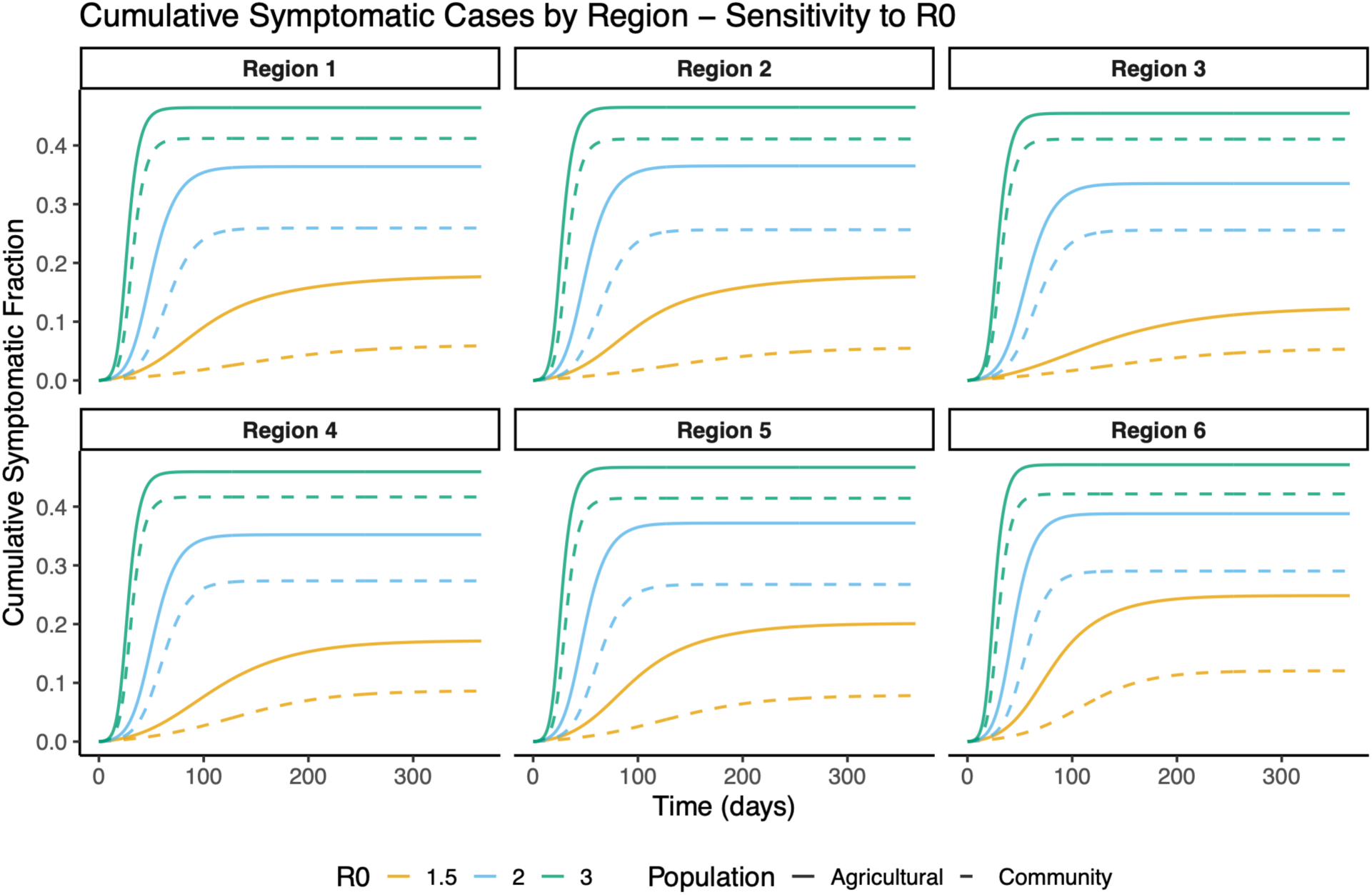
Impact of the basic reproduction number (*R*_0_) on cumulative cases among agricultural workers and the general community. Panels depict the simulated cumulative cases over time for agricultural workers (solid lines) and the general community (dashed lines) in each of the six NAWS regions across different *R*_0_ values (colors). All other parameter values are held at baseline (**Supplementary Table S2**).

**Figure S7.**
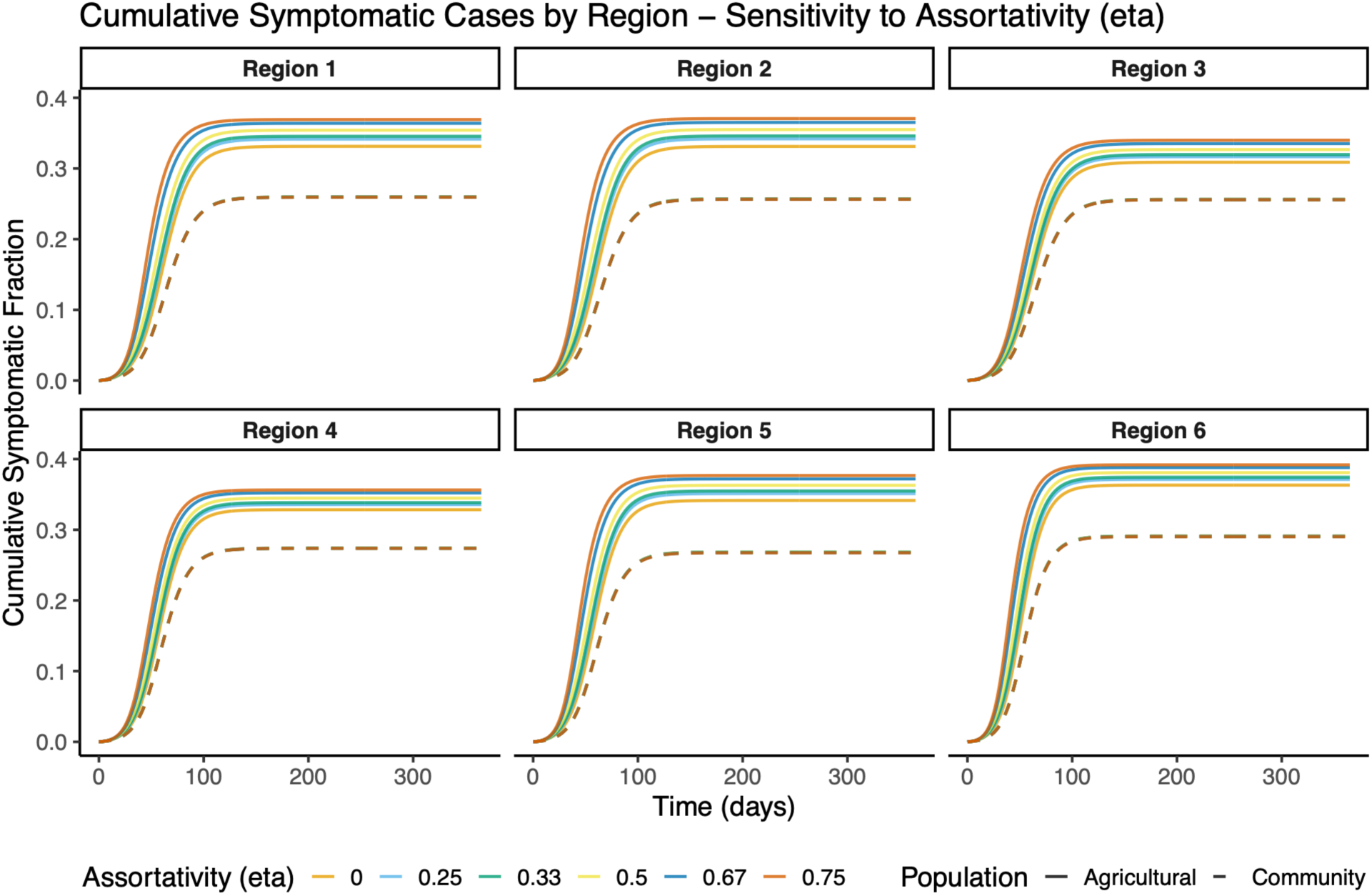
Impact of assortativity (*η*) on cumulative cases among agricultural workers and the general community. Panels depict the simulated cumulative cases over time for agricultural workers (solid lines) and the general community (dashed lines) in each of the six NAWS regions across different values of the assortativity parameter *η* (colors), where larger *η* corresponds to more within-group mixing. All other parameter values are held at baseline (**Supplementary Table S2**).

**Figure S8.**
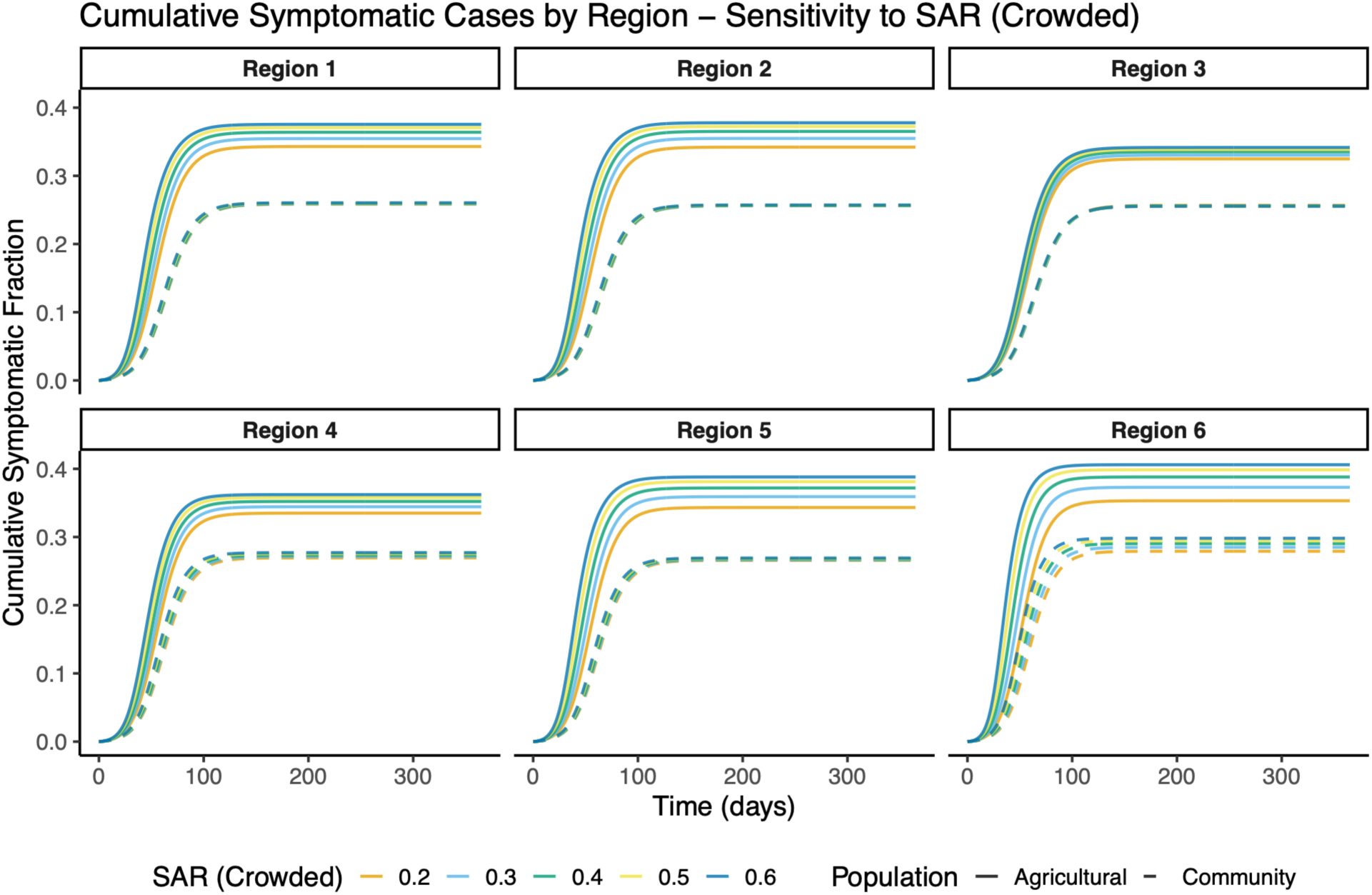
Impact of the secondary attack rate (SAR) in crowded households on cumulative cases among agricultural workers and the general community. Panels depict the simulated cumulative cases over time for agricultural workers (solid lines) and the general community (dashed lines) in each of the six NAWS regions across different values of the secondary attack rate (SAR) in crowded households. All other parameter values are held at baseline; note that the SAR in uncrowded households was held fixed at 0.2. (**Supplementary Table S2**).

**Figure S9.**
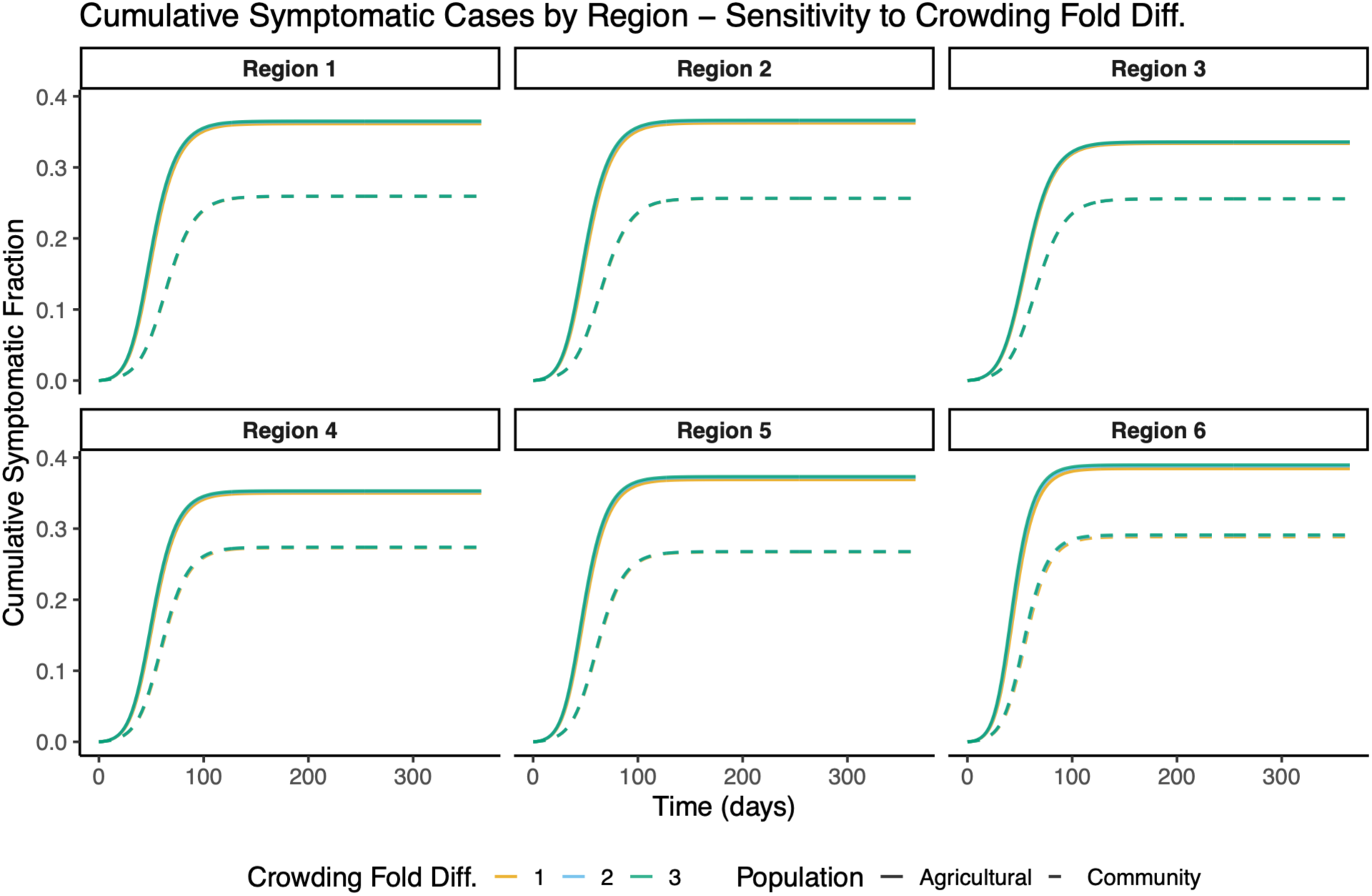
Impact of the crowding fold difference parameter *d* on cumulative cases among agricultural workers and the general community. Panels depict the simulated cumulative cases over time for agricultural workers (solid lines) and the general community (dashed lines) in each of the six NAWS regions across different values of the crowding fold-difference parameter *d* (colors), which represents how much more likely a household of size 7+ is to be crowded than a household of size 2. All other parameter values are held at baseline (**Supplementary Table S2**).

**Figure S10.**
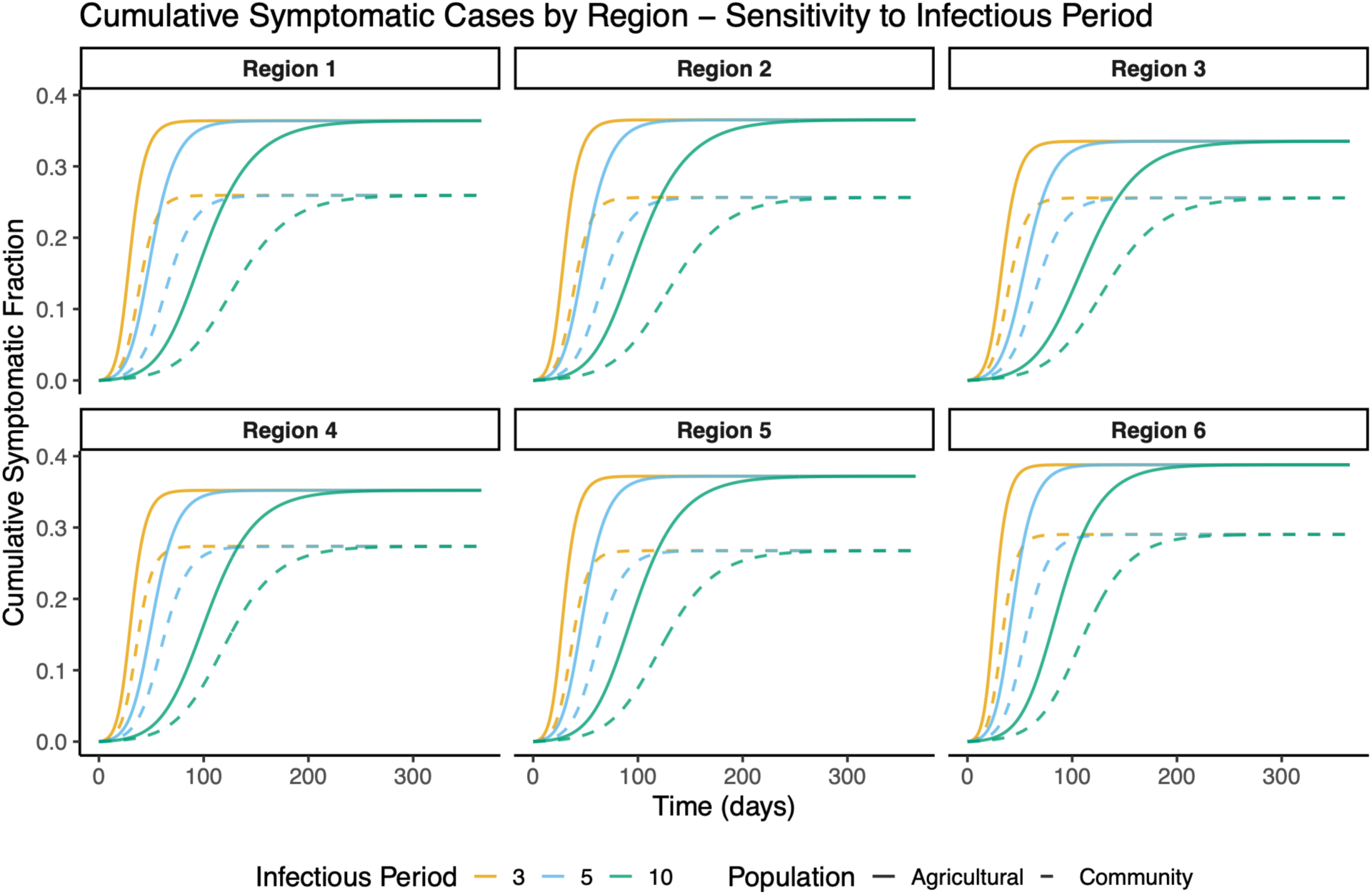
Impact of the infectious period 1/γ on cumulative cases among agricultural workers and the general community. Panels depict the simulated cumulative cases over time for agricultural workers (solid lines) and the general community (dashed lines) in each of the six NAWS regions across different values of the infections period 1/γ (colors). All other parameter values are held at baseline (**Supplementary Table S2**).

**Figure S11.**
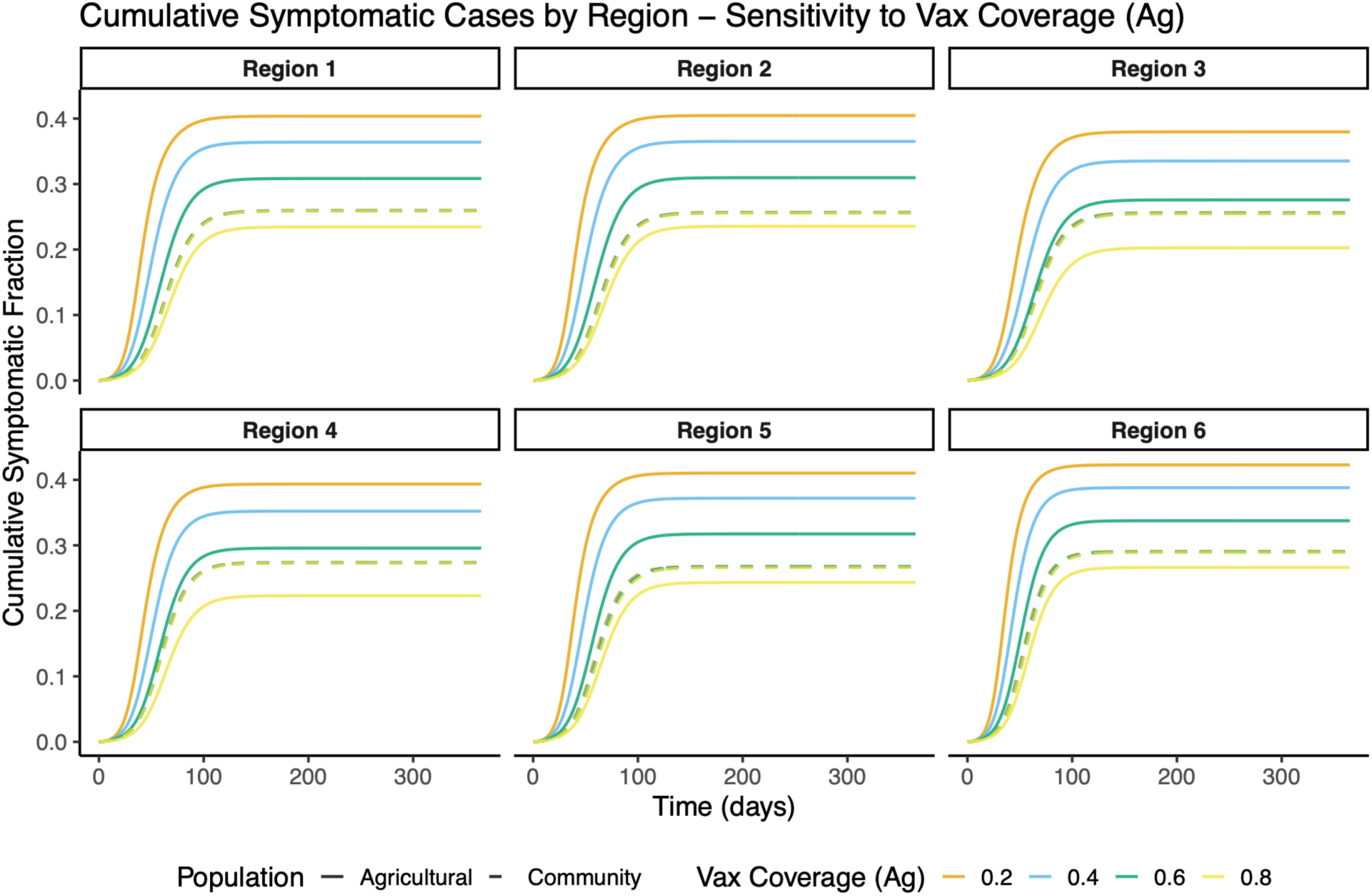
Impact of the agricultural-worker vaccination coverage on cumulative cases among agricultural workers and the general community. Panels depict the simulated cumulative cases over time for agricultural workers (solid lines) and the general community (dashed lines) in each of the six NAWS regions across different levels of vaccination coverage. All other parameter values are held at baseline (**Supplementary Table S2**).

**Figure S12.**
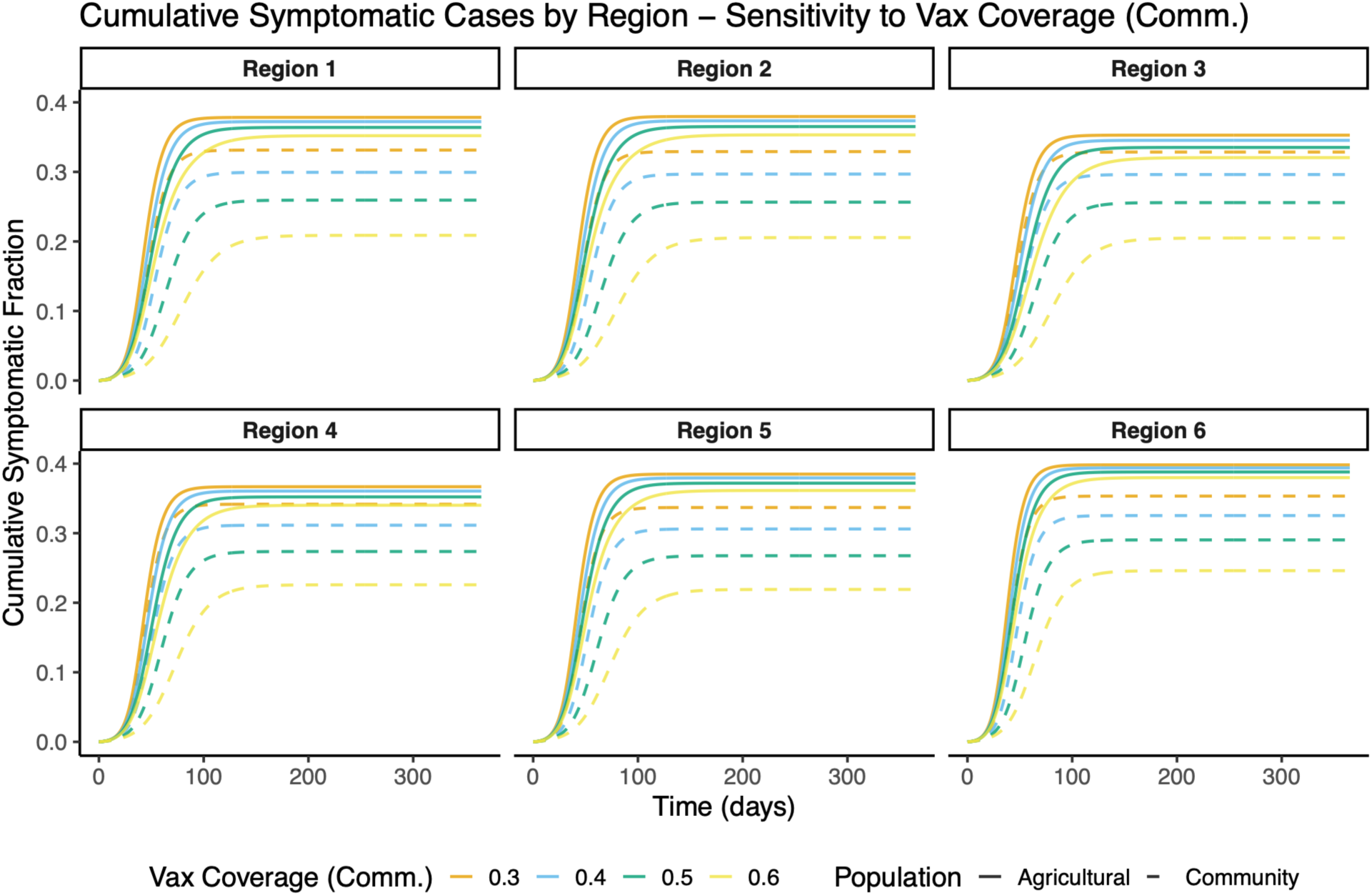
Impact of the general-community vaccination coverage on cumulative cases among agricultural workers and the general community. Panels depict the simulated cumulative cases over time for agricultural workers (solid lines) and the general community (dashed lines) in each of the six NAWS regions across different levels of vaccination coverage. All other parameter values are held at baseline (**Supplementary Table S2**).

**Figure S13.**
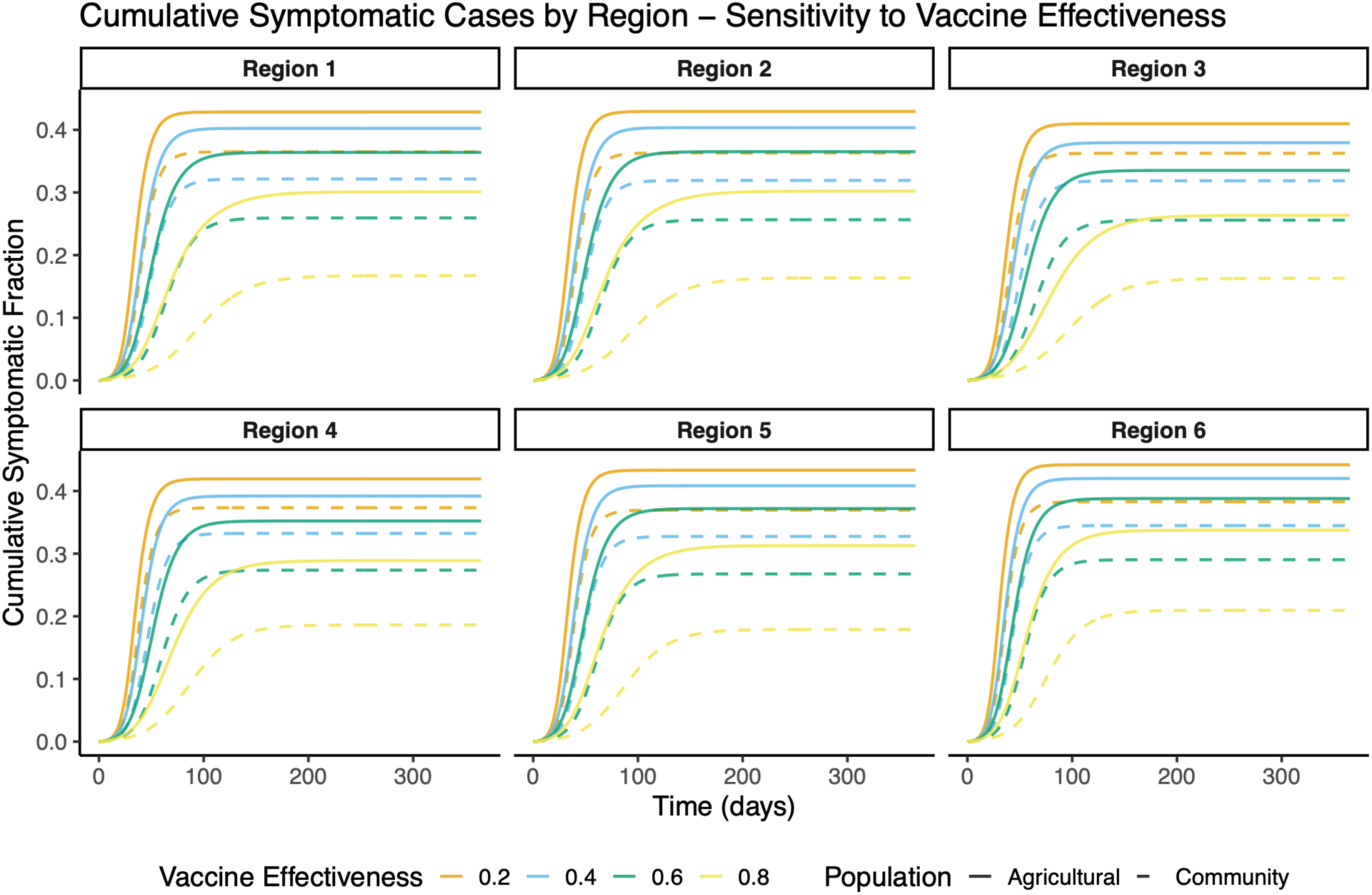
Impact of the vaccine effectiveness on cumulative cases among agricultural workers and the general community. Panels depict the simulated cumulative cases over time for agricultural workers (solid lines) and the general community (dashed lines) in each of the six NAWS regions across different values of vaccine effectiveness (colors). All other parameter values are held at baseline (**Supplementary Table S2**).

**Figure S14.**
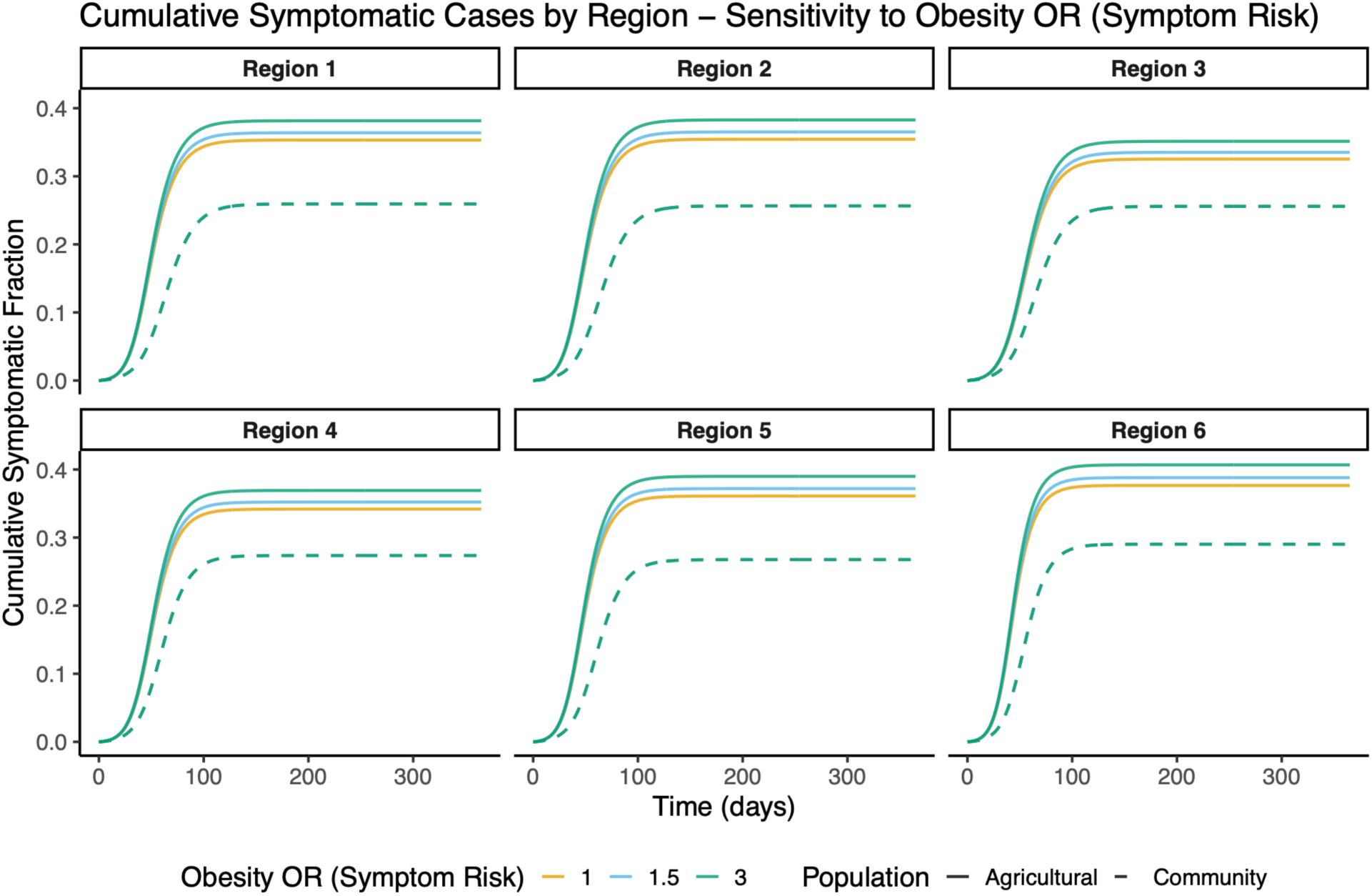
Impact of the odds ratio of developing symptoms given infection when obese on cumulative cases among agricultural workers and the general community. Panels depict the simulated cumulative cases over time for agricultural workers (solid lines) and the general community (dashed lines) in each of the six NAWS regions across different values of the odds ratio of developing symptoms, given infection, when obese *vs.* when not obese (colors). All other parameter values are held at baseline (**Supplementary Table S2**).

**Figure S15.**
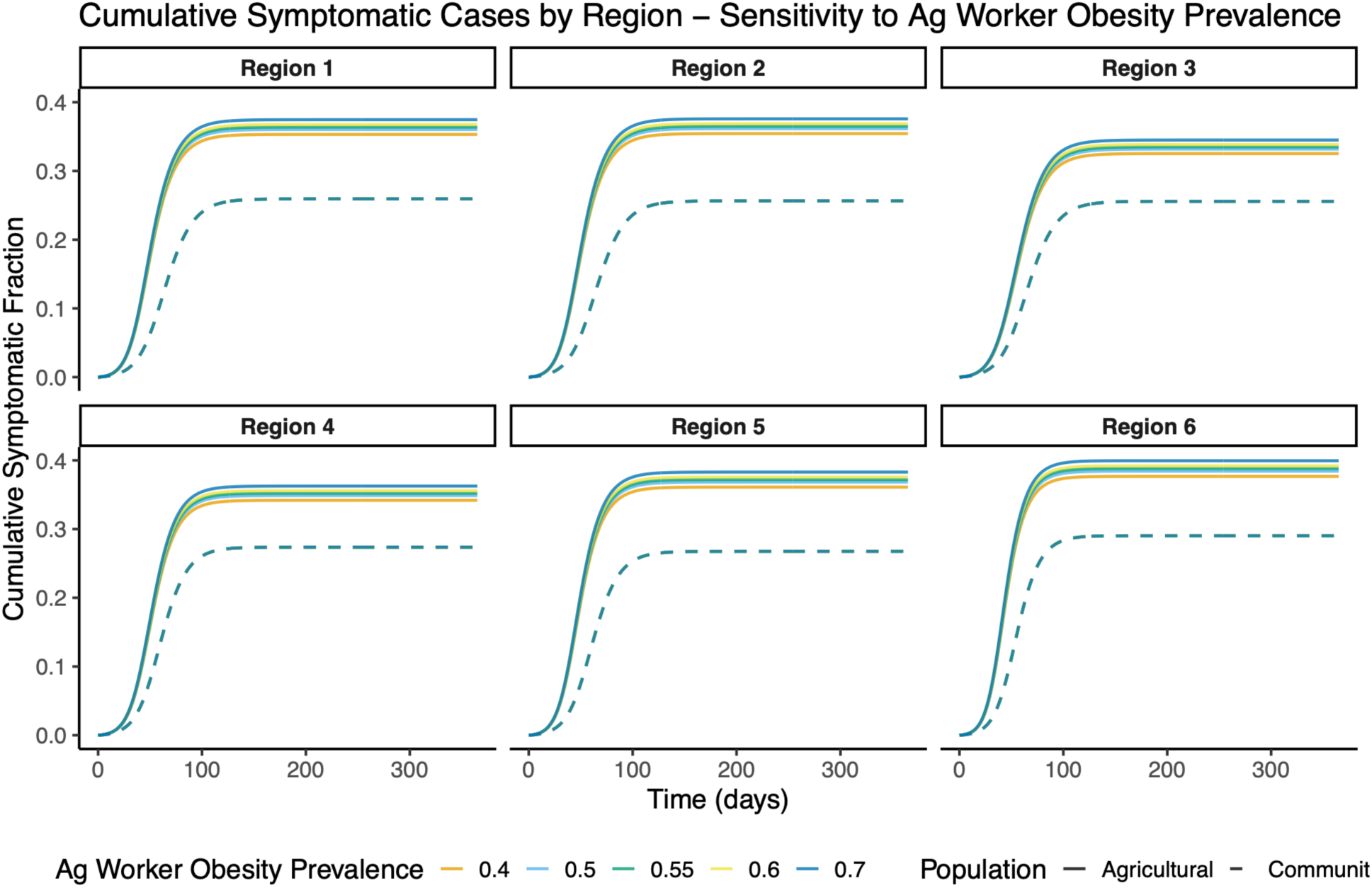
Impact of obesity prevalence among agricultural workers on cumulative cases among agricultural workers and the general community. Panels depict the simulated cumulative cases over time for agricultural workers (solid lines) and the general community (dashed lines) in each of the six NAWS regions across different values of obesity prevalence (colors). All other parameter values are held at baseline (**Supplementary Table S2**).

**Figure S16.**
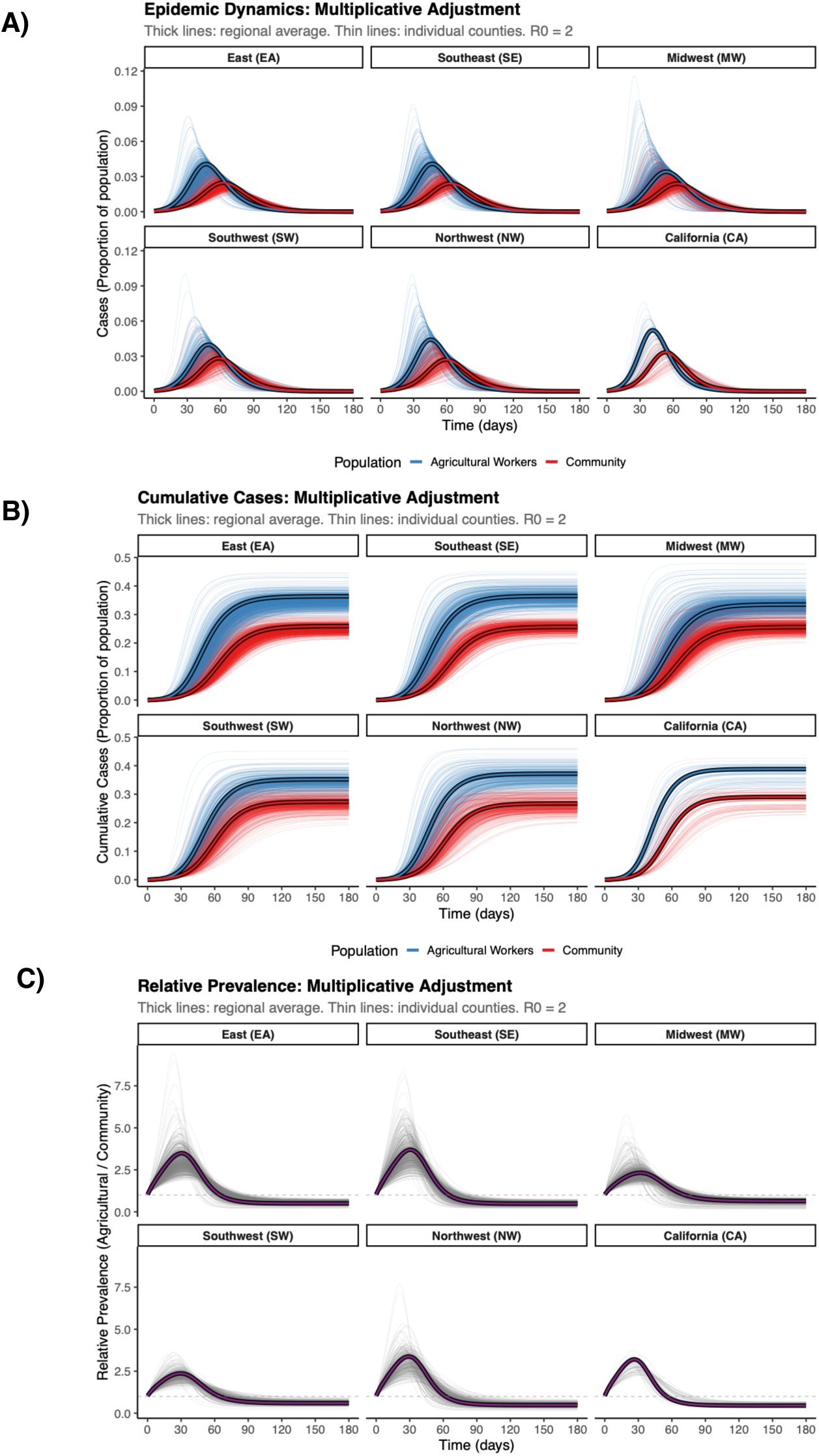
Epidemic trajectories under the multiplicative county-level imputation method for agricultural worker household characteristics. (A) Simulated infection prevalence over time for agricultural workers (blue) and the general community (red) for the six NAWS regions, with simulations at both the region level (thick lines with borders) and county level (thin, semi-transparent lines). (B) Cumulative infections over time for agricultural workers (blue) and the general community (red) for the six NAWS regions, with simulations at both the region level (thick lines with borders) and county level (thin, semi-transparent lines). (C) Prevalence ratio between agricultural workers and the general community for the six NAWS regions, with simulations at both the region level (thick lines with borders) and county level (thin, semi-transparent lines). County-level household attributes for agricultural workers are imputed using the “multiplicative” method, in which regional NAWS household attributes are adjusted by the ratio between the county-level ACS values and the regional ACS mean.

**Figure S17.**
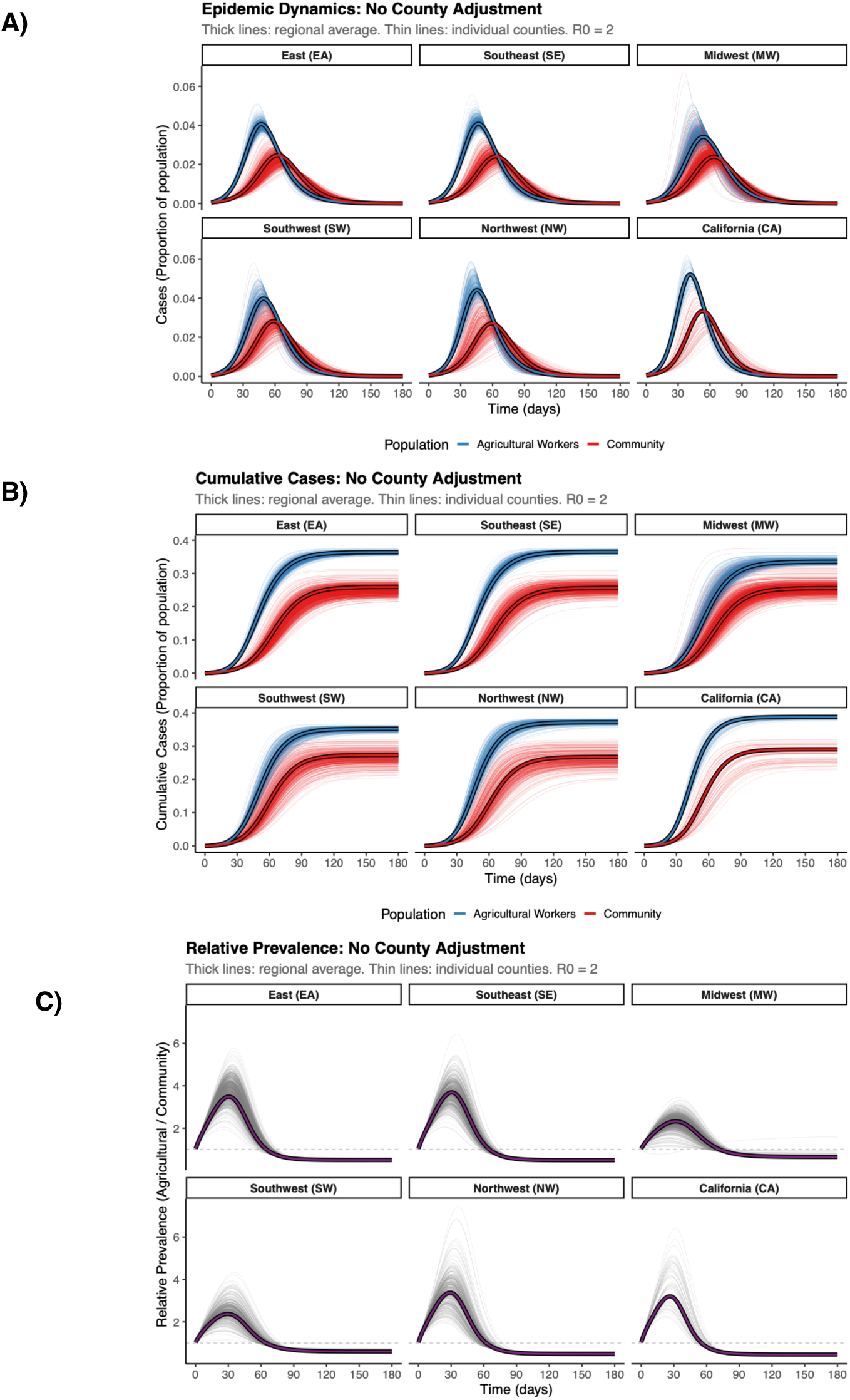
Epidemic trajectories under the “null” county-level imputation method for agricultural worker household characteristics. (A) Simulated infection prevalence over time for agricultural workers (blue) and the general community (red) for the six NAWS regions, with simulations at both the region level (thick lines with borders) and county level (thin, semi-transparent lines). (B) Cumulative infections over time for agricultural workers (blue) and the general community (red) for the six NAWS regions, with simulations at both the region level (thick lines with borders) and county level (thin, semi-transparent lines). (C) Prevalence ratio between agricultural workers and the general community for the six NAWS regions, with simulations at both the region level (thick lines with borders) and county level (thin, semi-transparent lines). County-level household attributes for agricultural workers are imputed using the “null” method, in which county-level household attributes for agricultural workers are taken to be equal to the regional NAWS value, with no adjustment.

**Figure S18.**
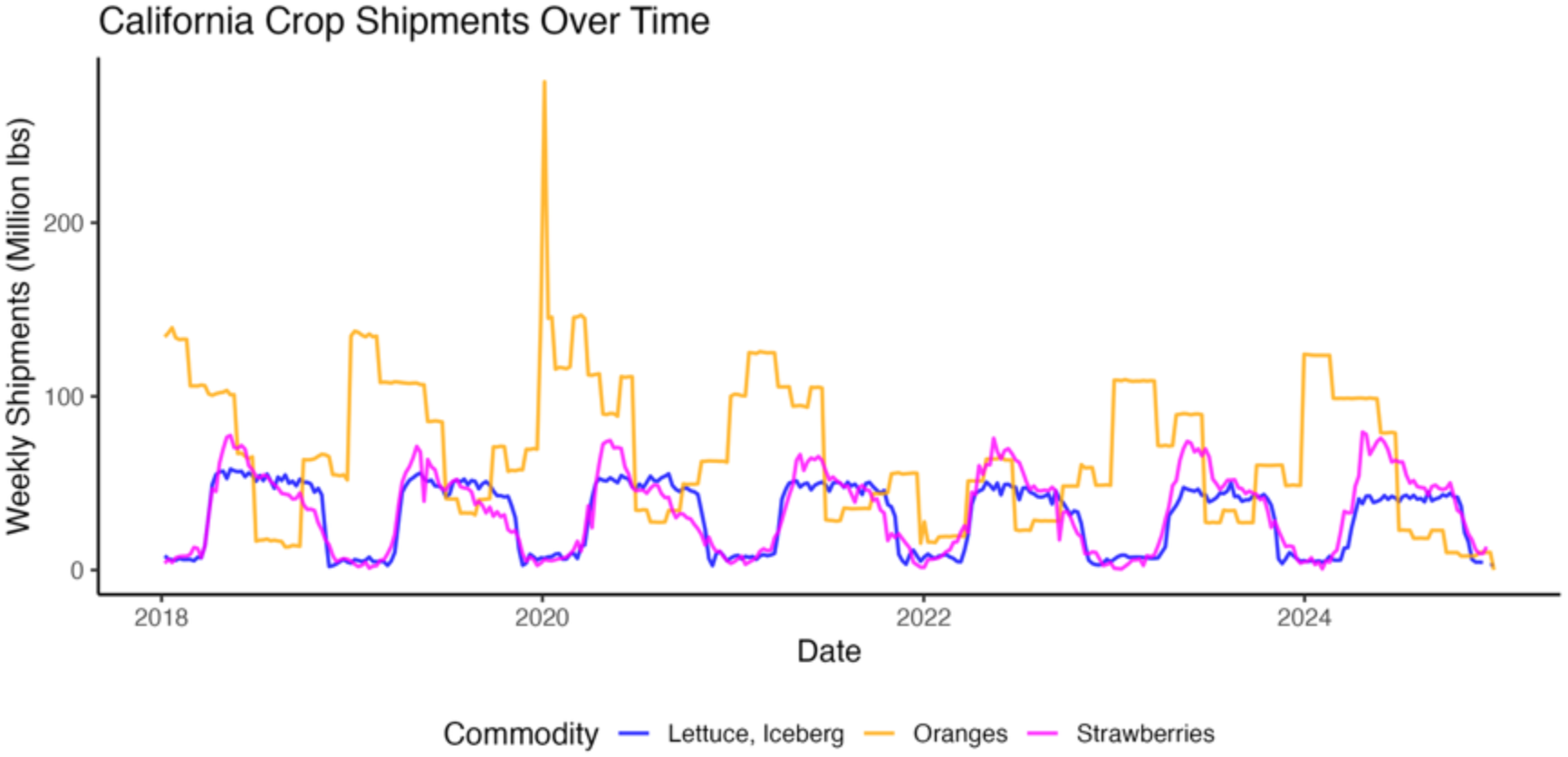
Weekly crop movements for three labor-intensive California crops Weekly point-to-point crop shipments (in million pounds) originating in California for iceberg lettuce (blue), oranges (orange), and strawberries (magenta), from 2018–2024.

**Figure S19.**
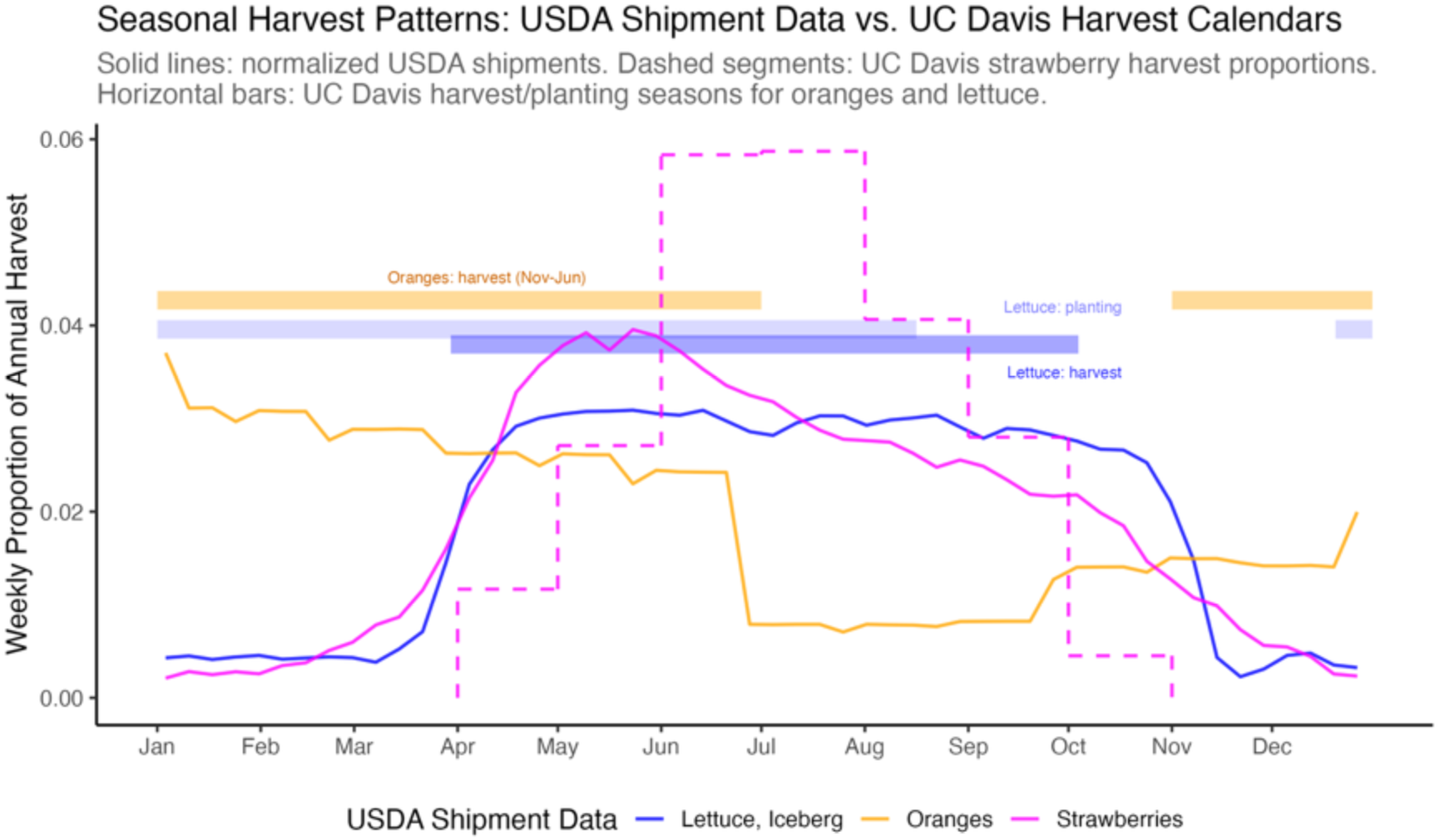
Average weekly crop movements with known harvesting patterns. Comparison of normalized average weekly crop movements (proportion of total annual volume from 2018-2024; solid lines) with harvest information from University of California Agriculture and Natural Resources Cooperative Extension reports (dashed line, semi-transparent bars) for iceberg lettuce (blue), oranges (orange), and strawberries (magenta) in California. For strawberries, the University of California report gives explicit monthly harvest proportions, which are re-scaled to approximate weekly harvest volumes (dashed magenta line). For oranges, the reported harvest season runs from November to June (semi-transparent orange bar). For iceberg lettuce, the planting season runs from late December to mid-August (lighter semi-transparent blue bar). We computed an approximate harvest season (darker semi-transparent blue bar) by shifting the planting window forward by 100 days at the cool-season (December) end and by 50 days at the warm-season end (August) to account for reported seasonal differences in maturation time.

**Figure S20.**
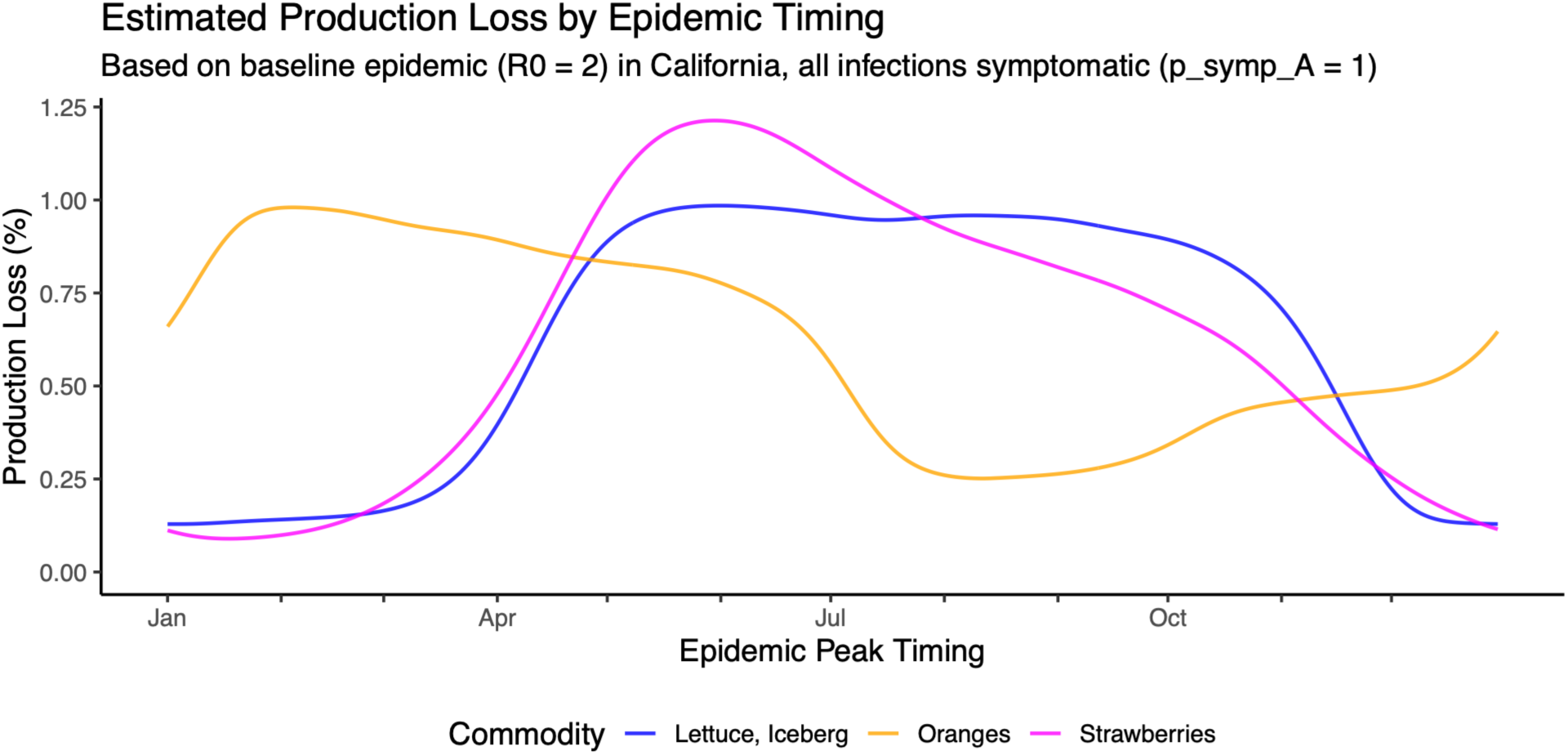
Estimated crop production loss as a function of epidemic peak timing when all infections are symptomatic. Simulated percent of total harvest volume impacted by outbreak-induced labor shortages for iceberg lettuce (blue), strawberries (magenta), and oranges (orange) under baseline parameter values. The horizontal axis represents the day of the year on which infection prevalence peaks in the general community (peak infections among agricultural workers occur a few days earlier).

## Supplementary Tables

**Table S1.** Household characteristics by region for agricultural workers and the general community. Mean household size is the population-weighted average across household sizes 1–7+. Crowding proportion is the fraction of households with more than 1 occupant per room. Agricultural worker data are from the National Agricultural Workers Survey (NAWS), and general community data are from the American Community Survey (ACS), aggregated to the regional level using population-weighted averages.

| Region | Mean household size |  | Prop. of households size 4+ |  | Crowding proportion |  |
| --- | --- | --- | --- | --- | --- | --- |
|  | Agricultural workers | General community | Agricultural workers | General community | Agricultural workers | General community |
| East | 3.9 | 2.4 | 54.7% | 21.7% | 20.3% | 2.8% |
| Southeast | 3.9 | 2.4 | 52.3% | 20.6% | 22.4% | 2.6% |
| Midwest | 3.3 | 2.4 | 41.1% | 20.7% | 11.2% | 1.9% |
| Southwest | 3.3 | 2.6 | 45.1% | 25.5% | 14.8% | 4.5% |
| Northwest | 3.9 | 2.5 | 58.6% | 23.4% | 27.3% | 3.2% |
| California | 4.1 | 2.8 | 61.7% | 29.3% | 32.8% | 8.3% |

**Table S2.** Baseline and sensitivity analysis parameter values for the disease transmission model. Each sensitivity analysis varies one parameter at a time while holding all others at baseline values (first row). Bold values indicate the parameter(s) being varied in each row. Columns represent the basic reproduction number (*R*0), assortativity (*η*), secondary attack rate (SAR) in crowded households, fold-difference in crowding probability for large *vs.* small households (d), transmission constant for uncrowded (τ) and crowded (τboost) households, between-household transmission constant (β), recovery rate (γ), vaccine effectiveness (VE), vaccine coverage in agricultural workers (VCA) and the general community (VCC), obesity prevalence among agricultural workers (ObA), odds ratio of developing symptoms while obese given infection (OROb), and the probability of developing symptoms for agricultural workers (psymp,A).

| $R_0$ | $\eta$ | SAR | $d$ | $\tau$ | $\tau_{\text{boost}}$ | $\beta$ | $\gamma$ | VE | $VC_A$ | $VC_C$ | $Ob_A$ | $OR_{Ob}$ | $p_{\text{symp},A}$ |
| --- | --- | --- | --- | --- | --- | --- | --- | --- | --- | --- | --- | --- | --- |
| 2 | 0.67 | 0.4 | 2 | 0.050 | 0.083 | 0.308 | 0.20 | 0.6 | 0.4 | 0.5 | 0.55 | 1.5 | 0.52 |
| **1.5** | 0.67 | 0.4 | 2 | 0.050 | 0.083 | 0.211 | 0.20 | 0.6 | 0.4 | 0.5 | 0.55 | 1.5 | 0.52 |
| **3** | 0.67 | 0.4 | 2 | 0.050 | 0.083 | 0.505 | 0.20 | 0.6 | 0.4 | 0.5 | 0.55 | 1.5 | 0.52 |
| 2 | **0.75** | 0.4 | 2 | 0.050 | 0.083 | 0.308 | 0.20 | 0.6 | 0.4 | 0.5 | 0.55 | 1.5 | 0.52 |
| 2 | **0.50** | 0.4 | 2 | 0.050 | 0.083 | 0.308 | 0.20 | 0.6 | 0.4 | 0.5 | 0.55 | 1.5 | 0.52 |
| 2 | **0.33** | 0.4 | 2 | 0.050 | 0.083 | 0.308 | 0.20 | 0.6 | 0.4 | 0.5 | 0.55 | 1.5 | 0.52 |
| 2 | **0.25** | 0.4 | 2 | 0.050 | 0.083 | 0.308 | 0.20 | 0.6 | 0.4 | 0.5 | 0.55 | 1.5 | 0.52 |
| 2 | **0.00** | 0.4 | 2 | 0.050 | 0.083 | 0.308 | 0.20 | 0.6 | 0.4 | 0.5 | 0.55 | 1.5 | 0.52 |
| 2 | 0.67 | **0.2** | 2 | 0.050 | 0.000 | 0.311 | 0.20 | 0.6 | 0.4 | 0.5 | 0.55 | 1.5 | 0.52 |
| 2 | 0.67 | **0.3** | 2 | 0.050 | 0.036 | 0.309 | 0.20 | 0.6 | 0.4 | 0.5 | 0.55 | 1.5 | 0.52 |
| 2 | 0.67 | **0.5** | 2 | 0.050 | 0.150 | 0.306 | 0.20 | 0.6 | 0.4 | 0.5 | 0.55 | 1.5 | 0.52 |
| 2 | 0.67 | **0.6** | 2 | 0.050 | 0.250 | 0.305 | 0.20 | 0.6 | 0.4 | 0.5 | 0.55 | 1.5 | 0.52 |
| 2 | 0.67 | 0.4 | **1** | 0.050 | 0.083 | 0.308 | 0.20 | 0.6 | 0.4 | 0.5 | 0.55 | 1.5 | 0.52 |
| 2 | 0.67 | 0.4 | **3** | 0.050 | 0.083 | 0.307 | 0.20 | 0.6 | 0.4 | 0.5 | 0.55 | 1.5 | 0.52 |
| 2 | 0.67 | 0.4 | 2 | 0.083 | 0.139 | 0.513 | **0.33** | 0.6 | 0.4 | 0.5 | 0.55 | 1.5 | 0.52 |
| 2 | 0.67 | 0.4 | 2 | 0.025 | 0.042 | 0.154 | **0.10** | 0.6 | 0.4 | 0.5 | 0.55 | 1.5 | 0.52 |
| 2 | 0.67 | 0.4 | 2 | 0.050 | 0.083 | 0.308 | 0.20 | 0.6 | **0.2** | 0.5 | 0.55 | 1.5 | 0.52 |
| 2 | 0.67 | 0.4 | 2 | 0.050 | 0.083 | 0.308 | 0.20 | 0.6 | **0.6** | 0.5 | 0.55 | 1.5 | 0.52 |
| 2 | 0.67 | 0.4 | 2 | 0.050 | 0.083 | 0.308 | 0.20 | 0.6 | **0.8** | 0.5 | 0.55 | 1.5 | 0.52 |
| 2 | 0.67 | 0.4 | 2 | 0.050 | 0.083 | 0.308 | 0.20 | 0.6 | 0.4 | **0.3** | 0.55 | 1.5 | 0.52 |
| 2 | 0.67 | 0.4 | 2 | 0.050 | 0.083 | 0.308 | 0.20 | 0.6 | 0.4 | **0.4** | 0.55 | 1.5 | 0.52 |
| 2 | 0.67 | 0.4 | 2 | 0.050 | 0.083 | 0.308 | 0.20 | 0.6 | 0.4 | **0.6** | 0.55 | 1.5 | 0.52 |
| 2 | 0.67 | 0.4 | 2 | 0.050 | 0.083 | 0.308 | 0.20 | **0.2** | 0.4 | 0.5 | 0.55 | 1.5 | 0.52 |
| 2 | 0.67 | 0.4 | 2 | 0.050 | 0.083 | 0.308 | 0.20 | **0.4** | 0.4 | 0.5 | 0.55 | 1.5 | 0.52 |
| 2 | 0.67 | 0.4 | 2 | 0.050 | 0.083 | 0.308 | 0.20 | **0.8** | 0.4 | 0.5 | 0.55 | 1.5 | 0.52 |
| 2 | 0.67 | 0.4 | 2 | 0.050 | 0.083 | 0.308 | 0.20 | 0.6 | 0.4 | 0.5 | 0.55 | **1** | 0.50 |
| 2 | 0.67 | 0.4 | 2 | 0.050 | 0.083 | 0.308 | 0.20 | 0.6 | 0.4 | 0.5 | 0.55 | **1.5** | 0.52 |
| 2 | 0.67 | 0.4 | 2 | 0.050 | 0.083 | 0.308 | 0.20 | 0.6 | 0.4 | 0.5 | 0.55 | **3** | 0.54 |
| 2 | 0.67 | 0.4 | 2 | 0.050 | 0.083 | 0.308 | 0.20 | 0.6 | 0.4 | 0.5 | **0.4** | 1.5 | 0.50 |
| 2 | 0.67 | 0.4 | 2 | 0.050 | 0.083 | 0.308 | 0.20 | 0.6 | 0.4 | 0.5 | **0.5** | 1.5 | 0.51 |
| 2 | 0.67 | 0.4 | 2 | 0.050 | 0.083 | 0.308 | 0.20 | 0.6 | 0.4 | 0.5 | **0.55** | 1.5 | 0.52 |
| 2 | 0.67 | 0.4 | 2 | 0.050 | 0.083 | 0.308 | 0.20 | 0.6 | 0.4 | 0.5 | **0.6** | 1.5 | 0.52 |
| 2 | 0.67 | 0.4 | 2 | 0.050 | 0.083 | 0.308 | 0.20 | 0.6 | 0.4 | 0.5 | **0.7** | 1.5 | 0.53 |

**Table S3.**
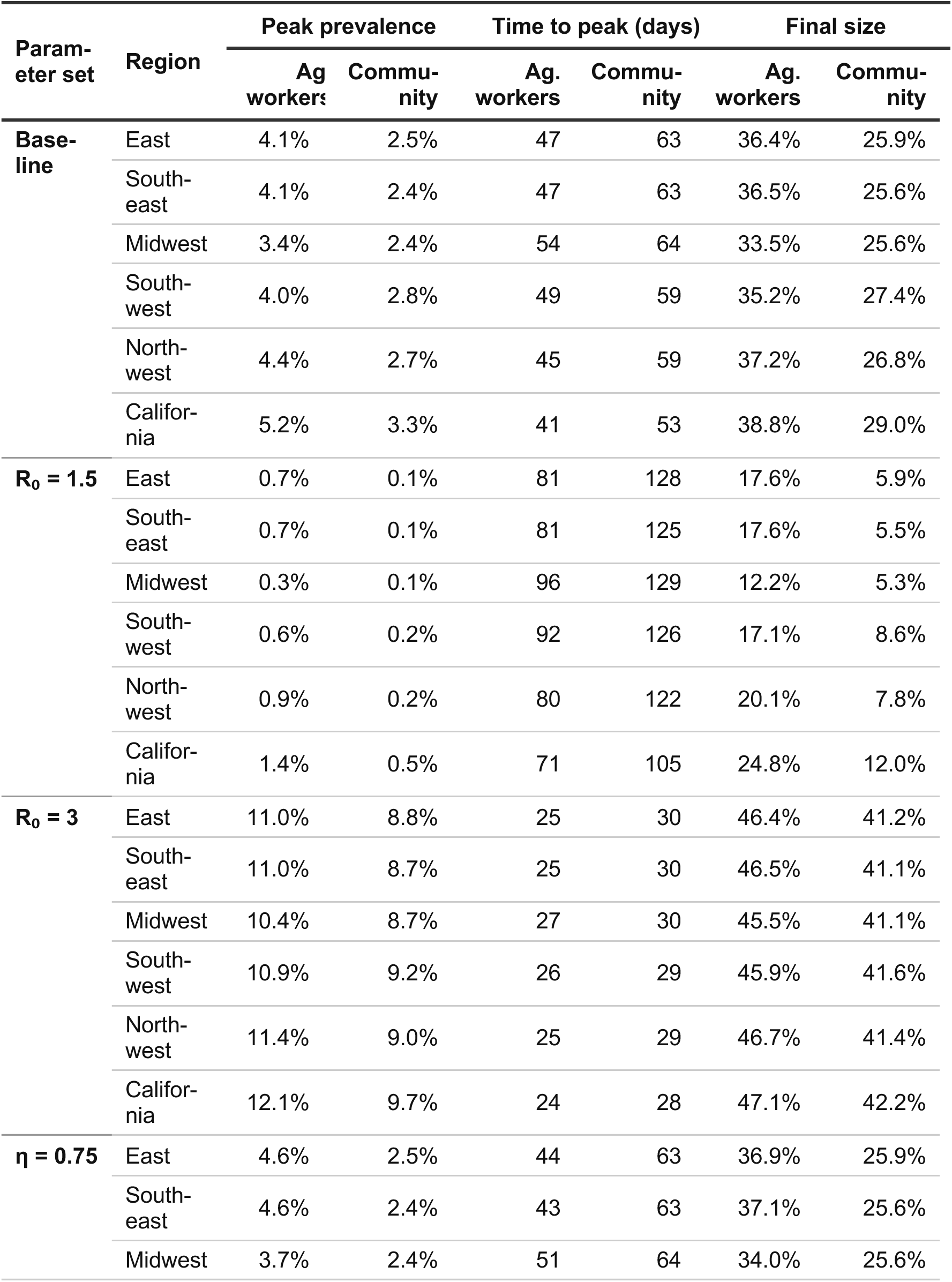
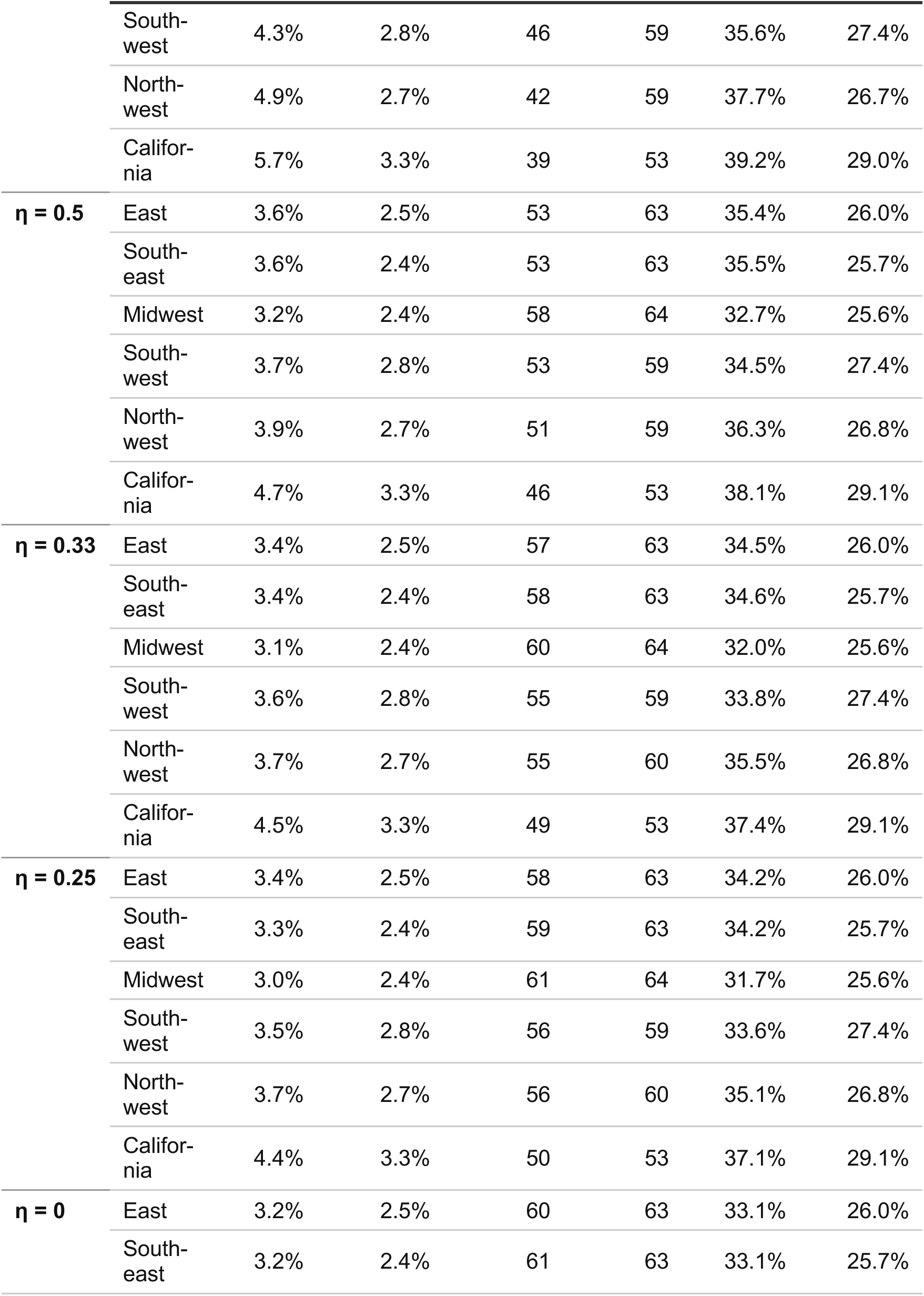
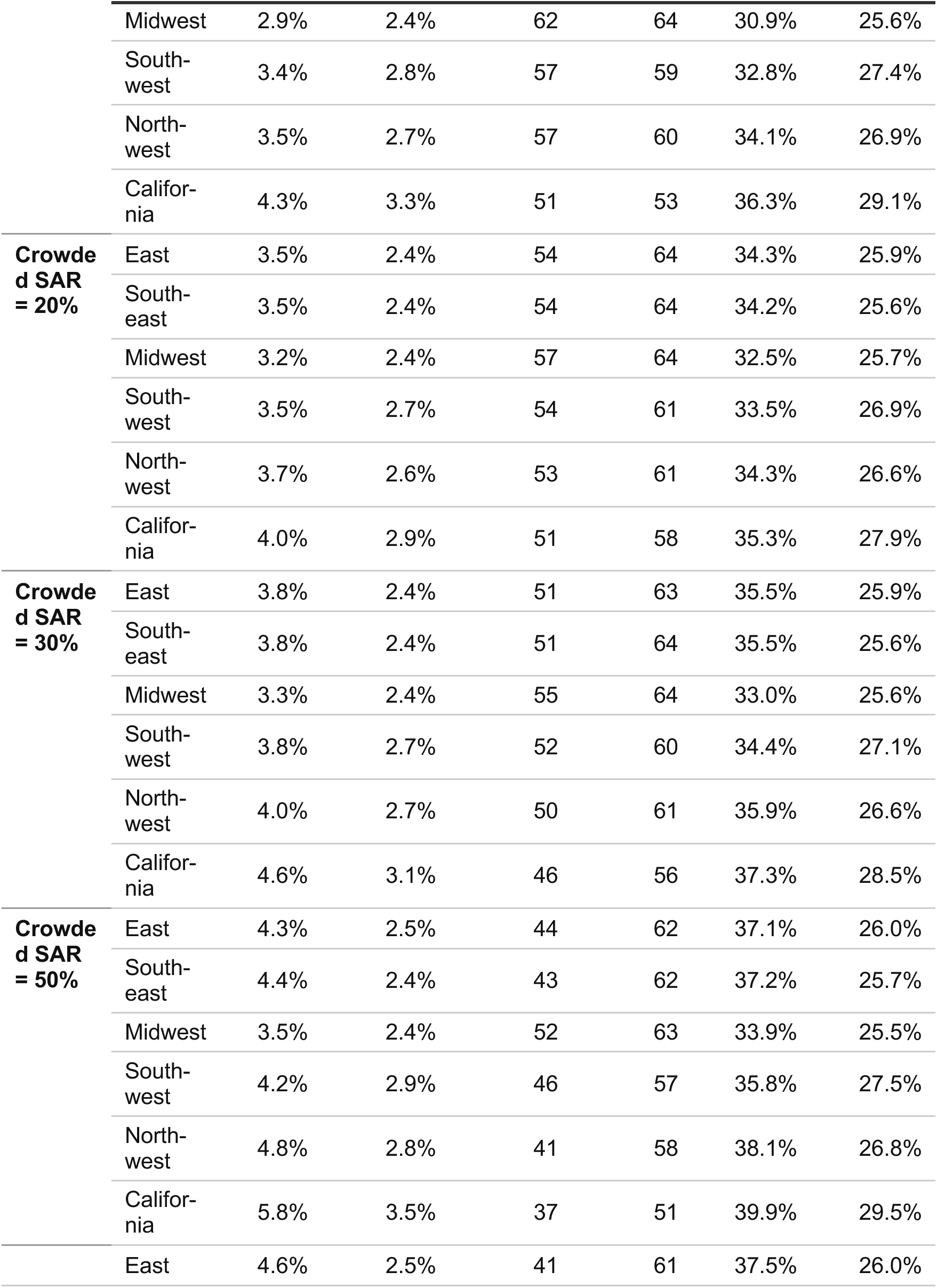
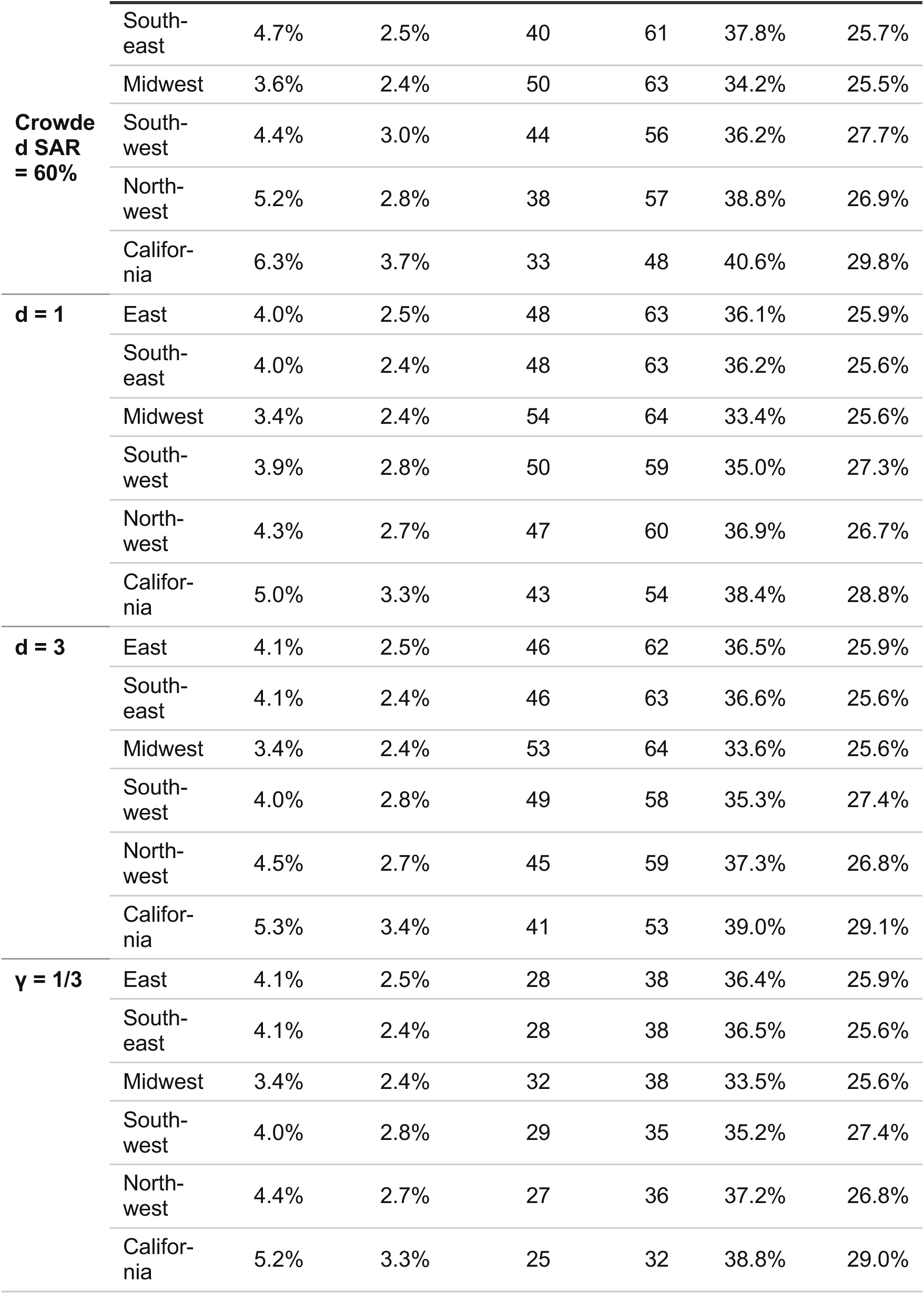
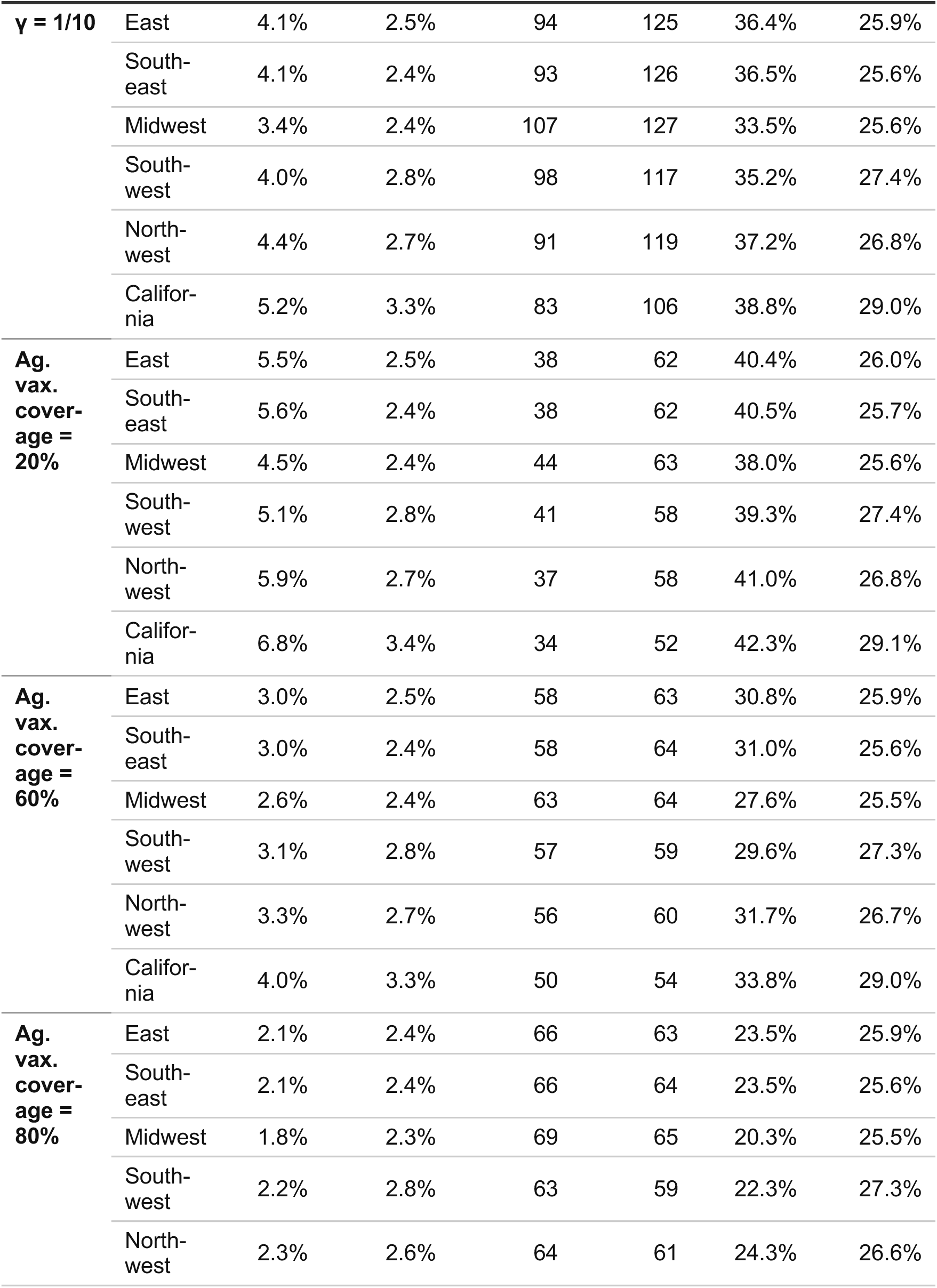
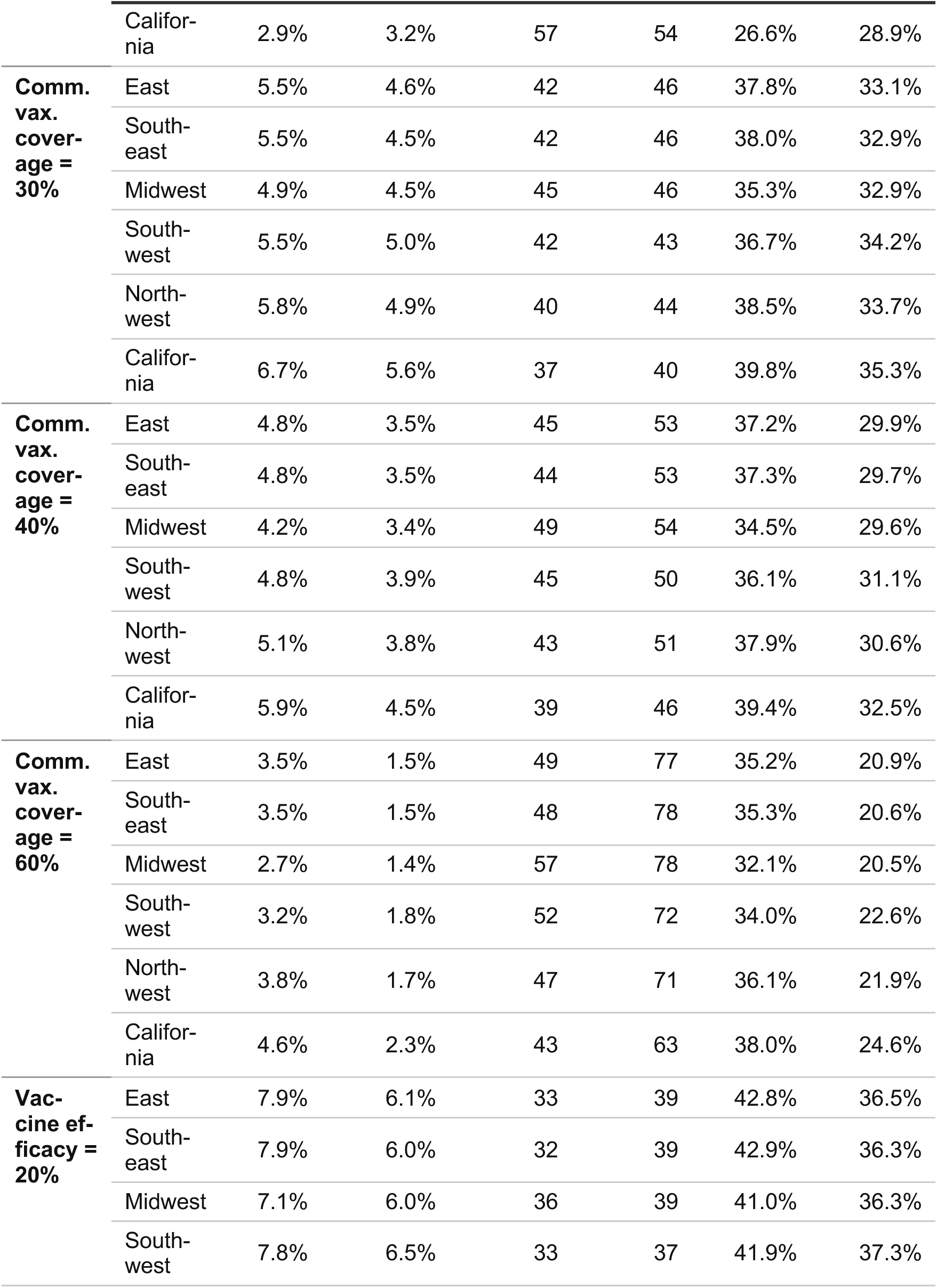
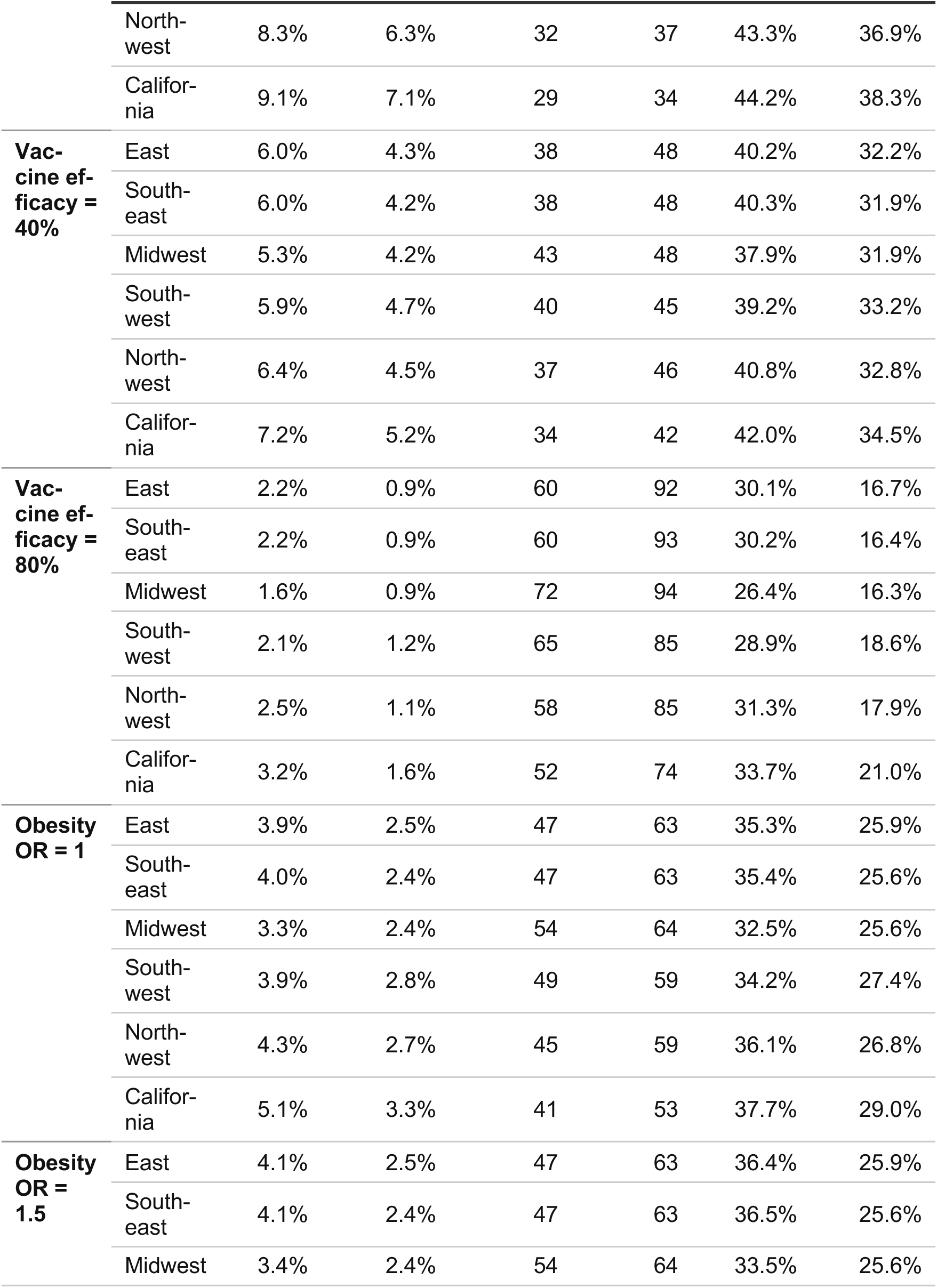
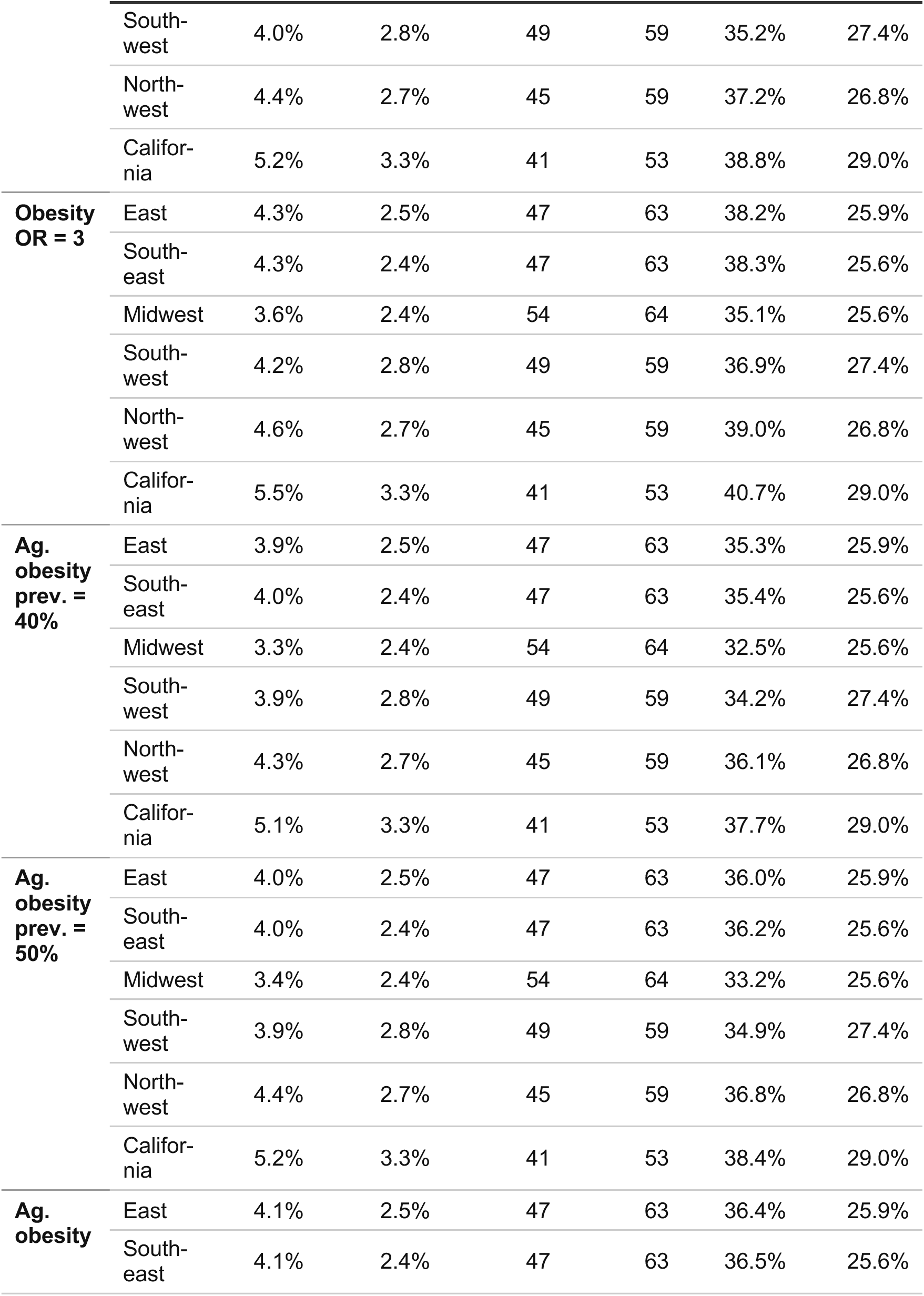
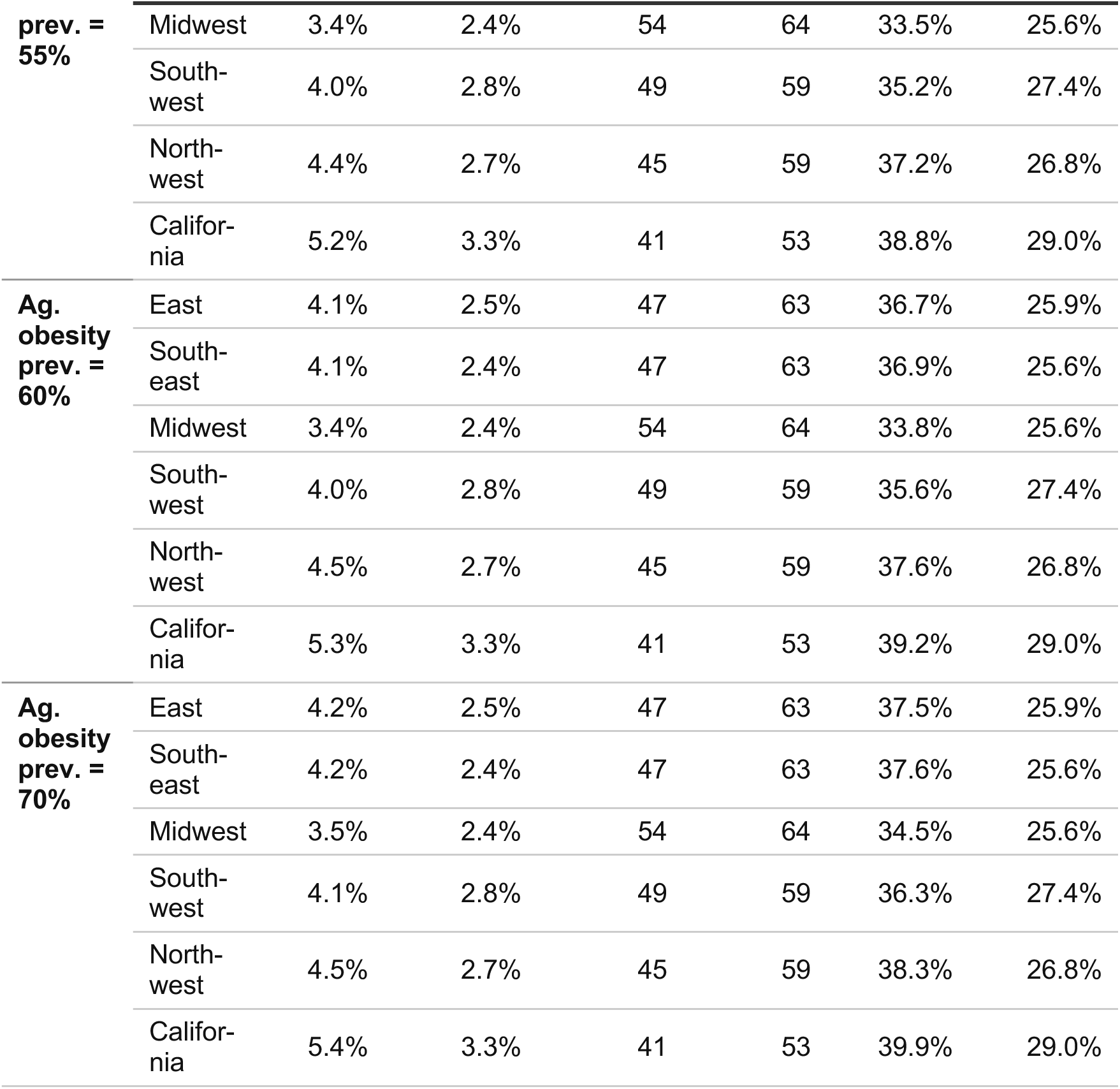
Summary statistics for simulated epidemics across regions and parameter sets. Simulated peak prevalence, time to epidemic peak, and final symptomatic attack rate (final size) for agricultural workers (Ag. workers) and the general community (Community). The baseline uses *R*0 = 2.0; each other row varies a single parameter (**Supplementary Table S2**).

**Table S4.**
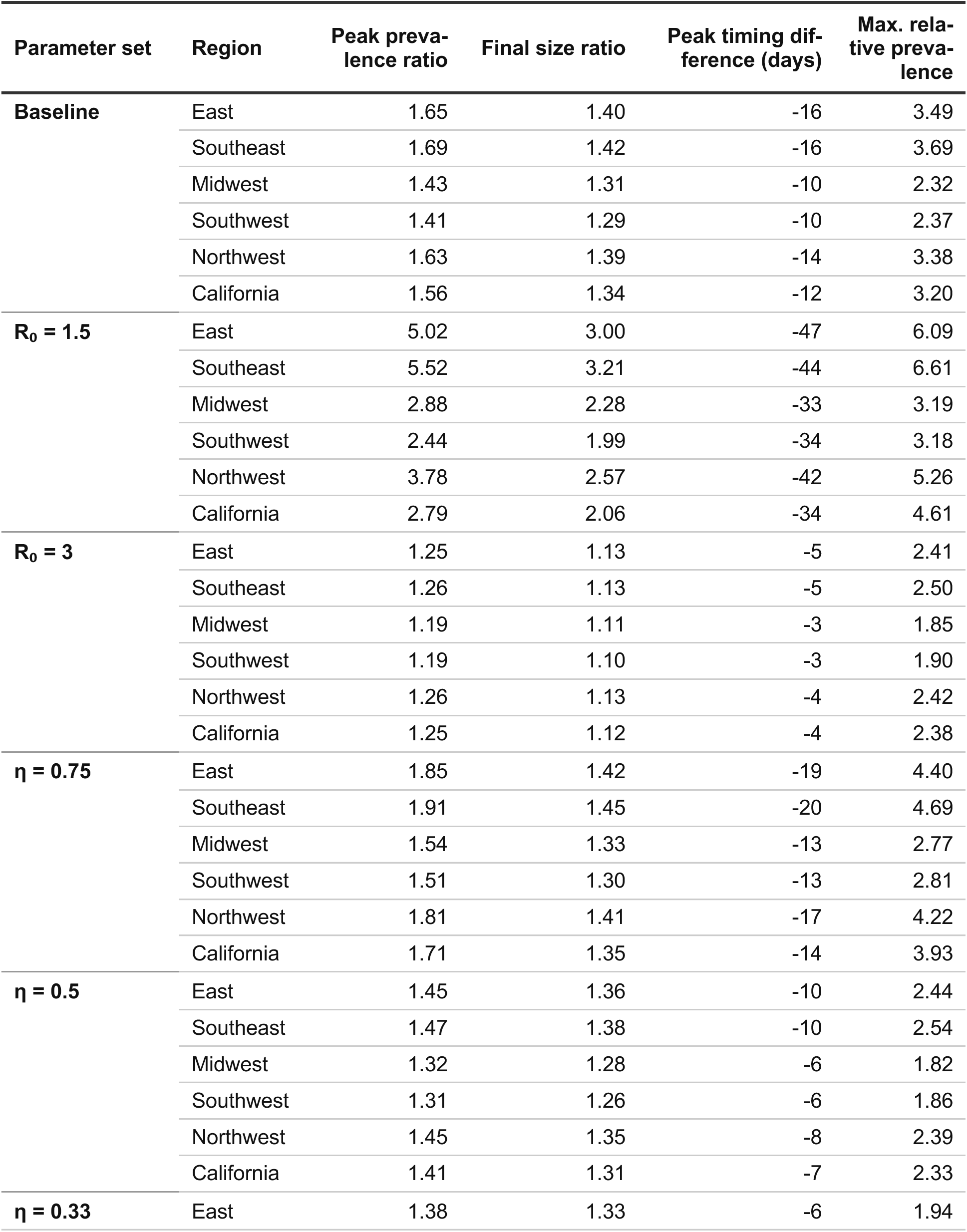
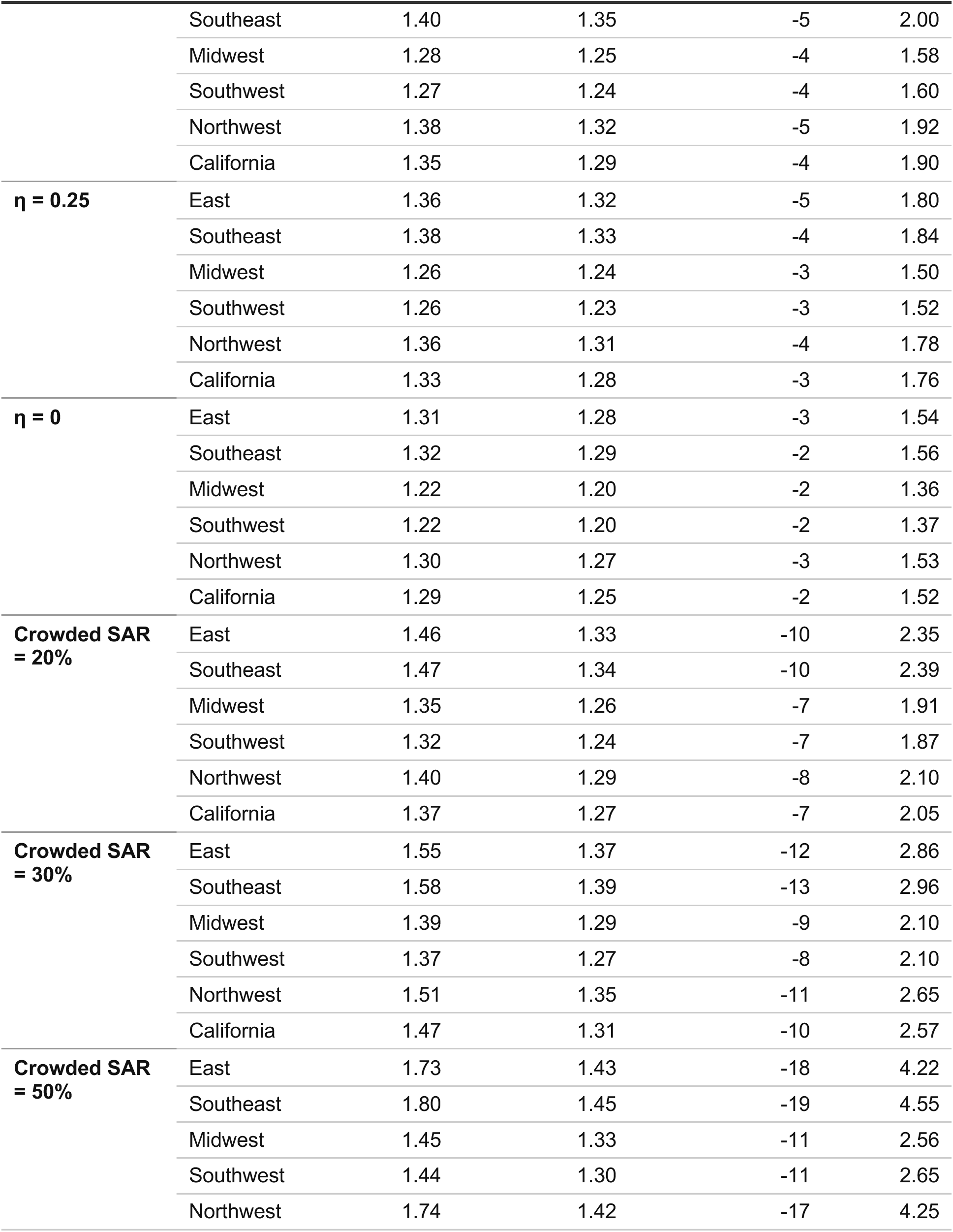
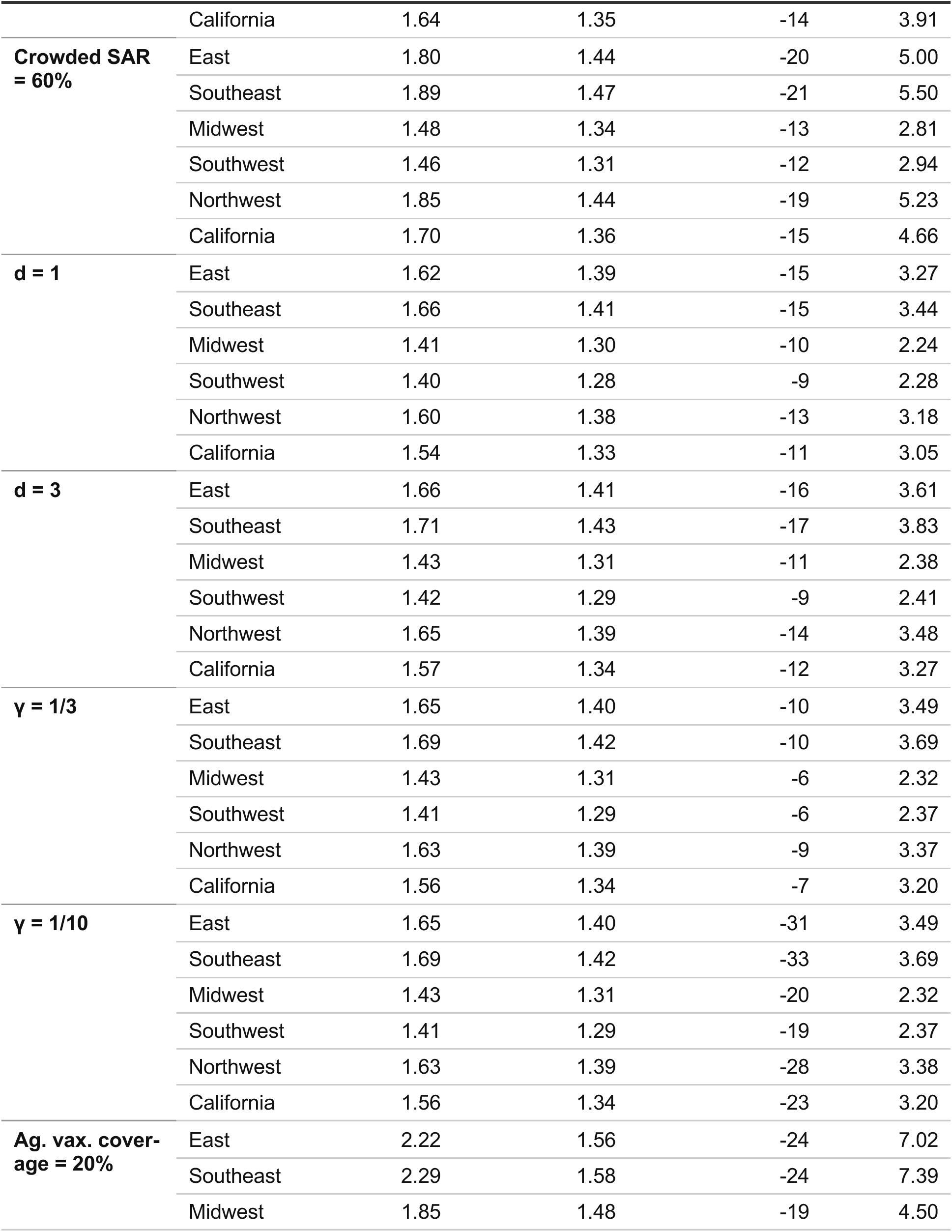
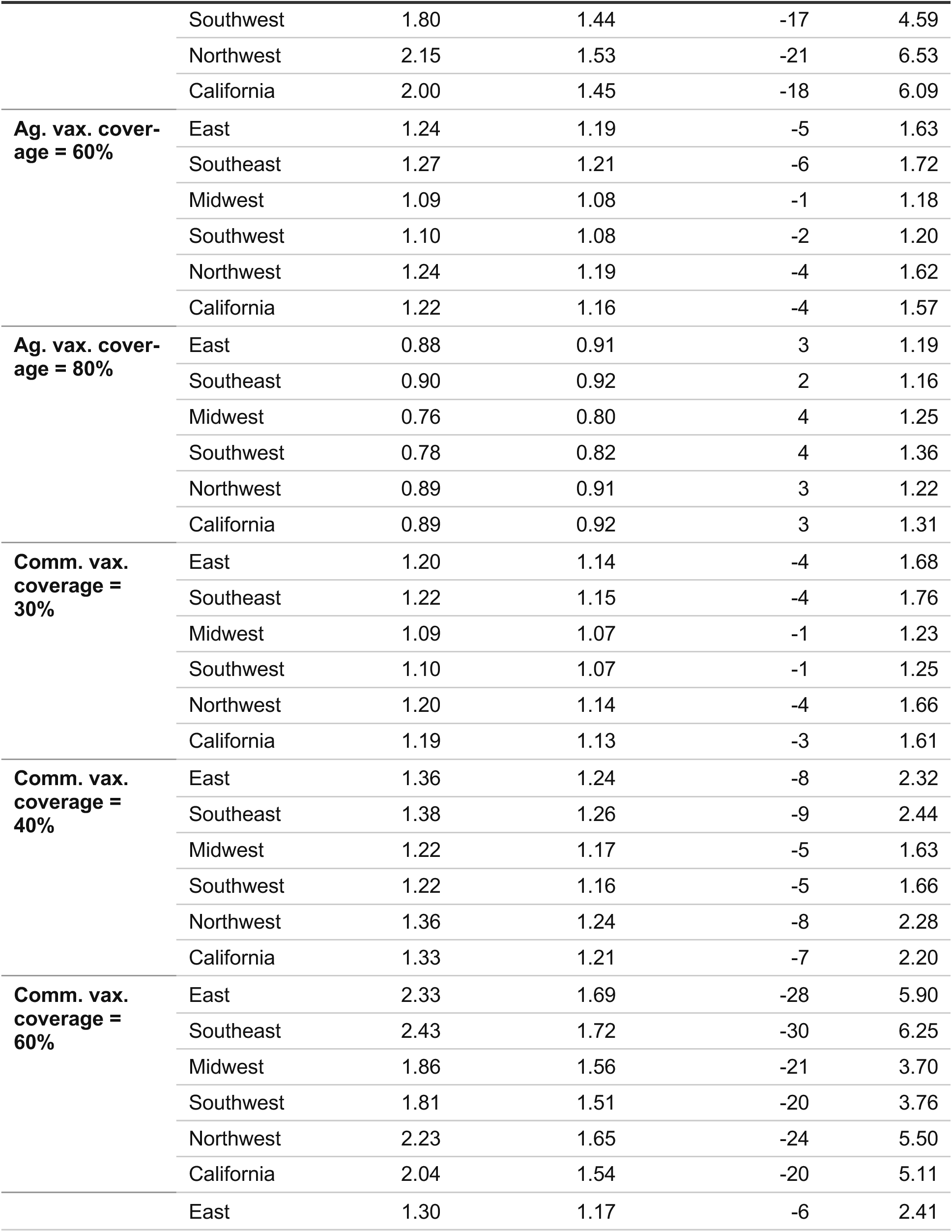
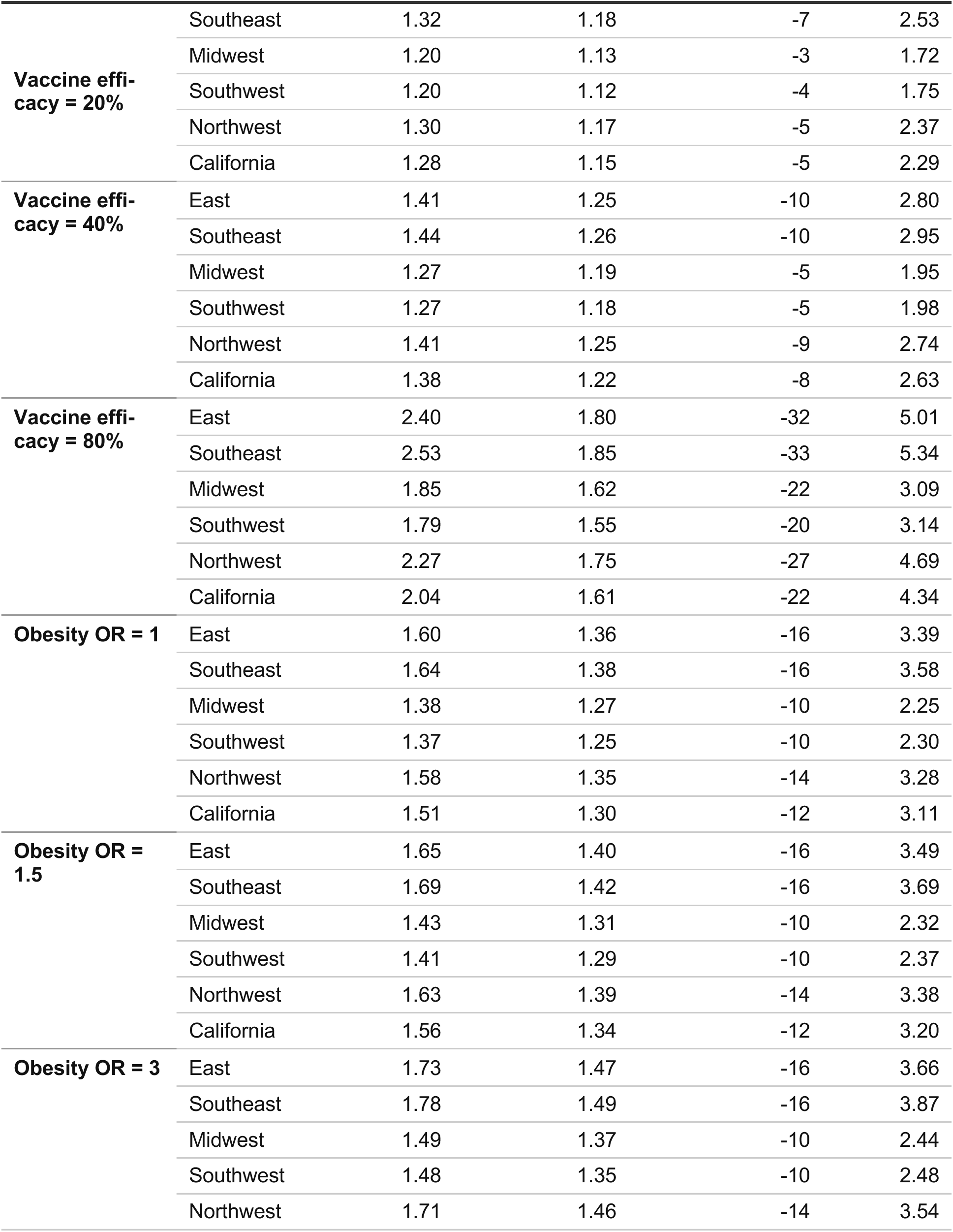
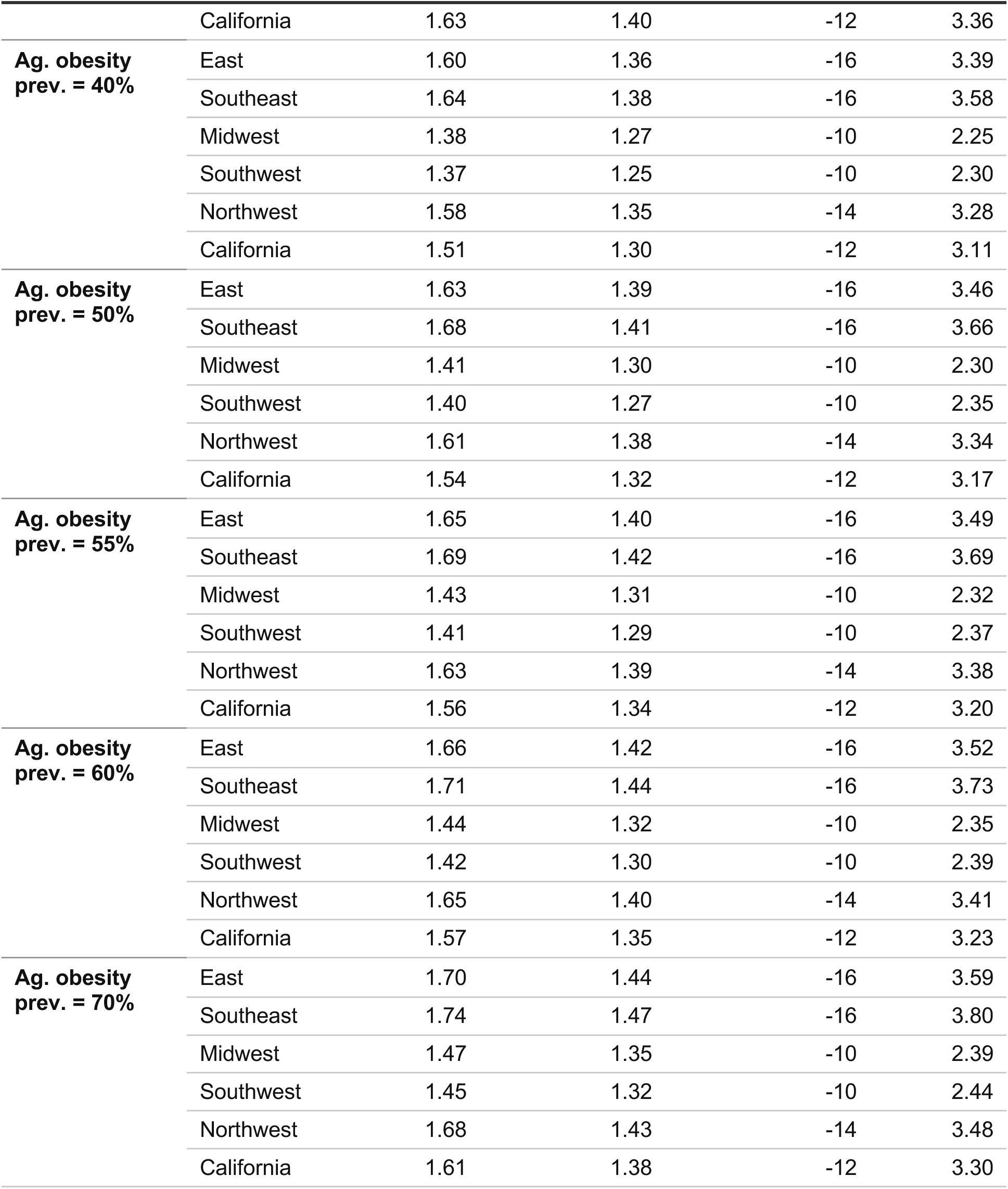
Differential metrics between agricultural workers and the general community across regions and parameter sets. Peak prevalence ratio, final size ratio, peak timing difference, and maximum relative prevalence between agricultural workers and the general community. The baseline uses *R*0 = 2.0.

**Table S5.** Symptomatic case rates by model scenario and NAWS region. Each scenario represents a set of model parameters that are allowed to vary between the agricultural worker sub-population and the general community; all other parameters are held fixed across both sub-populations at their general-community levels.

| Scenario | Region | Symptomatic Case Rate, Ag Workers | Symptomatic Case Rate, Community | Difference | % of Full-Model Disparity |
| --- | --- | --- | --- | --- | --- |
| **Null (all equal)** | East | 25.9% | 25.9% | 0.0% | 0% |
|  | Southeast | 25.6% | 25.6% | 0.0% | 0% |
|  | Midwest | 25.5% | 25.5% | 0.0% | 0% |
|  | Southwest | 27.3% | 27.3% | 0.0% | 0% |
|  | Northwest | 26.7% | 26.7% | 0.0% | 0% |
|  | California | 29.0% | 29.0% | 0.0% | 0% |
| **Household structure only** | East | 32.8% | 25.9% | 6.9% | 66% |
|  | Southeast | 33.0% | 25.6% | 7.3% | 68% |
|  | Midwest | 29.8% | 25.6% | 4.3% | 54% |
|  | Southwest | 31.6% | 27.4% | 4.3% | 55% |
|  | Northwest | 33.7% | 26.7% | 6.9% | 67% |
|  | California | 35.4% | 29.0% | 6.4% | 66% |
| **Vaccination disparity only** | East | 28.7% | 25.9% | 2.8% | 27% |
|  | Southeast | 28.4% | 25.6% | 2.8% | 26% |
|  | Midwest | 28.3% | 25.5% | 2.8% | 35% |
|  | Southwest | 30.1% | 27.3% | 2.7% | 35% |
|  | Northwest | 29.4% | 26.7% | 2.7% | 26% |
|  | California | 31.6% | 29.0% | 2.6% | 27% |
| **Obesity disparity only** | East | 26.7% | 25.9% | 0.8% | 8% |
|  | Southeast | 26.4% | 25.6% | 0.8% | 7% |
|  | Midwest | 26.3% | 25.5% | 0.8% | 10% |
|  | Southwest | 28.2% | 27.3% | 0.8% | 11% |
|  | Northwest | 27.5% | 26.7% | 0.8% | 8% |
|  | California | 29.8% | 29.0% | 0.9% | 9% |
| **HH structure + Obesity** | East | 33.8% | 25.9% | 7.9% | 76% |
|  | Southeast | 34.0% | 25.6% | 8.3% | 77% |
|  | Midwest | 30.7% | 25.6% | 5.2% | 65% |
|  | Southwest | 32.6% | 27.4% | 5.2% | 67% |
|  | Northwest | 34.7% | 26.7% | 8.0% | 76% |
|  | California | 36.5% | 29.0% | 7.5% | 77% |
| **HH + Vaccination (no obesity)** | East | 35.3% | 25.9% | 9.4% | 90% |
|  | Southeast | 35.4% | 25.6% | 9.8% | 90% |
|  | Midwest | 32.5% | 25.6% | 6.9% | 88% |
|  | Southwest | 34.2% | 27.4% | 6.8% | 87% |
|  | Northwest | 36.1% | 26.8% | 9.3% | 90% |
|  | California | 37.7% | 29.0% | 8.6% | 88% |
| **Vaccination + Obesity** | East | 29.6% | 25.9% | 3.6% | 35% |
|  | Southeast | 29.3% | 25.6% | 3.6% | 34% |
|  | Midwest | 29.2% | 25.5% | 3.6% | 46% |
|  | Southwest | 31.0% | 27.3% | 3.6% | 46% |
|  | Northwest | 30.3% | 26.7% | 3.6% | 35% |
|  | California | 32.6% | 29.0% | 3.6% | 37% |
| **Full model (all factors)** | East | 36.4% | 25.9% | 10.5% | 100% |
|  | Southeast | 36.5% | 25.6% | 10.9% | 100% |
|  | Midwest | 33.5% | 25.6% | 7.9% | 100% |
|  | Southwest | 35.2% | 27.4% | 7.8% | 100% |
|  | Northwest | 37.2% | 26.8% | 10.4% | 100% |
|  | California | 38.8% | 29.0% | 9.8% | 100% |

Case rates are symptomatic fractions (infections × $p_{\text{symp}}$ ) of the regional population. “% of Full-Model Disparity” is the ag worker – community difference as a percentage of the Full Model (all factors) difference within each region. Scenarios with “no obesity” apply a common $p_{\text{symp}}$ to both subpopulations; all others apply subpopulation-specific $p_{\text{symp}}$ .

**Table S6.** Estimated harvest-related crop production losses due to epidemic-induced workforce illness across R₀ values and symptomatic proportions. For each crop we report the worst-case epidemic peak timing, the corresponding maximum production loss as a percentage of total annual production, and the estimated dollar value of that loss based on 2024 California crop values, at the baseline symptomatic probability (p_symp,A_ adjusting for obesity) and assuming all infections are symptomatic (p_symp,A_ = 1).

| $R_0$ | $p_{\text{symp},A}$ | Crop | 2024 value (USD) | Worst peak day | Month | Max loss (%) | Max loss (USD) |
| --- | --- | --- | --- | --- | --- | --- | --- |
| 1.5 | 0.52 | Lettuce, Ice-berg | \$1,245,105,000 | 191 | July | 0.31% | \$3,906,375 |
| 1.5 | 0.52 | Oranges | \$852,507,000 | 77 | March | 0.30% | \$2,543,164 |
| 1.5 | 0.52 | Strawberries | \$3,456,522,000 | 181 | June | 0.35% | \$12,228,765 |
| 2 | 0.52 | Lettuce, Ice-berg | \$1,245,105,000 | 152 | June | 0.51% | \$6,316,496 |
| 2 | 0.52 | Oranges | \$852,507,000 | 35 | February | 0.50% | \$4,304,130 |
| 2 | 0.52 | Strawberries | \$3,456,522,000 | 150 | May | 0.63% | \$21,607,084 |
| 3 | 0.52 | Lettuce, Ice-berg | \$1,245,105,000 | 142 | May | 0.62% | \$7,729,725 |
| 3 | 0.52 | Oranges | \$852,507,000 | 12 | January | 0.64% | \$5,486,051 |
| 3 | 0.52 | Strawberries | \$3,456,522,000 | 148 | May | 0.78% | \$26,912,547 |
| 1.5 | 1 | Lettuce, Ice-berg | \$1,245,105,000 | 191 | July | 0.61% | \$7,583,032 |
| 1.5 | 1 | Oranges | \$852,507,000 | 77 | March | 0.58% | \$4,936,775 |
| 1.5 | 1 | Strawberries | \$3,456,522,000 | 181 | June | 0.69% | \$23,738,406 |
| 2 | 1 | Lettuce, Ice-berg | \$1,245,105,000 | 152 | June | 0.98% | \$12,261,545 |
| 2 | 1 | Oranges | \$852,507,000 | 35 | February | 0.98% | \$8,355,151 |
| 2 | 1 | Strawberries | \$3,456,522,000 | 150 | May | 1.21% | \$41,943,545 |
| 3 | 1 | Lettuce, Ice-berg | \$1,245,105,000 | 142 | May | 1.21% | \$15,004,897 |
| 3 | 1 | Oranges | \$852,507,000 | 12 | January | 1.25% | \$10,649,490 |
| 3 | 1 | Strawberries | \$3,456,522,000 | 148 | May | 1.51% | \$52,242,478 |

